# Genomic insights into substance use and disinhibitory disorders

**DOI:** 10.64898/2026.02.09.26344198

**Authors:** Camille M. Williams, Holly E. Poore, Diego Londono-Correa, Yuchen Ning, Natasia S. Courchesne-Krak, Maia Choi, Justin D. Tubbs, Matthew Rosenblatt, Peter T. Tanksley, Fazil Aliev, Nathaniel Y. Bell, Joseph D. Deak, Jean Gonzalez, Mariela Jennings, Emma C. Johnson, Tanya N. Phung, Marijn Schipper, Ronald de Vlaming, Kai Yuan, Hang Zhou, Emily Balcke, Sarah J. Brislin, COGA Collaborators, Javier de la Fuente, Michael J. Gandal, Tian Ge, Joel Gelernter, Danielle Posthuma, Jordan W. Smoller, Elliot M. Tucker-Drob, Irwin D. Waldman, Abraham A. Palmer, Peter B. Barr, Sandra Sanchez-Roige, Richard Karlsson Linnér, Danielle M. Dick, K. Paige Harden, Travis T. Mallard

**Affiliations:** Laboratoire de Sciences Cognitives et Psycholinguistique, Département d′Études Cognitives, École Normale Supérieure, EHESS, CNRS, PSL University, Paris, France; Department of Psychology, University of Texas at Austin, Austin, TX, USA; Population Research Center, University of Texas at Austin, Austin, TX, USA; Department of Psychiatry, Rutgers Robert Wood Johnson Medical School, Piscataway, NJ, USA; Rutgers Addiction Research Center, Brain Health Institute, Rutgers Health, Piscataway, NJ, USA; Department of Psychiatry, Academic Medical Center, Amsterdam, The Netherlands; Department of Economics, School of Business and Economics, Vrije Universiteit Amsterdam, Amsterdam, The Netherlands; Department of Psychiatry, University of California, San Diego, CA, USA; Department of Psychology, School of Arts and Sciences, Rutgers University, NJ, USA; Center for Precision Psychiatry, Department of Psychiatry, Massachusetts General Hospital, Boston, MA, USA; Department of Psychiatry, Harvard Medical School, Massachusetts General Hospital, Boston, MA, USA; Psychiatric and Neurodevelopmental Genetics Unit, Center for Genomic Medicine, Massachusetts General Hospital, Boston, MA, USA; Stanley Center for Psychiatric Research, Broad Institute of MIT and Harvard, Boston, MA, USA; ALERRT Center, Texas State University, San Marcos, TX, USA; Department of Complex Trait Genetics, Center for Neurogenomics and Cognitive Research, Amsterdam Neuroscience, Vrije Universiteit Amsterdam, Amsterdam, The Netherlands; Department of Psychiatry, Yale School of Medicine, New Haven, CT, USA; Veterans Affairs Connecticut Healthcare System, West Haven, CT, USA; Department of Psychiatry, Washington University School of Medicine, St Louis, MO, USA; Department of Econometrics and Data Science, School of Business and Economics, Vrije Universiteit Amsterdam, Amsterdam, The Netherlands; Analytic and Translational Genetics Unit, Massachusetts General Hospital, Boston, MA, USA; Department of Medicine, Harvard Medical School, Boston, MA, USA; Department of Biomedical Informatics and Data Science, Yale School of Medicine, New Haven, CT, USA; Center on Aging and Population Sciences, University of Texas at Austin, Austin, TX, USA; Lifespan Brain Institute at Penn Med and the Children’s Hospital of Philadelphia, University of Pennsylvania, Philadelphia, PA, USA; Department of Genetics, Yale School of Medicine, New Haven, CT, USA; Department of Neuroscience, Yale School of Medicine, New Haven, CT, USA; Department of Psychology, Emory University, Atlanta, GA, USA; Institute for Genomic Medicine, University of California, San Diego, CA, USA; Department of Psychiatry and Behavioral Sciences, State University of New York Downstate Health Sciences University, Brooklyn, NY, USA; Department of Community Health Sciences, School of Public Health, State University of New York Downstate Health Sciences University, Brooklyn, NY, USA; Institute for Genomics in Health, State University of New York Downstate Health Sciences University, Brooklyn, NY, USA; VA New York Harbor Healthcare System, Brooklyn, NY, USA; Division of Genetic Medicine, Vanderbilt University Medical Center, Nashville, TN, USA; Department of Economics, Leiden Law School, Universiteit Leiden, Leiden, The Netherlands

## Abstract

Externalizing spectrum disorders—spanning attention-deficit/hyperactivity disorder, conduct disorder, substance use disorders, and other disorders characterized by disinhibition—frequently co-occur within individuals due, in part, to shared genetic etiology. To advance understanding of this genetic architecture, we conducted a multi-ancestry, multivariate genome-wide association analysis of more than 4 million individuals, identifying 1,294 genomic regions linked to an externalizing factor. Fine-mapping and gene prioritization efforts identified 961 effector genes, with the putative causal variant associations showing robust replication in the All of Us Research Program sample. Bioinformatic analyses revealed a broadly distributed neural architecture with early and sustained involvement of GABAergic and glutamatergic neurons. Drug repurposing analyses further highlighted the role of GABA_A_ receptors, as well as dopaminergic signaling, excitatory-inhibitory balance, and neurosteroid pathways. A genome-wide polygenic index predicted ∼12% of the variance in externalizing in independent cohorts of individuals with European-like ancestry, compared to ∼3% in individuals with African-like ancestry, and was associated with myriad health and life outcomes. Together, these findings map the shared genetic etiology of externalizing psychopathology and identify neurodevelopmental and synaptic mechanisms with translational relevance.

---

Multiple psychiatric disorders are characterized by persistent difficulty regulating impulsive thoughts and actions. These disorders include attention-deficit/hyperactivity disorder (ADHD), conduct disorder (CD), antisocial personality disorder (ASPD), and substance use disorders (SUDs). Together, disinhibitory disorders and related behaviors comprise the **externalizing spectrum**^1–3^, which encompasses many of the world’s most common and costly public health burdens. In the United States alone, excessive alcohol use is estimated to cost $250 billion per year^4^, and illicit drug use another $190 billion^5^. Among children and adolescents, ADHD is associated with over $150 billion in annual costs^6,7^, while CD is one of the most common reasons that youth are referred to mental health services^8^.

Despite the profound impact of externalizing psychopathology, there are few effective treatments. Pharmacological therapies for ADHD stand alone in the externalizing spectrum; currently, no FDA-approved medications exist for CD or ASPD^9^, while available pharmacological treatments for SUDs typically focus on reducing substance-specific craving and withdrawal symptoms and are only modestly effective and infrequently used^10,11^. Existing psychosocial interventions also tend to achieve modest effects^12–14^, highlighting the pressing need for novel therapeutic approaches.

Externalizing spectrum disorders are highly comorbid^15^. Twin- and family-based studies suggest that this comorbidity is primarily driven by shared genetic influences^2^. Genomic studies, therefore, hold promise for illuminating etiological mechanisms shared across the externalizing spectrum, and for identifying novel targets for pharmacological and psychosocial treatments that modulate core disinhibitory processes^16,17^. However, until recently, the pace of genetic discovery has been slow, in part due to the difficulty of amassing sufficient data from diverse populations for any single disorder.

We previously addressed this challenge with a multivariate genome-wide association study (GWAS) that pooled data on approximately 1.5 million people by (i) aggregating data across multiple externalizing behaviors and disorders, and (ii) incorporating data on non-clinical health-risk behaviors that also reflect difficulties with disinhibition^18^. Here, we report results from a pre-registered GWAS of externalizing (https://osf.io/7pfgj/) that more than doubled the sample size to 4 million people, including individuals of European-like (EUR-like) and African-like (AFR-like) ancestries. In addition to substantially advancing genetic discovery, our analyses reveal novel neurodevelopmental mechanisms of risk for externalizing spectrum disorders, identify potential drug repurposing candidates, create one of the most robustly predictive psychiatric polygenic indices to date, and demonstrate the utility of genetic data for studying myriad adverse psychiatric, social, and medical outcomes across the entire human lifespan. Overall, our study provides new insights into the etiology of a stigmatized constellation of psychiatric disorders that impose a profound burden on human health and well-being.

## Results

### Modeling the shared genetic architecture of the externalizing spectrum

We applied Genomic Structural Equation Modeling (Genomic SEM)^19^ to GWAS data from seven phenotypes, estimating a single latent factor of externalizing liability in EUR-like^20^ individuals (**Table 1**). The phenotypes were: (i) attention-deficit/hyperactivity disorder (ADHD)^21^, (ii) problematic alcohol use (ALCP)^22^, (iii) cannabis use disorder (CUD)^23^, (iv) reverse-coded age at first sexual intercourse (FSEX)^18^, (v) number of sexual partners (NSEX)^18^, (vi) self-reported risk tolerance (RISK)^18^, and (vii) lifetime smoking initiation (SMOK)^24^. Phenotypes were selected to be consistent with theory and previous research^18^, and to maximize sample size (**Supplementary Table 1**). We previously demonstrated that sample-size differences among indicators do not unduly influence model estimates of the joint polygenic architecture^18,19^. The common factor approach captures genetic liability shared across all indicators, and larger sample sizes increase power to detect single-nucleotide polymorphism (SNP) associations.

**Table 1.** Summary of the externalizing phenotypes included in the discovery GWAS.

|  | European (EUR)-like ancestry group |  |  |  |  |  |  | African (AFR)-like ancestry group |  |  |  |  |  |  |
| --- | --- | --- | --- | --- | --- | --- | --- | --- | --- | --- | --- | --- | --- | --- |
| | *N* | *h*<sup>2</sup> (s.e.) | $\lambda_{GC}$ | Mean $\chi^2$ | Intercept | Ratio | Ref | *N* | *h*<sup>2</sup> (s.e.) | $\lambda_{GC}$ | Mean $\chi^2$ | Intercept | Ratio | Ref |
| **ADHD** | 103,136 | 0.176 (0.008) | 1.355 | 1.449 | 1.026 | 0.058 | <sup>21</sup> | — | — | — | — | — | — | — |
| **ALCP** | 352,373 | 0.062 (0.003) | 1.368 | 1.509 | 1.022 | 0.042 | <sup>22</sup> | 105,433 | 0.111 (0.021) | 1.11 | 1.124 | 1.039 | 0.318 | <sup>22</sup> |
| **CUD** | 141,943 | 0.071 (0.004) | 1.23 | 1.279 | 0.994 | -0.023 | <sup>23</sup> | 59,641 | 0.05 (0.01) | 1.073 | 1.083 | 1.033 | 0.401 | <sup>23</sup> |
| **FSEX** | 357,187 | 0.118 (0.004) | 1.624 | 1.868 | 1.021 | 0.024 | <sup>18</sup> | — | — | — | — | — | — | — |
| **NSEX** | 336,121 | 0.101 (0.004) | 1.492 | 1.674 | 1.018 | 0.027 | <sup>18</sup> | — | — | — | — | — | — | — |
| **RISK** | 426,379 | 0.098 (0.005) | 1.369 | 1.459 | 1.012 | 0.026 | <sup>18</sup> | — | — | — | — | — | — | — |
| **SMOK** | 2,557,182 | 0.135 (0.004) | 3.533 | 5.601 | 1.011 | 0.002 | <sup>24</sup> | 94,299 | 0.142 (0.02) | 1.169 | 1.174 | 1.084 | 0.481 | <sup>24</sup> |

Statistics were derived from LD Score regression in Genomic SEM, using ancestry-specific HapMap3 SNPs with MAF $\geq 0.01$ for EUR-like data and MAF $\geq 0.05$ for AFR-like data. SNP-based heritability estimates ( $h^2$ ) are reported on the liability scale when appropriate. The genomic inflation factor, $\lambda_{GC}$ , is the median $\chi^2$ statistic divided by the expected median of the $\chi^2$ distribution with 1 d.f. Mean $\chi^2$ refers to the average $\chi^2$ statistic. The intercept is the estimated LD Score regression intercept. The ratio measures stratification bias, defined as: (intercept - 1)/(mean $\chi^2$ - 1). *N* corresponds to the maximum number of participants and refers to $N_{\text{eff}}$ (sum of effective sample size)<sup>25</sup> for binary traits. An em dash (—) means there is no applicable value.

In line with previous work^18^, genetic correlations among the seven phenotypes were moderate to high (**Fig. 1a**). In EUR-like individuals, a single factor model fit the data well (χ^2^(14) = 446.528; CFI = 0.941; SRMR = 0.067; **Fig. 1b**), supporting the assertion that these diverse phenotypes are all influenced by a shared underlying liability (**Supplementary Table 4**). The genetic correlation between this externalizing (EXT_EUR_) genomic factor and our previously published externalizing GWAS was indistinguishable from unity (*r*_g_ = 0.990, s.e. = 0.036). This result indicates that the factor analytic approach robustly identifies the same dimension of genetic liability, even when there are changes in individual indicators and relative sample sizes.

**Fig. 1.**
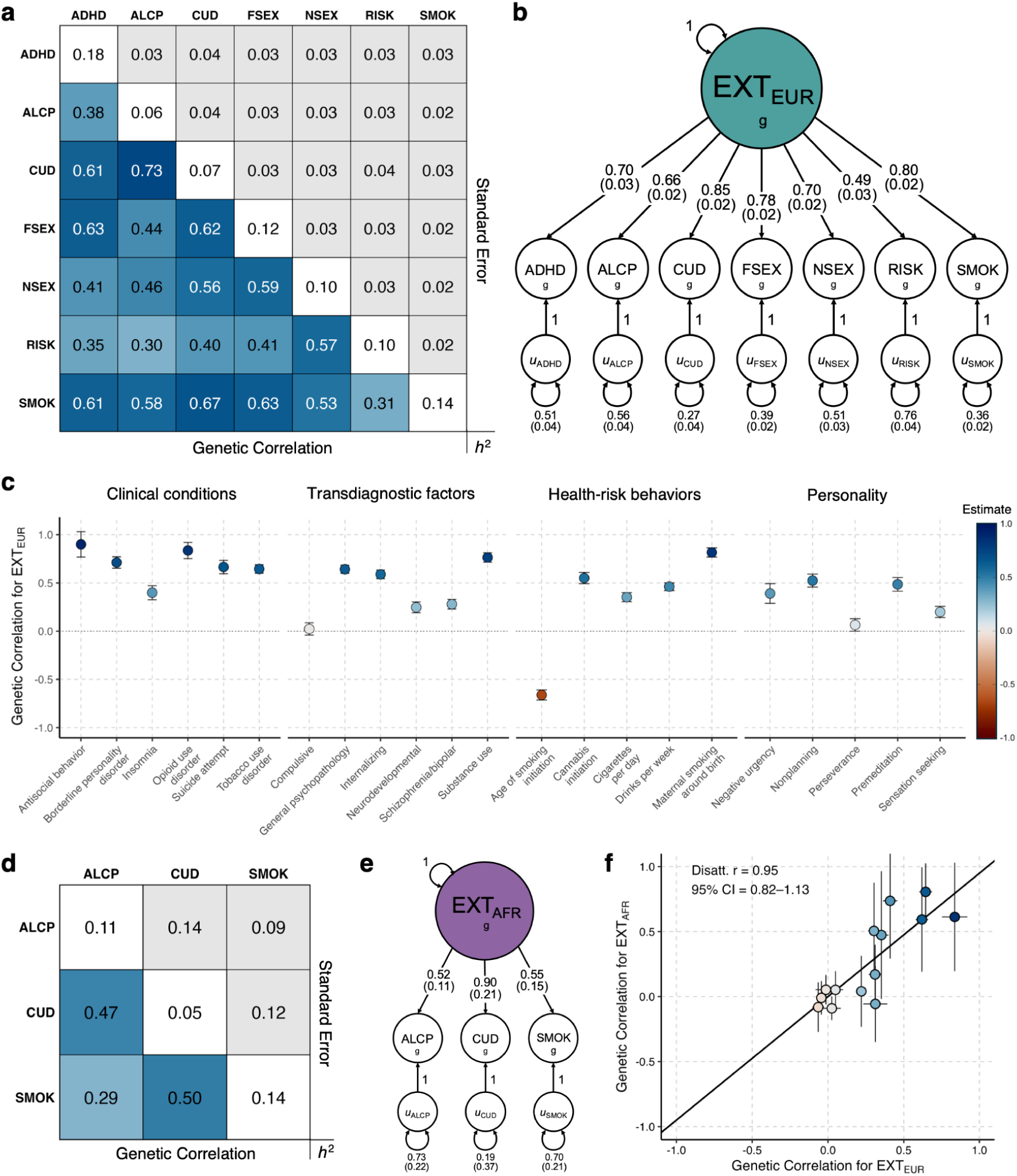
Genomic structural equation models of the externalizing spectrum in European-like and African-like ancestry groups. **a**,**d**, LD Score regression results for Genomic SEM indicators in European (EUR)-like (**a**) and African (AFR)-like (**d**) ancestry samples (**Table 1**). The lower triangle of the matrix displays the pairwise genetic correlation (*r*_g_) estimates, while the upper triangle displays the corresponding standard errors (s.e.). The SNP heritability (*h*^2^) estimates are reported on the diagonal, with liability-scale transformations applied when appropriate. **b**,**e** Path diagram of the common factor model estimated in EUR-like (**b**) and AFR-like (**e**) data. Standardized factor loadings are presented with the corresponding s.e. reported in parentheses. **c**, The *r*_g_ estimates between the externalizing (EXT) factor in EUR-like individuals and a selection of complex traits (**Supplementary Table 7**). Error bars represent 95% confidence intervals (CIs) centered on the *r*_g_ estimate, computed as 1.96 times the s.e. **f**, The correlation between the ancestry-specific *r*_g_ estimates of the externalizing genomic factor with 14 external phenotypes ordered by decreasing genetic correlation in AFR-like ancestry: tobacco use disorder, depression, opioid use disorder, post-traumatic stress disorder, schizophrenia, cigarettes per day, ischemic heart disease, breast cancer, total cholesterol, body mass index, height, COVID-19, prostate cancer, and LDL cholesterol (**Supplementary Table 8**; **Supplementary Information**). The solid line denotes the regression line with the slope equal to the disattenuated correlation coefficient and an intercept of 0.

We then estimated genetic correlations between the EXT_EUR_ factor and 95 medical, psychiatric, psychological, and social phenotypes, also using GWAS summary statistics from EUR-like individuals (**Fig. 1c**; **Supplementary Table 7**). As predicted, EXT_EUR_ was strongly genetically correlated with clinically significant forms of harmful disinhibition, which were not included as indicators in the factor model, including antisocial behavior (*r*_g_ = 0.899, s.e. = 0.067), opioid use disorder (OUD; *r*_g_ = 0.836, s.e. = 0.043), borderline personality disorder (BPD; *r*_g_ = 0.712, s.e. = 0.030), and suicide attempt (*r*_g_ = 0.665, s.e. = 0.035). These genetic correlations were equivalent to or larger than several standardized factor loadings in the model, demonstrating that we successfully identified a dimension of genetic liability as strongly related to clinically severe pathology as to any single discovery phenotype.

For AFR-like^20^ individuals, only three externalizing-related GWASs met inclusion criteria (**Supplementary Information**): (1) ALCP^22^, (2) CUD^23^, and (3) SMOK^24^ (**Supplementary Table 2**). As with EUR-like individuals, these phenotypes were positively genetically correlated (**Fig. 1d**). To evaluate whether a comparable factor could be modeled with this reduced set, we first tested whether, in EUR-like individuals, a truncated factor estimated with these three indicators converged on the same genetic signal as the full EXT_EUR_ factor estimated with all seven indicators. It did: the full and truncated EXT factors in EUR-like individuals were genetically correlated at unity (*r*_g_ = 1, s.e. = 0.029).

We subsequently estimated this three-indicator model in AFR-like individuals (EXT_AFR_, **Fig. 1e**; **Supplementary Table 4**), as well as genetic correlations between this factor and 14 phenotypes using GWAS summary statistics from AFR-like individuals (**Fig. 1f**). Only correlations with tobacco use disorder (TUD) and depression significantly differed from zero after Bonferroni correction. However, after correcting for error in both sets of ancestry-specific estimates, the genetic correlations of EXT with other phenotypes were highly concordant across ancestries (disattenuated *r* = 0.950, 95% CI: 0.816–1.127; **Fig. 1f**; **Methods**).

Finally, we used Popcorn^26^ to estimate the genetic correlation between the EXT_AFR_ and EXT_EUR_ factors. This estimate was not significantly different from 1 (*r*_g_ = 1.409 [unbounded estimate, **Supplementary Information**], s.e. = 0.296) and exceeded our pre-registered threshold for proceeding with downstream analyses, including multi-ancestry meta-analysis.

### Identifying putatively causal variants and effector genes

Using Genomic SEM, we estimated associations between autosomal SNPs and EXT, separately in EUR-like individuals (*N*_observations_ = 5,726,594; *N*_individuals_ ≥ 3,786,374) and AFR-like individuals (*N*_observations_ = 362,830; *N*_individuals_ ≥ 243,163) (**Supplementary Tables 1–2**). The EXT_EUR_ GWAS identified a strong polygenic signal (mean χ^2^ = 5.862, λ_GC_ = 3.707) with minimal bias from population stratification (LD score regression intercept = 1.043 [s.e. = 0.034], attenuation ratio = 0.009 [s.e. = 0.007]). The EXT_AFR_ GWAS also identified a discernible polygenic signal (mean χ^2^ = 1.160, λ_GC_ = 1.131, LD score regression intercept = 1.043 [s.e. = 0.013], attenuation ratio = 0.270 [s.e. = 0.079]), despite markedly lower power compared to the EXT_EUR_ GWAS. Given evidence that the EXT factors in each ancestry group were converging on the same genetic signal (**Supplementary Information**), we then used METAL^27^ to conduct a fixed-effects meta-analysis across populations. Pooling across ancestries, we identified 1,294 genomic regions significantly associated with EXT (**Fig. 2**; **Methods**).

**Fig. 2.**
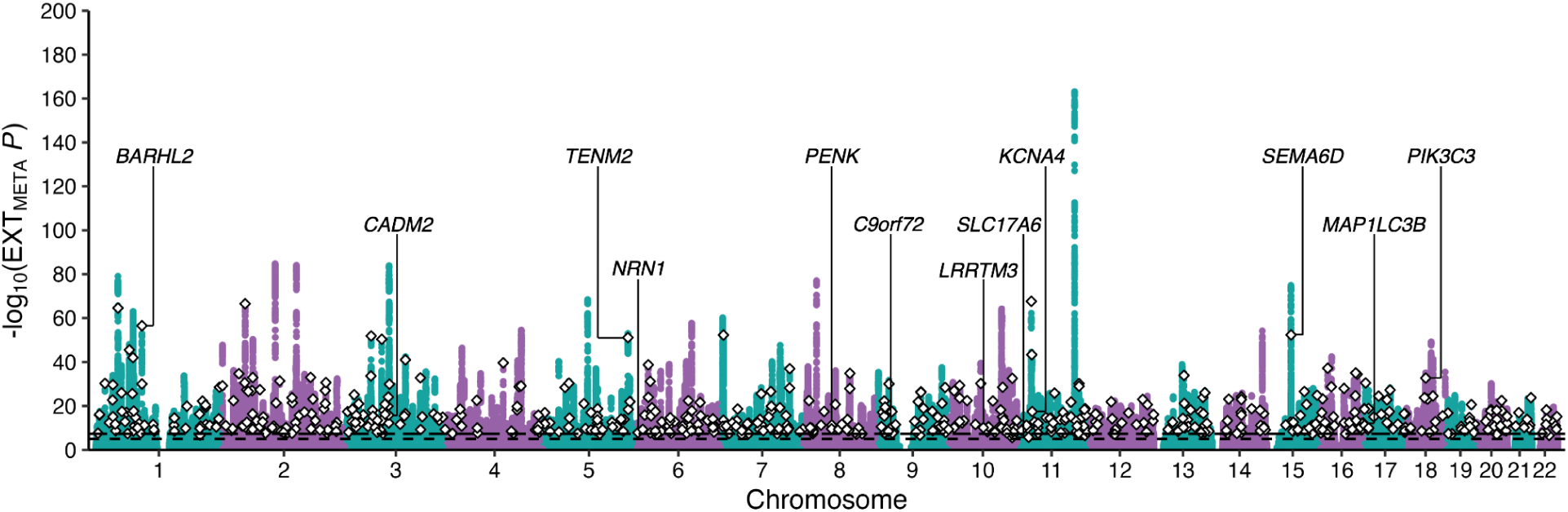
Multi-ancestry genome-wide association analysis of externalizing liability. Scatterplot of −log_10_ *P* values from a two-sided *Z* test in a fixed-effects meta-analysis of the EXT_EUR_ and EXT_AFR_ multivariate GWAS results. The x-axis indexes chromosomal position, the y-axis refers to statistical significance on a −log_10_ scale, the horizontal dashed line denotes genome-wide significance (*P* = 5e-8), and the horizontal dotted line marks suggestive significance (*P* = 1e-5). White diamonds represent the putative causal SNPs that were fine-mapped with moderate confidence (PIP ≥ 0.50) and had non-significant *Q*_SNP_ statistics (*Q*_SNP_ *P* > 5e-8 in both EXT_EUR_ and EXT_AFR_; **Supplementary Table 11**). Gene labels are reported for select effector genes (cumulative precision ≥ 0.75).

Using SuSiEx^28^, a Bayesian method for cross-population fine-mapping of shared and ancestry-specific GWAS signals, we identified 647 putative causal SNPs across 485 regions with a moderate level of confidence (posterior inclusion probability [PIP] ≥ 0.50), and 214 SNPs across 180 regions with high confidence (PIP ≥ 0.95). Inspection of *Q*_SNP_ signals in these regions revealed that only 1% (5/485) of moderate-confidence loci, and a single high-confidence locus, harbored a causal variant with significantly heterogeneous effects (**Supplementary Table 11**). Collectively, these results indicate that the vast majority of putative causal associations are consistent with the common pathway indexed by the latent EXT factor.

We further evaluated the robustness of fine-mapped signals via quasi-replication analyses in the independent All of Us Research Program^29,30^ (**Supplementary Tables 12–13**; **Methods**). We estimated externalizing factor scores from clinical phenotypes in electronic health record data (e.g., diagnoses of ADHD, CD, SUDs, and ASPD), separately in EUR-like and AFR-like samples (*N*_EUR_ = 201,904; *N*_AFR_ = 61,974). We then conducted ancestry-specific GWASs and meta-analyzed results across ancestries. Fine-mapped SNPs (PIP ≥ 0.50) showed stronger associations than background SNPs in the All of Us multi-ancestry meta-analysis (*P* = 2.58e-55). Moreover, 547/634 (86%, *P* = 1.96e-82) fine-mapped SNPs showed concordant effect direction and 212/634 (33%, *P* = 1.83e-84) had nominally significant associations (*α* = 0.05). These results provide clear evidence that the fine-mapped causal variants exhibit robust, replicable signals in a large independent GWAS dataset with clinically diagnosed phenotypes.

Finally, we used FLAMES^31^ to identify the most likely effector genes that mediate locus-trait associations (**Methods**). FLAMES identified 961 unique effector genes across the 1,294 genomic regions with cumulative precision ≥ 0.75, including 242 genes prioritized with very high confidence (cumulative precision ≥ 0.999). The prioritized genes included several with established relevance for self-regulated behavior (e.g., *CADM2*, *CHRM2*, *GABRA2*, *BDNF*, *SEMA6D*). Cross-referencing the prioritized gene list against the GWAS Catalog (**Supplementary Information**) reveals that 207 effector genes, including 28 mapped with very high confidence (**Supplementary Table 24**), represent novel associations not previously linked to any externalizing phenotype (e.g., *SLC17A6*, *KCNA4*, *NRN1, LRRTM3*).

### Contextualizing the biology of externalizing liability

We used MAGMA^32^ to compute gene-based association statistics within each ancestry group and meta-analyze results across populations (**Supplementary Table 14**). We then conducted MAGMA gene property and gene-set enrichment analyses to test whether polygenic signal for externalizing is associated with gene expression concentrated in particular tissues, developmental epochs, cell types, or biological pathways (**Supplementary Tables 15–23**).

Across 54 tissue-specific gene expression profiles, liability for externalizing was broadly enriched throughout the brain (*P*s ≤ 3.18e-7) (**Supplementary Table 15**). Similarly, analysis of gene expression profiles across 11 neurodevelopmental windows found significant enrichment across all periods (*P*s ≤ 5.76e-5). However, after controlling for average expression across windows, enrichment was only observed during early prenatal (*P* = 1.85e-5), early mid-prenatal (*P* = 1.20e-7), and late mid-prenatal periods (*P* = 2.15e-7) (**Fig. 3a**; **Supplementary Table 16**).

**Fig. 3.**
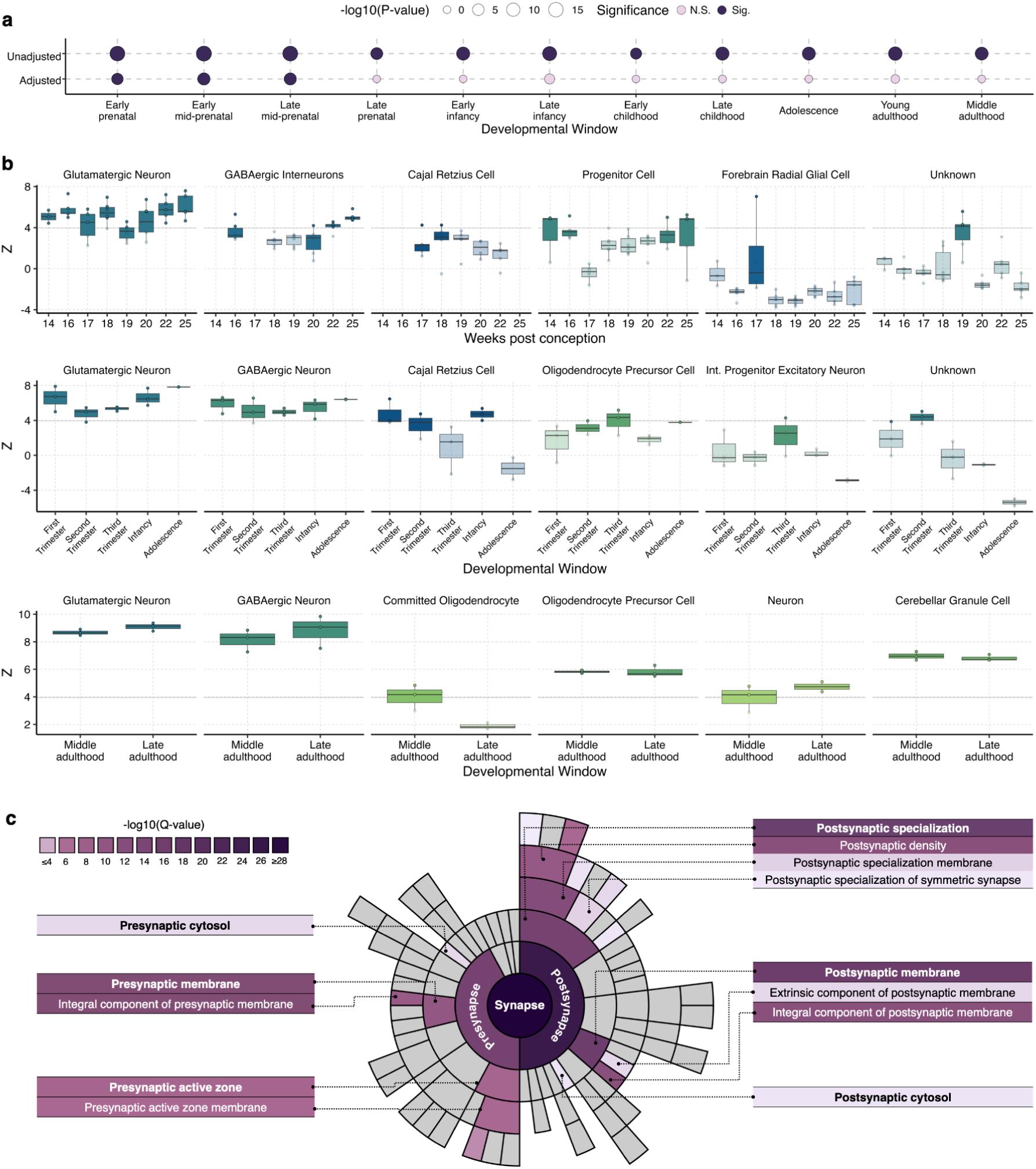
Bioinformatic characterization of externalizing liability. **a**, Gene property results depicting enrichment of polygenic signal for externalizing across developmental epochs in the Brainspan dataset, unadjusted and adjusted for average expression. The size of each circle corresponds to the magnitude of the enrichment statistic and color denotes statistical significance (**Supplementary Table 16**). **b**, Select gene property results depicting enrichment across specific cell types measured in three datasets: (i) mid-to-late prenatal (top), (ii) first trimester to adolescence (middle), and (iii) middle and late adulthood (bottom). Cellular granularity levels were selected to facilitate comparisons across datasets. Full results are provided in **Supplementary Tables 19–20, 22**. **c**, Sunburst plot from SynGO depicting the synaptic cellular components linked to the 961 effector genes prioritized via pathway-naïve FLAMES, applying a high-stringency evidence filter in SynGO. Colored segments represent cellular component terms that were significantly enriched at 1% FDR, while grey segments denote terms that were non-significant. The top-level “synapse” term is represented as the full circle at the center, with successive outer layers representing increasingly specific subcomponents (**Supplementary Table 25**). See **Supplementary Information** for further description of data.

We then interrogated the relevance of specific brain cell types across development, leveraging six human brain cell-resolved transcriptomic datasets spanning early prenatal development through older adulthood (**Methods**; **Supplementary Information**). Across two complementary early-gestation datasets (5.5–15 weeks post-conception, [wpc]), we observed enrichment in neurons and neuroblasts (**Supplementary Fig. 5**; **Supplementary Table 17**), with the strongest signals in GABAergic cell lineages (*P*s ≤ 1.45e-16) and additional signals in dopaminergic (*P*s ≤ 2.85e-8) and serotonergic cells (*P* = 2.89e-9) (**Supplementary Table 18**). Between 14–25 wpc, enrichment was sustained for glutamatergic neurons across the entire interval, whereas enrichment in GABAergic interneurons—local inhibitory neurons that regulate excitatory circuit activity—emerged later, beginning at 22 wpc (**Fig. 3b**; **Supplementary Fig. 6**; **Supplementary Table 19**).

When taking a broader assay of neocortical development spanning the first trimester through adolescence^33^, we found robust patterns of enrichment for glutamatergic and GABAergic neurons throughout development, with complementary signals in Cajal-Retzius cells during the first trimester and infancy (**Figure 3b**; **Supplementary Figure 7**; **Supplementary Table 20**). In adulthood^34,35^, signal was enriched in glutamatergic and GABAergic neurons and oligodendrocyte precursor cells broadly distributed throughout the brain, both in gray and white matter (**Fig. 3b**; **Supplementary Figs. 8–9**; **Supplementary Tables 21–22**).

To complement the cell-type analyses, we performed gene-set enrichment analyses to test whether the polygenic signal for externalizing is specific to certain biological processes, cellular components, and molecular functions. Briefly, results implicated processes governing neuronal differentiation and projection and involved in synaptic assembly and organization. Enrichment was also pronounced for postsynaptic architecture and specializations (**Supplementary Fig. 10**; **Supplementary Table 23**), particularly the postsynaptic (*P* = 9.13e-10) and somatodendritic compartments (*P* = 5.75e-10), including the dendritic tree (*P* = 1.03e-8) and neuron spine (*P* = 4.46e-6). Prioritized effector genes implicated similar pathways. Using pathway-naïve FLAMES and SynGO^36^ (**Methods**), we found that effector genes were enriched across both presynaptic (*P* = 2.72e-16) and postsynaptic (*P* = 3.01e-27) annotations, with strong enrichment in the postsynaptic membrane and postsynaptic density, as well as pathways regulating postsynapse assembly, receptor levels, and localization (*Ps* ≤ 6.47e-4) (**Fig. 3c**; **Supplementary Table 25**).

Overall, bioinformatic results converge on a biological profile in which externalizing risk is rooted in broadly distributed neural processes that begin in prenatal neurodevelopment. This systems-level architecture, which shows limited regional specificity, predominantly features the early and sustained involvement of GABAergic and glutamatergic neurons. At the level of the synapse, signals converge on the machinery of assembly, organization, and specialization, most prominently at the postsynaptic interface.

### Prioritizing candidate compounds for drug repurposing

We used DRUGSETS^37^, which leverages multimodal information about genes coding for drug targets and interacting proteins, to identify both categories and specific compounds that are acting on disease-relevant processes, some of which may be candidates for drug repurposing (**Supplementary Tables 26–27**). As DRUGSETS does not consider the direction of pharmacological effect, compounds with opposing effects on the same target (e.g., agonists and antagonists of the same receptor) may both be identified as significant, reflecting their action on the same biological process.

An evident inhibitory-synapse theme emerged. When grouping compounds by their mechanism of action, we found significant enrichment in GABA-receptor antagonists (*P* = 5.82e-12), with complementary signals for general anesthetics (N01A; *P* = 1.71e-8) and antiepileptics (N03A; *P* = 2.38e-7) based on Anatomical Therapeutic Chemical (ATC) classifications. Results based on clinical indications aligned with this pattern, with seizures (*P* = 8.25e-5) and anesthesia (*P* = 1.37e-4) showing significant enrichment. At the level of individual compounds, we observed drug-level enrichment for barbiturates (butalbital, metharbital, primidone, thiopental) and anesthetics (etomidate, methoxyflurane, enflurane, isoflurane, propofol, halothane, sevoflurane). Results also implicated the benzodiazepine-site antagonist flumazenil, as well as etifoxine, a non-benzodiazepine modulator with dual actions on GABA_A_ β2 and β3 subunits. Collectively, these signals situate GABA_A_ biology as a recurrent locus of effect.

Two additional themes concerned dopaminergic signaling and the balance of synaptic excitation and inhibition. Grouping by mechanism of action, polygenic signal was significantly enriched for dopamine-receptor antagonists (*P* = 8.77e-21), with complementary enrichment in antipsychotics (N05A; *P* = 7.07e-13) and drugs used to treat schizophrenia (*P* = 2.71e-12) and psychosis (*P* = 1.94e-7) in the ATC and clinical indication groupings, respectively. Representative compounds included paliperidone, haloperidol, perospirone, blonanserin, cariprazine, lurasidone, fluphenazine decanoate, and perphenazine. To a lesser degree, dopamine receptor agonists (mechanism of action; *P* = 9.02e-8) and dopaminergic agents (ATC, N04B; *P* = 1.47e-5) showed enrichment, including rasagiline, safinamide, talipexole, levodopa, rotigotine, and apomorphine. Polygenic signal was also enriched for calcium-channel blockers (*P* = 4.75e-5) and glutamate-receptor antagonists (*P* = 2.90e-4), including acamprosate, topiramate, and gabapentin, implicating the modulation of glutamatergic tone and reinforcement of inhibitory transmission. These patterns are congruent with the broader biological profile of externalizing and align with the clinical literature on modulating glutamate in the treatment of alcohol use disorder (AUD)^38–40^, documented effects of antipsychotics on aggressive behavior^41^, and the propensity of dopamine agonists to precipitate impulse-control disorders^42,43^.

A final set of signals implicated neurosteroid and sex-hormone pathways. Specifically, we found that progesterone-receptor agonists were enriched within mechanism-of-action groupings (*P* = 2.74e-5), with significant drug-level enrichments for dehydroepiandrosterone, dehydroepiandrosterone sulfate, estriol, fosfestrol, and lasofoxifene. These enrichment patterns are noteworthy given the role of neurosteroids and sex steroids as modulators of synaptic physiology^44^.

Collectively, the category- and compound-level enrichments situate a considerable component of externalizing liability in postsynaptic receptor complexes—most prominently GABA_A_ and dopaminergic synapses—and outline clinically relevant mechanisms for future pharmacogenomic consideration.

### Characterizing polygenic index associations

We calculated a genome-wide polygenic index (PGI) for EXT in EUR-like and AFR-like individuals from five independent cohorts (**Methods**; **Supplementary Information**). Specifically, we used PRS-CSx^45^ to calculate ancestry-specific and multi-ancestry versions of the EXT PGI. Here, we report results from the multi-ancestry PGI, which maximizes predictive performance and generalizability. We estimated both population and within-family “direct” effects of the EXT PGI on outcomes conceptually linked to externalizing liability. Results are summarized below (**Fig. 4a–c**), with cohort-specific details and sensitivity analyses reported in the supplementary materials (**Supplementary Tables 28–35; Supplementary Information).**

**Fig. 4.**
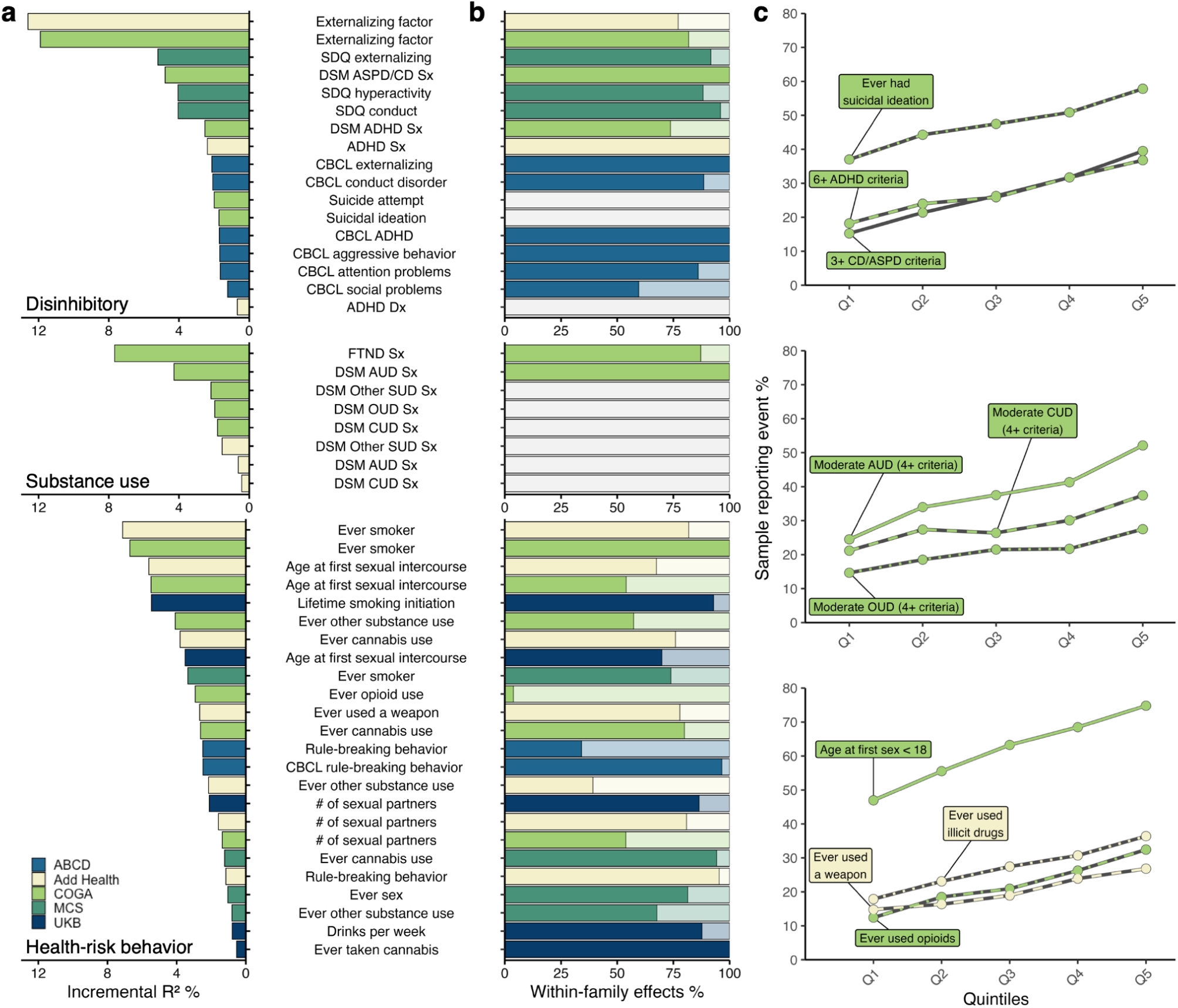
Associations between externalizing liability and clinically relevant disorders, symptoms, and health-risk behaviors in EUR-like participants. **a**, Incremental proportion of variance explained (*R*^2^) by the multi-ancestry EXT PGI across disinhibitory (top panel), substance use (middle panel), and health-risk behavior (bottom panel) phenotypes in the population analyses (**Supplementary Tables 29–33**). Different colors indicate different samples. All presented associations were statistically significant. **b**, Proportion of PGI effects due to direct (dark) versus indirect (light) genetic effects from the within-family models (**Supplementary Tables 29–33**). Associations that were not significant in the family-based subsamples are colored in grey. **c**, Relative risk across quintiles of the EXT PGI for 10 illustrative outcomes in COGA and Add Health (**Supplementary Table 35**). Note: Dx = diagnosis; Sx = symptoms.

In the Collaborative Study on the Genetics of Alcoholism (COGA) and the National Longitudinal Study of Adolescent to Adult Health (Add Health), we estimated a phenotypic latent factor of externalizing from seven indicators to parallel our Genomic SEM analysis (**Supplementary Table 28**). Among EUR-like individuals in both samples, the EXT PGI explained approximately 12% of the variance in the latent phenotypic externalizing factor (COGA: *β* = 0.352, s.e. = 0.016, incremental *R*^2^ = 11.9%; Add Health: *β* = 0.359, s.e. = 0.016, incremental *R*^2^ = 12.6%; **Supplementary Tables 29–30**), an effect size comparable to many well-established environmental correlates, including socioeconomic status^46^, child maltreatment^47^, and peer victimization^48^. Moreover, the EXT PGI showed significant associations with symptoms of SUDs (AUD, CUD, OUD, nicotine dependence [ND]) and with symptoms of other clinical disorders of childhood (ADHD and CD) and adulthood (ASPD) (incremental *R*^2^ = 0.4%–7.6%; **Fig. 4a**).

In EUR-like youths from the Adolescent Brain Cognitive Development (ABCD) and Millennium Cohort Study (MCS) cohorts (**Supplementary Tables 31–32**), the EXT PGI was associated with ADHD and CD symptoms, as well as measures of aggressive behavior (incremental *R*^2^ = 1.2%–5.2%; **Fig. 4a**). Across all datasets, the EXT PGI had significant associations in EUR-like individuals with health-risk behaviors, including smoking, cannabis use, opioid use, earlier age at first sex, and a greater number of sexual partners. Within-family models indicated that most of the associations largely reflected direct genetic effects, with a median direct genetic effect proportion of 79% (**Fig. 4b**; **Supplementary Tables 29–33**).

Among EUR-like individuals in Add Health, the EXT PGI also accounted for a significant proportion of variance in negative social outcomes, including school suspension, criminal justice system involvement, lower educational attainment, lower income, lack of health insurance, housing insecurity, and worse overall health. In this domain, population associations typically accounted for ∼1% to 6% of the variance (**Fig. 4a**). Notably, PGI associations with these social outcomes showed greater attenuations when estimated within-families (median proportion of direct genetic effects = 60%; **Fig. 4b**; **Supplementary Table 30**).

Among AFR-like individuals in COGA and Add Health, the EXT PGI was significantly associated with the phenotypic externalizing factor, albeit to a much-attenuated degree (COGA: *β* = 0.192, s.e. = 0.027, incremental *R*^2^ = 2.6%; Add Health: *β* = 0.207, s.e. = 0.029, incremental *R*^2^ = 3.4%). Broadly, results in AFR-like individuals followed a pattern of attenuated generalizability, with 10/30 (33%) phenotypes in COGA and 19/39 (49%) phenotypes in Add Health exhibiting statistically significant, yet considerably weaker, associations (*R*^2^ ≤ 3.4%). Among substance use phenotypes, the largest effects were observed for AUD symptoms, smoking initiation, and cannabis use. For disinhibited behavior phenotypes, the strongest associations included ASPD/CD symptoms, rule-breaking behavior, and ever using a weapon; for negative social outcomes, educational attainment and criminal justice system involvement showed the largest effects (**Supplementary Tables 29–30**).

We next investigated the medical correlates of the EXT PGI by conducting a phenome-wide association study (PheWAS) in the Vanderbilt University Medical Center Biobank (BioVU), analyzing up to 66,184 EUR-like and 12,250 AFR-like individuals with linked genotypes and International Classification of Diseases (ICD)-9/10–derived phecodes (**Methods**; **Supplementary Information**). In EUR-like participants, the EXT PGI was associated with 16% (223/1,440) of phecodes at Bonferroni-corrected significance (**Fig. 5a**; **Supplementary Table 36**), with odds ratios (ORs) ranging from 0.75 to 1.56 per standard deviation increase in the PGI. The largest effects were observed for substance-related diagnoses (TUD: OR = 1.51; SUDs: OR = 1.42; AUD: OR = 1.35) and their medical sequelae (viral hepatitis C: OR = 1.56; obstructive chronic bronchitis: OR = 1.40; alcoholic liver damage: OR = 1.35; emphysema: OR = 1.33; chronic airway obstruction: OR = 1.31; cancer of the larynx: OR = 1.30; abdominal aortic aneurysm: OR = 1.25). The EXT PGI was also associated with increased odds of ASPD/BPD (OR = 1.47), posttraumatic stress disorder (OR = 1.39), bipolar disorder (OR = 1.28), CD (OR = 1.28), and ADHD (OR = 1.22). Finally, we observed robust associations with outcomes linked to risky sexual behavior (HIV infection: OR = 1.47; other sexually transmitted infections: OR = 1.32), mental disorders during/after pregnancy (OR = 1.32), and suicidality (suicide or self-inflicted injury: OR = 1.43; suicidal ideation or attempt: OR = 1.35). In AFR-like participants, only three phecodes were significant at a Bonferroni-corrected threshold: SUDs (OR = 1.42), TUD (OR = 1.38), and mood disorders (OR = 1.15).

**Fig. 5.**
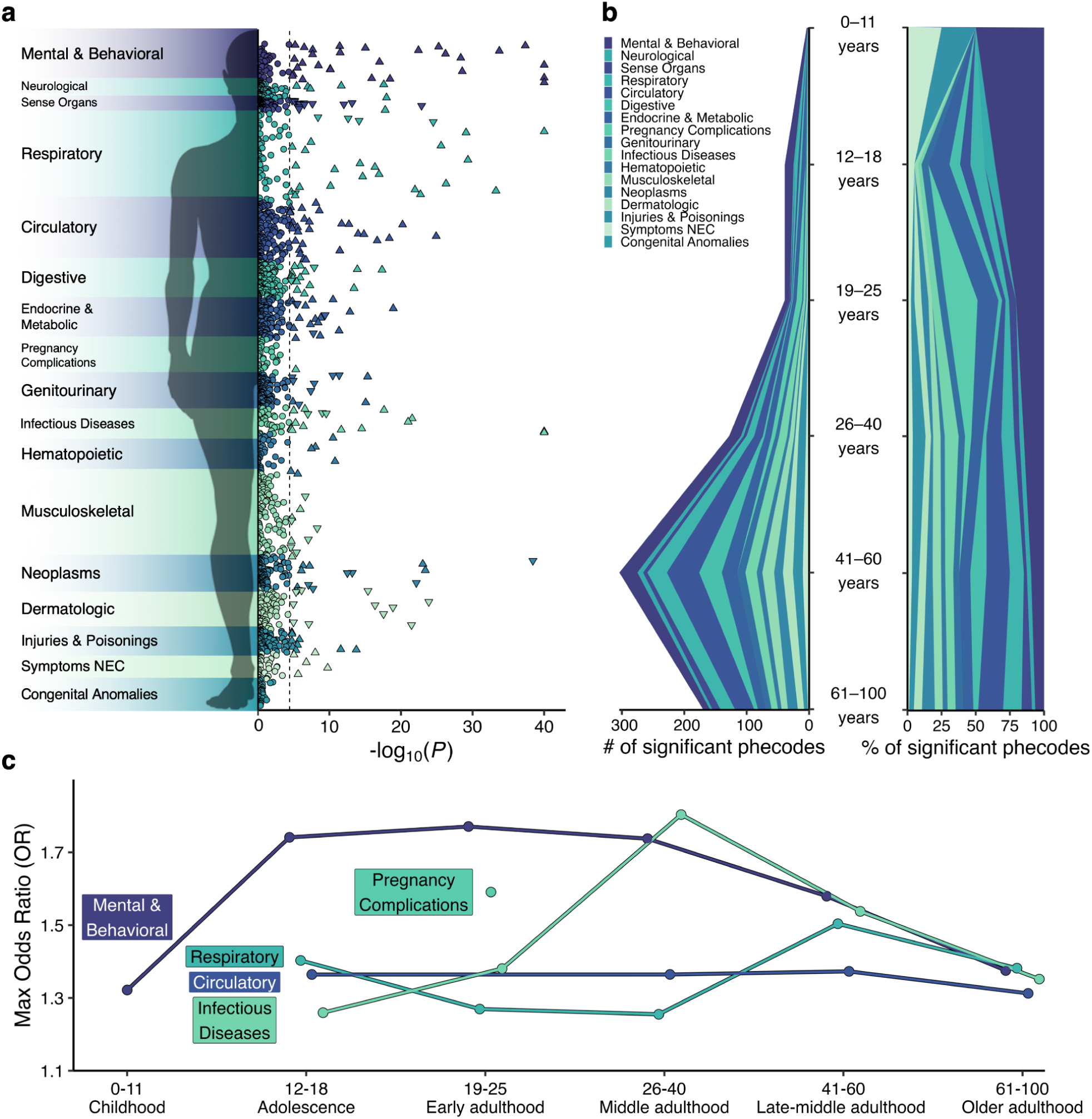
Associations between externalizing liability and medical outcomes. **a**, Manhattan plot displaying −log10 *P* values from a two-sided *Z* test of the association between the EXT PGI and 1,440 medical outcomes in up to 66,184 participants of EUR-like ancestry (**Supplementary Table 36**). The dashed line represents the Bonferroni-corrected significance threshold accounting for the number of tested medical conditions. Triangles indicate Bonferroni-corrected significant associations, and circles indicate non-significant associations. The direction of the triangle refers to the direction of effect. NEC = not elsewhere classified. **b**, Distribution of PGI-phecode associations across the lifespan, with a raw density plot on the left and a conditional density plot on the right (**Supplementary Table 37**). **c**, Plot of the maximum odds ratios in each age strata for select phecode categories with at least one significant association (**Supplementary Table 37**).

We then conducted age-stratified PheWAS analyses within EUR-like individuals to estimate associations specific to childhood (ages 0–11; *n* = 8,443), adolescence (ages 12–18; *n* = 6,893), early adulthood (ages 19–25; *n* = 7,211), middle adulthood (ages 26–40; *n* = 15,437), late-middle adulthood (ages 41–60; *n* = 31,346), and older adulthood (ages 61–100; *n* = 26,819) (**Fig. 5b,c**; **Supplementary Table 37**). In childhood and adolescence, a higher EXT PGI was primarily associated with increased odds of psychiatric conditions, including ADHD, CD, and suicidality. In early adulthood, 9 of the 39 significant associations were related to pregnancy complications (e.g., early or threatened labor)—potential consequences of substance use and infection during pregnancy. By middle and late adulthood, the EXT PGI was strongly associated with SUDs, personality disorders, and suicidality, as well as infectious, circulatory, and respiratory diseases. This health burden persisted across the lifespan into older adulthood (**Fig. 5c**).

## Discussion

Externalizing spectrum disorders—including ADHD and CD in childhood and SUDs and ASPD in adulthood—are highly comorbid, impairing, and costly psychiatric conditions^1–8,49,50^. While genomic studies have the potential to illuminate the etiology of these disorders and to accelerate the development of efficacious treatments^16^, amassing sufficient data on any one disorder has been challenging. Here, we overcome that challenge by jointly analyzing genetic influences on multiple externalizing phenotypes, leveraging their polygenic overlap to significantly advance gene discovery. We modeled an updated externalizing genomic factor that represents a cross-cutting dimension of liability to harmful disinhibition, spanning relatively normative health-risk behaviors to clinically significant psychiatric diagnoses and symptoms.

Critically, our study characterizes this externalizing liability across ancestral populations. By jointly analyzing data from groups with EUR- and AFR-like ancestries, we demonstrate that the common variant architecture of externalizing is highly similar across populations, and that its etiological relationships with other traits largely align. Although predictive performance of the PGI is attenuated in cohorts with AFR-like ancestry, underscoring the need for larger discovery samples with non-EUR-like ancestries^51^, the shared architecture nevertheless provides essential context for generalizability: The biological pathways and candidate therapeutics identified in one population are likely relevant in others. Moreover, this concordance motivates multi-ancestry genomic discovery efforts, as undertaken here.

Our multivariate, multi-ancestry approach to genetic discovery is substantially more powerful than any GWAS of a single externalizing disorder. The results of our meta-analysis identified nearly 1,300 associated genomic loci, more than double the number of loci identified in our previous multivariate GWAS of externalizing—and approximately 14× more loci than previously identified for AUD alone^22^, 15× more than TUD alone^52^, 43× more than CUD alone^23^, 48× more than ADHD alone^21^, and 72× more than identified for OUD alone^53^. Fine-mapping implicated more than 600 putative causal variants with cross-cutting effects, and integrative gene prioritization identified more than 950 effector genes, many at high confidence and novel to any given externalizing spectrum disorder. Critically, these findings exhibited replicable signals in a large independent dataset with clinically diagnosed externalizing disorders.

With this improved statistical power, our results sharpen our understanding of the etiology of externalizing psychopathology. First, the biology is developmentally situated, with genetic signal concentrating in early- to mid-prenatal neurodevelopment, when cortical lineages diversify, projection neurons extend long-range axons, and inhibitory programs differentiate and migrate^54,55^. Within this period, glutamatergic and GABAergic neuronal lineages show enrichment as early as 5–15 weeks post-conception. The role of GABAergic interneurons, specifically, becomes more prominent at ∼22 weeks, when these cells are completing their tangential migration and integrating into the cortical plate^56–58^, forming the initial wiring of local inhibitory circuits^58^, and acting as early excitatory drivers^59,60^. These signals are observed across multiple cell-resolved transcriptomic datasets, persisting throughout neurodevelopment with limited spatial specificity. Together, these findings situate liability for externalizing spectrum disorders within early neurodevelopment programs that calibrate excitatory-inhibitory relationships.

The results of our gene-set enrichment analyses further implicate the synapse as a specific subcellular locus. Both genome-wide and prioritized-gene bioinformatic results converge on synaptic biology with a marked emphasis on postsynaptic assembly, organization, and specialization. Enrichment spans canonical postsynaptic compartments and dendritic spine specialization, as well as processes that position and stabilize receptors and the synaptic face (e.g., regulation of postsynaptic levels and localization). One parsimonious interpretation of these patterns is that common genetic variation may tune the receptor-scaffold interface and modulate synaptic gain and plasticity^61–63^, though further study is required to test this hypothesis.

Drug repurposing results provide mechanistic clues and point to testable avenues for treatment development for the externalizing spectrum, where pharmacotherapies remain scarce. The clearest pattern involved the over-representation of pharmacological compounds that act on GABA_A_ receptor systems, with positive allosteric modulator compounds exhibiting particularly pronounced signals. Dopamine-receptor antagonists also showed robust enrichment, aligning with preliminary clinical evidence that some modern antipsychotics with favorable receptor profiles may be tools for the short-term management of aggressive behavior in children and adolescents^64^. Signals for calcium-channel blockers and glutamate-receptor antagonists further suggest that excitatory-inhibitory balance plays a key role in the pathophysiology of externalizing spectrum disorders. Finally, enrichment in neurosteroid/sex-hormone compounds highlights another developmentally plausible pathway. Importantly, these results only indicate the etiological involvement of pathways, not whether any given compound would ameliorate or exacerbate symptomatology. Accordingly, these drug classes and compounds should be viewed as a prioritized list of modifiable pathways that can guide *in silico* and, ultimately, *in vivo* translational efforts.

Polygenic index analyses indicate the utility of genetic data for epidemiological and clinical research. An EXT PGI explained ∼12% of the variance in a phenotypic externalizing factor in two independent samples of adults, an effect size that is commensurate with recognized environmental risk factors (e.g., childhood maltreatment, peer victimization, parental socioeconomic status)^47,48,46^. Predictive performance was attenuated in AFR-like cohorts, consistent with cross-ancestry portability constraints^65,66^, but the direction and pattern of associations were preserved. We also found that the EXT PGI was reliably and robustly associated with multiple clinically relevant phenotypes, including symptoms of DSM-defined psychiatric disorders, health-risk behaviors, adverse social outcomes, and multiple diseases affecting all bodily systems. Critically, results from within-family models indicate that most genetic associations between the EXT PGI and psychiatric phenotypes predominantly reflect direct genetic effects rather than uncorrected population stratification, indirect genetic effects, or assortative mating. We emphasize that genotypes are not deterministically related to individual outcomes, and that the EXT PGI is most useful for research when combined with information about other relevant biopsychosocial factors.

Our study has limitations. Despite including individuals of AFR-like ancestry, statistical power in these cohorts remains limited, and individuals with other genomic backgrounds are not included here. Sex-pooled analyses of the autosomes do not shed light on potential genetic contributions to the well-established sex difference in the prevalence and presentation of externalizing spectrum disorders. Relatedly, our focus on common genetic variants necessarily omits the effects of rare variants that have been found to contribute to risk for specific externalizing spectrum disorders like ADHD^67^. Moreover, the phenotypic variance in an externalizing factor explained by our PGI, while consistent with or exceeding the effect sizes commonly seen for measured environmental risk factors, is still far lower than the heritability estimated in twin models, and the likely reasons for “missing” or “hidden” heritability remain contested. Finally, the genetic architecture of each externalizing spectrum disorder reflects a mixture of both shared and disorder-specific liability. Our multivariate approach is a powerful strategy for identifying the shared genetic etiology; this strategy can and should be extended to also study the disorder-specific effects, so that we can delineate the myriad of genetically influenced pathways that contribute to each disorder.

The present study significantly advances our etiological understanding of a highly stigmatized and insufficiently understood set of conditions and behaviors. We delineate a shared, developmentally rooted genetic liability that operates through broadly distributed neural systems, implicates postsynaptic biology, and generates novel pharmacogenomic hypotheses with plausible mechanistic grounding. We demonstrate that individuals with elevated genetic liability for externalizing spectrum disorders are at increased risk for multiple psychiatric, social, and health burdens from early life to adulthood, underscoring the urgent need for more effective psychosocial and pharmacological interventions that target this transdiagnostic liability. By integrating multivariate, multi-ancestry genomic discovery with thorough downstream biological contextualization and characterization, this study establishes a foundation for targeted mechanistic work and translation geared at mitigating disinhibition and its sequelae.

## Methods

This manuscript is accompanied by **Supplementary Information**, which includes additional tables, figures, and methodological details. This study followed a preregistered analysis plan (https://osf.io/7pfgj/) that specified procedures for discovery stage analyses.

### GWAS data acquisition and preprocessing

We obtained univariate GWAS summary statistics for externalizing phenotypes from public and private repositories, including 23andMe Research Institute; the GWAS & Sequencing Consortium of Alcohol and Nicotine Use (GSCAN) Consortium; the Integrative Psychiatric Research Consortium (iPSYCH) study; the Million Veterans Program (MVP); the PsycheMERGE (electronic MEdical Record and GEnomics) Network; and the Psychiatric Genomics Consortium (PGC). Several GWAS summary statistics from our prior multivariate GWAS of the externalizing spectrum^18^ were also used. The univariate GWASs considered for analysis, and the constituent cohorts that contributed to those GWASs, are described in **Supplementary Tables 1–3**.

Across the seven discovery phenotypes included in the EUR-like model, the combined data included a total of 5,726,594 observations. After conservatively assuming full sample overlap when a cohort contributed to multiple indicator GWASs, we found that at least 3,786,374 unique individuals contributed to these GWASs (**Supplementary Tables 1–2**). For the AFR-like analyses, the indicator GWASs included a total of 362,830 observations from at least 243,163 unique individuals. Together, our discovery-stage analyses were based on 6,089,424 observations measured in at least 4,029,537 unique individuals across EUR-like and AFR-like ancestry groups.

All univariate GWASs were subjected to a standardized quality control and harmonization protocol prior to multivariate modeling (**Supplementary Information**). Specifically, summary statistics were processed using EasyQC^68^, following the procedures employed in our previous externalizing GWAS^18^. These procedures ensured that discovery inputs were harmonized across data sources, restricted to high-quality SNPs, and ancestrally matched to appropriate linkage disequilibrium (LD) reference panels. We also conducted sensitivity analyses to confirm that the inclusion of data from 23andMe did not bias our results (**Supplementary Information**).

### Genomic structural equation modeling

We used Genomic SEM^19^ v0.0.5 to: (i) fit a common factor model to an updated set of externalizing-related GWASs, (ii) evaluate genetic relationships between externalizing and other complex traits, (iii) estimate the effects of individual single-nucleotide polymorphisms (SNPs) on a latent externalizing factor, and (iv) characterize heterogeneous genetic effects across externalizing phenotypes. These analyses were conducted separately in EUR-like and AFR-like populations, using ancestry-matched reference panels and following the best practices outlined by the Genomic SEM developers. The specific procedures and parameters relevant to each analysis are described below.

### Genomic confirmatory factor analysis

The first set of Genomic SEM analyses was a series of confirmatory factor models informed by our prior work^18^. Ancestry-specific GWAS summary statistics were processed with the *munge* function, retaining only HapMap3 SNPs outside of the major histocompatibility complex regions with an imputation quality score ≥ 0.90 (when available). Alleles with a minor allele frequency < 0.01 were excluded from analysis for EUR-like data, while alleles with a frequency < 0.05 were excluded from analysis for AFR-like data. The more conservative threshold in AFR-like data was selected to ameliorate concerns about potential mismatch between the reference panel and GWAS source data. For case-control indicator GWASs, we applied a liability-scale adjustment with the ‘sum of *N*_eff_’ approach^25^. Multivariable LD score regression was used to estimate the genetic covariance matrix and its sampling covariance matrix for downstream modeling. Model selection and evaluation followed preregistered criteria based on the standard fit indices (χ^2^ as a comparative statistic, CFI, SRMR), interpreted as recommended for large-sample GWAS applications.

### Genetic correlations with other complex traits

Genomic SEM was used to estimate genetic correlations between the EXT factor and other complex traits in both EUR-like and AFR-like ancestries (**Supplementary Information**). GWAS summary statistics were processed with the *munge* function of Genomic SEM, using the same approach described above. All genetic correlations with the common factor were also subjected to *Q*_Trait_ tests^69,70^, which quantify the degree to which genetic relationships between model indicators and an exogenous trait plausibly operate via the implied common pathway. When *Q*_Trait_ was significant, we fit follow-up models that iteratively freed direct paths until either heterogeneity was no longer detectable or the model was saturated. In the presence of heterogeneity, genetic correlations were taken from the final adjusted model. A Bonferroni correction was used to adjust for multiple comparisons within each ancestry separately (EUR-like *P* ≤ 0.05/95 = 5.26e-4, AFR-like *P* ≤ 0.05/14 = 3.57e-3).

To assess how similar EXT’s associations with other complex traits were across ancestries, we calculated the correlation between the genetic correlation estimates in EUR-like and AFR-like data. Given the uncertainty of those estimates—especially in the AFR-like data—we used an errors-in-variables approach to calculate a disattenuated correlation coefficient (**Supplementary Information**). This correlation coefficient is corrected for attenuation due to measurement error, defined here as the sampling variability of the genetic correlation estimates. Specifically, we computed the disattenuated correlation by taking the cross-trait covariance of the observed pairs and dividing by the product of the error-corrected standard deviations in each ancestry, obtained by subtracting the average squared standard error from the corresponding across-trait variance. Confidence intervals were obtained by a nonparametric bootstrap over traits.

### Multivariate GWAS of externalizing

For each ancestry, Genomic SEM was used to estimate SNP effects on EXT and to evaluate SNP-level heterogeneity. Prior to analysis, the *sumstats* function was used to align effect alleles across indicators and to standardize effect estimates and standard errors. To maximize coverage for discovery, multivariate GWAS inputs applied filters for MAF ≥ 0.01, yielding 6,331,678 SNPs for analysis for EUR-like data and 7,115,963 SNPs for analysis for AFR-like data. Models used diagonally weighted least squares (DWLS) with ancestry-matched LD scores from 1000 Genomes Phase 3 v5^71^. The estimated sample size of the factor (*N^*) was computed using the approach described by Mallard and colleagues^72^, restricting to SNPs with 0.10 ≤ MAF ≤ 0.40. *N^* was estimated as 3,943,075 and 303,641 for EXT_EUR_ and EXT_AFR_, respectively. Heterogeneity at the SNP level was assessed with the *Q*_SNP_ statistic^19^, which compares a model where the SNP loads on the factor (i.e., operating via a common pathway) versus a model allowing direct SNP effects on indicators (i.e., operating via independent pathways). We evaluated statistical significance for both the multivariate GWAS and *Q*_SNP_ at the conventional genome-wide threshold of *P* ≤ 5e-8.

### Multi-ancestry fine-mapping & gene prioritization

#### Fine-mapping putative causal variants

To identify the putative causal variants associated with externalizing, we first defined approximately independent regions harboring significant associations in the METAL meta-analysis results via LD-based clumping, performed separately in EUR-like and AFR-like results using ancestry-matched reference data from the 1000 Genomes Phase 3 v5 dataset. Specifically, we used PLINK^73^ v 1.9.0-b.7.7 to identify variants at a genome-wide significance threshold of *P* ≤ 5e-8, aggregating nearby signals with *P* ≤ 0.05 and LD *R*^2^ ≥ 0.10 within a 250-kb window, restricting to SNPs with a MAF ≥ 0.005. The boundaries of clumped loci were extended by ±100 kb to capture regulatory elements proximal to the index SNPs. We generated sets of clumped loci for EUR-like and AFR-like results separately, then merged across ancestries to create a unified list for cross-ancestry fine-mapping and other downstream analyses.

Within each unified locus, we used SuSiEx^28^ v1.1.2 to perform cross-population fine-mapping to identify the putative causal variants that are associated with externalizing, integrating data across ancestries while modeling population-specific allele frequencies and LD patterns. This yielded posterior inclusion probability (PIP) estimates for each variant, as well as 95% credible sets. We prioritized causal variants based on PIP values, defining candidates at two confidence thresholds: PIP ≥ 0.50 for moderate confidence and PIP ≥ 0.95 for high confidence. We also required fine-mapped variants to exhibit a non-significant *Q*_SNP_ *P* value in order to be selected for replication (i.e., restricting to candidates that plausibly act via the common pathway).

#### Effector gene prioritization

We used FLAMES^31^, a novel framework for integrating multimodal evidence to predict the most likely effector gene within a locus, to identify the specific genes underlying externalizing liability. FLAMES requires three inputs: PIP values from a fine-mapping analysis (here, SuSiEx), gene-based test statistics from MAGMA, and polygenic prioritization scores from PoPS. All procedures are summarized below (and further described in the **Supplementary Information**).

First, we used SuSiEx to calculate PIP values for SNPs within each credible set, as described above. Next, we used MAGMA v1.10 to conduct gene-based association tests, mapping SNP-based signals to protein coding genes within and across ancestry groups. Consistent with best practices, we used the SNP-wise mean model to map autosomal SNP-level signals to genes based on physical location, using gene boundaries defined by Ensembl build 110. Gene-based association analyses were conducted separately in EUR-like and AFR-like data with ancestry-matched LD reference data from the 1000 Genomes Phase 3 v5 dataset, yielding ancestry-specific gene-level *Z* statistics and gene-gene correlation matrices. To calculate multi-ancestry gene-based association results, we used MAGMA’s standard implementation of Stouffer’s method to meta-analyze signals across populations, allowing genes present in either population to enter the meta-estimate. We also used MAGMA’s default analytic procedures to generate a multi-ancestry gene-gene correlation matrix.

Finally, we used PoPS^74^ to calculate polygenic prioritization scores for each gene in the multi-ancestry MAGMA results. Briefly, PoPS uses gene-level associations to learn the joint polygenic enrichments of gene features derived from cell-type-specific gene expression, biological pathways, and protein-protein interactions, and then uses these enrichments to calculate a priority score for every protein-coding gene in the feature space. Here, we followed the suggestions of FLAMES developers, conducting PoPS analyses using default parameters and the default feature set. Note that we also performed this analysis excluding pathway-based features (i.e., pathway-naïve PoPS) to ensure that any downstream analysis of FLAMES prioritized genes would not be susceptible to false positives caused by non-independence of training and testing data^31^.

Following the recommendations of FLAMES developers, we then combined these inputs with GTEx-based tissue-specific weights in order to identify the most likely effector genes underlying externalizing. We also repeated gene prioritization using the pathway-naïve PoPS scores, as described above. All FLAMES analyses were employed using standard procedures and default settings.

#### Multi-ancestry meta-analysis

We used METAL^27^ (v2011-03-25) to perform a sample-size-weighted fixed-effect meta-analysis of multivariate GWAS results from EUR-like and AFR-like ancestry groups, including variants present in either group. Weights were proportional to the estimated sample size (*N^*). The multi-ancestry meta-analytic results were used to define independently associated regions across ancestry groups in the finemapping analyses and for data visualization purposes (e.g., producing a single cross-ancestry Manhattan plot) and the replication sign test.

#### Replication

Fine-mapped variants were selected from the SuSiEx results, yielding two sets after removing variants with a significant *Q*_SNP_ signal: a moderate-confidence set (PIP ≥ 0.50; *n*_SNPs_ = 637) and a high-confidence set (PIP ≥ 0.95; *n*_SNPs_ = 209). We evaluated the robustness of these fine-mapped signals using a quasi-replication design in the multi-ancestry context with an independent dataset (see **Supplementary Information** for full details). Specifically, we estimated a phenotype externalizing factor score using electronic health record data on externalizing spectrum disorders in 201,904 EUR-like and 61,974 AFR-like participants from the All of Us Research Program. We then conducted within-ancestry GWASs of these factor scores and meta-analyzed the results using METAL (following the same analytic procedure used for discovery). The multi-ancestry GWAS results were then used for quasi-replication analyses.

For each variant set, we constructed effective genotype variance-matched, LD-pruned background sets from the All of Us GWAS (*R*^2^ < 0.10; per-SNP *N* ≥ 80% of the maximum *N;* **Supplementary Information**). The background PIP ≥ 0.50 SNP set included 64,387 SNPs, and the background PIP ≥ 0.95 SNP set included 64,395 SNPs. Replication was then evaluated with three criteria: (i) joint enrichment via a one-sided Mann-Whitney (Wilcoxon rank-sum) test comparing the distribution of All of Us *P* values for fine-mapped SNPs versus background SNPs; (ii) sign concordance via a binomial test against a null of 0.50; and (iii) nominal and Bonferroni-corrected significance via binomial tests that compared the proportion of fine-mapped SNPs with *P* ≤ 0.05 and *P* ≤ 0.05/number of fine-mapped SNPs relative to expectations based on the matched background.

### Bio-annotation & bioinformatic analyses

To contextualize the multivariate GWAS signals associated with externalizing, we implemented a multi-ancestry bio-annotation and bioinformatics pipeline designed to characterize the relevant biology across tissues, developmental stages, cell types, and biological pathways via MAGMA^32^, DRUGSETS^37^, and SynGO^36^. All analyses were implemented in an ancestry-aware manner and then combined to guide multi-ancestry inference. Note that *P* value thresholds reported in the main text correspond to the results based on multiple ancestral populations; ancestry-specific thresholds, which may differ slightly, are reported in the **Supplementary Information**.

#### Bioinformatic characterization of genome-wide architecture

We used MAGMA^32^ v1.10 to conduct a series of gene property and gene-set enrichment analyses for the EUR-like, AFR-like, and multi-ancestry results. These analyses were based on the gene-based association statistics described above, using default MAGMA parameters and standard procedures. For gene property analyses, we used the GTEx^75^ v8 dataset to examine enrichment in 54 bodily tissues, the BrainSpan^76^ dataset to assess enrichment across 11 neurodevelopmental epochs, and 6 cell-resolved transcriptomic datasets to assess enrichment across brain-related cell types (see **Supplementary Information** for description of these data). For gene-set enrichment analyses, we tested 10,468 ontological gene sets^77^ from the Molecular Signatures Database (MSigDB)^78^ v2023.1, corresponding to 7,576 biological processes, 1,040 cellular components, and 1,852 molecular functions. Statistical significance was evaluated with Bonferroni-corrected thresholds (GTEx: *P* ≤ 0.05/54 = 9.26e-4; BrainSpan: *P* ≤ 0.05/11 = 4.54e-3; MSigDB: *P* ≤ 0.05/10,468 = 4.77e-6; cell-resolved transcriptomic: *P* value thresholds are dataset-specific, see **Supplementary Information**).

Finally, we applied an adjusted version of DRUGSETS^37^, which implements MAGMA’s competitive gene-set framework, to test whether medications whose protein targets (or well-curated interactors) are enriched among externalizing-associated genes. In this adjusted version, MAGMA *Z* statistics were replaced with PoPS scores, a gene-level metric that integrates GWAS summary statistics with functional genomic features to prioritize putative causal genes. Drug-gene interactions were drawn from DGIdb^79^ and the Drug Repurposing Hub^80^, yielding 1,201 compound-level drug-gene sets. For each drug, we ran competitive gene-set analyses on the multi-ancestry and ancestry-specific gene statistics described above, conditioning on the union of all drug-target genes to account for properties shared across pharmacological targets. Statistical significance was evaluated with Bonferroni thresholds reflecting the number of drug-gene sets tested in each analysis: 1,171 for multi-ancestry (*P* ≤ 0.05/1,171 = 4.23e-5), 1,163 for EUR-like (*P* ≤ 0.05/1,163 = 4.29e-5), and 1,169 for AFR-like (*P* ≤ 0.05/1,169 = 4.27e-5). To contextualize compound-level signals, we used DRUGSETS’ built-in grouping function to aggregate drug-level results by Anatomical Therapeutic Chemical (ATC) level III class, clinical indication, and mechanism of action. Within each grouping, a multiple linear regression model is fit with the drug-level gene-set *t* statistic as the outcome and indicators for group membership as predictors (restricting to groups with ≥ 5 compounds) and applied Bonferroni correction within grouping (ATC: *P* ≤ 5.56e-4; indication: *P* ≤ 4.24e-4; MOA: *P* ≤ 6.33e-4). All analyses were based on standard DRUGSETS procedures.

#### Bioinformatic characterization of prioritized genes

We used SynGO^36^ to characterize the synaptic biology linked to our set of prioritized genes. Specifically, we tested whether those genes were over-represented in synaptic cellular component and biological process gene sets, using brain-expressed genes as the background set and applying a high stringency evidence filter. For a term to be considered, we required that a minimum of three genes overlap with the annotation. Statistical significance was set as a Benjamini-Hochberg false discovery rate (FDR) of 1% (the default for SynGO).

### Polygenic index analyses

We conducted polygenic prediction analyses examining antecedents, correlates, and sequelae of externalizing psychopathology across six independent cohorts: the ABCD Study, COGA, Add Health, MCS, the UK Biobank sibling subsample (UKB-siblings), and BioVU. These samples were selected to provide comprehensive phenotypic coverage across the lifespan, as well as data on multiple ancestral groups and related individuals. The polygenic prediction analyses and cohorts are succinctly summarized below (see **Supplementary Information** for further details).

Polygenic indices (PGIs) were calculated in a harmonized manner across all study cohorts. Briefly, we used PRS-CSx to apply a continuous shrinkage prior to ancestry-specific SNP effect estimates and infer posterior SNP weights for common HapMap3 SNPs (MAF ≥ 0.01), using the 1000 Genomes Phase 3 v5 reference dataset to model ancestry-specific LD patterns. We generated three sets of weights: those for a multi-ancestry PGI (1,133,211 SNPs), a EUR-specific PGI (1,031,200 SNPs), and an AFR-specific PGI (552,102 SNPs). Within each cohort, PGIs were computed in PLINK as the sum of effect allele dosages weighted by posterior effects and were standardized to mean zero and unit variance within ancestry. All analyses were conducted in an ancestry-specific manner to minimize potential confounding by population stratification. We prioritize reporting of results based on the multi-ancestry PGI weights in the main text, with ancestry-specific results reported in the supplementary materials.

#### Polygenic prediction of externalizing outcomes

Polygenic prediction of clinical symptoms and disorders, health-risk behaviors, personality traits, and social and medical correlates was performed following a standardized protocol designed to quantify both the population and direct effects of the externalizing PGI (**Supplementary Information**). Briefly, for each cohort, associations between the externalizing PGI and outcomes were assessed with a three-step approach.

1. Population effect estimation: Regression models were fit to the full cohort sample to estimate the overall effect of the externalizing PGI on each outcome. Robust standard errors were used to account for relatedness among individuals.
2. Sibling subsample validation: Prior to quantifying the direct effects of the externalizing PGI, we first sought to evaluate whether observed polygenic associations were suitable for within-family analyses. To this end, significant associations from the previous step were tested within the sibling subsample of each cohort, again using robust standard errors. This allowed us to evaluate whether population effects were still observable despite lower statistical power due to reduced sample size and varied patterns of outcome endorsement.
3. Direct effect estimation: For outcomes with significant associations in the sibling subsample, within-family regression models were fit to estimate the direct effect of the externalizing PGI. Models included family fixed effects (family-level dummy variables) and robust standard errors.

Within each cohort and ancestry, models included a common covariate set (sex, age, and ≥10 genetic principal components, with cohort-specific adjustments as appropriate). All effects were estimated with ordinary least squares for both continuous and binary outcomes (i.e., a linear probability model; see **Supplementary Information** for further discussion of this approach).

#### Phenome-wide association analyses

To examine associations between externalizing liability and a broad range of medical outcomes, we conducted a phenome-wide association study (PheWAS) in BioVU (*n*_EUR_ = 66,184; *n*_AFR_ = 12,250). We used a standard “phecode” framework for these analyses, defining and measuring medical outcomes based on ICD-9 or ICD-10 diagnostic codes from structured electronic health record data. After quality control procedures, 1,440 phecodes were selected for analysis. The PheWAS R package (v0.99.5-2) was used to conduct phenome-wide association analyses. For each phecode, we fit a logistic regression of case status on the PGI, adjusting for sex, the median age from the longitudinal electronic health record data, and the first 10 genetic principal components. Phenome-wide significance was set at a Bonferroni-adjusted threshold of *P* ≤ 0.05/1,440 = 3.47e-5.

To characterize the developmental patterning of associations, we repeated the PheWAS in EUR-like BioVU participants within six age bins based on median age in the electronic health records: 0–11 (childhood; *n* = 7,042–8,443), 12–18 (adolescence; *n* = 5,886–6,893), 19–25 (young adulthood; *n* = 6,241–7,211), 26–40 (adulthood; *n* = 13,261–15,437), 41–60 (midlife; *n* = 26,269–31,346), and 61–100 (older adulthood; *n* = 22,405–26,819). As age was indexed at the code level, individuals could appear in more than one age bin if they had relevant ICD codes recorded at different ages. Age-stratified analyses were not conducted in AFR-like participants due to limited sample size. To formally test heterogeneity across age groups, we focused on phecodes that were FDR-significant in at least two bins and computed Cochran’s Q using the metafor v4.8-0 R package (**Supplementary Information**).

## Acknowledgements

This research was carried out under the auspices of the Externalizing Consortium. The Externalizing Consortium has been supported by the National Institute on Drug Abuse (R01DA050721 to D.M.D.) and the National Institute on Alcohol Abuse and Alcoholism (R01AA015416, administrative supplement to D.M.D.). Additional support for co-authors’ effort includes: E.C.J. (K01DA051759), H.E.P. (K01DA059657), J.D.D. (K01DA058807), S.J.B. (K23DA058808), S.S.R. (DP1DA054394; R01DA061977; T32IR5226), J.G. (1R01DA058862 and 5I01BX006482), M.C. (T32 DA55569), R.K.L. (Dutch Research Council [NWO] Veni project VI.Veni.221E.080), N.S.C.K. (T33KT669; T35MS9695; NIH Loan Repayment Program 1L40AA031140-01), E.M.T.D. and J.d.l.F. (R01MH120219 and R01AG073593; P30AG066614), J.D.T. (1K01MH141330), T.T.M. (K08MH135343), M.R. (5T32HG010464), A.A.P. (P30DA060810), K.P.H. (R01HD092548 and R01HD114721), and D.M.D. (U10AA008401 and P50AA022537). E.M.T.D., K.P.H., J.d.l.F., and C.M.W. are members of the Population Research Center at The University of Texas at Austin, supported by NIH grant P2CHD042849.

This study was made possible by the generous public sharing of summary statistics from multiple published genome-wide association studies and consortia, including the Psychiatric Genomics Consortium (PGC), the Million Veteran Program (MVP), the International Cannabis Consortium, the GWAS & Sequencing Consortium of Alcohol and Nicotine use (GSCAN), the Social Science Genetic Association Consortium (SSGAC), the Genetics of Personality Consortium, the Broad Antisocial Behavior Consortium, and the PsycheMERGE Network. We thank the many studies that made these consortia possible, the researchers involved, and the participants in those studies, without whom this effort would not have been possible.

Data used in the preparation of this article were obtained from the Adolescent Brain Cognitive Development^SM^ (ABCD) Study (https://abcdstudy.org), held in the NIMH Data Archive (NDA). This is a multisite, longitudinal study designed to recruit more than 10,000 children age 9-10 and follow them over 10 years into early adulthood. The ABCD Study® is supported by the National Institutes of Health and additional federal partners under award numbers U01DA041048, U01DA050989, U01DA051016, U01DA041022, U01DA051018, U01DA051037, U01DA050987, U01DA041174, U01DA041106, U01DA041117, U01DA041028, U01DA041134, U01DA050988, U01DA051039, U01DA041156, U01DA041025, U01DA041120, U01DA051038, U01DA041148, U01DA041093, U01DA041089, U24DA041123, U24DA041147. A full list of supporters is available at https://abcdstudy.org/federal-partners.html. A listing of participating sites and a complete listing of the study investigators can be found at https://abcdstudy.org/consortium_members/. ABCD consortium investigators designed and implemented the study and/or provided data but did not necessarily participate in the analysis or writing of this report. This manuscript reflects the views of the authors and may not reflect the opinions or views of the NIH or ABCD consortium investigators.

This research used data from Add Health, a program project directed by K. M. Harris (principal investigator) and designed by J. R. Udry, P. S. Bearman and K. M. Harris at the University of North Carolina at Chapel Hill, and funded by grant P01HD031921 from the Eunice Kennedy Shriver NICHD, with cooperative funding from 23 other federal agencies and foundations. Information on how to obtain the Add Health data files is available on the Add Health website (https://addhealth.cpc.unc.edu/). This research used Add Health GWAS data funded by Eunice Kennedy Shriver NICHD grants R01HD073342 to K. M. Harris (principal investigator) and R01HD060726 to K. M. Harris, J. D. Boardman, and M. B. McQueen (multiple principal investigators).

We gratefully acknowledge All of Us participants for their contributions, without whom this research would not have been possible. We also thank the National Institutes of Health’s All of Us Research Program for making available the participant data examined in this study.

We thank Lea K. Davis for providing access to BioVU. Specifically, data were obtained from Vanderbilt University Medical Center’s BioVU, which is supported by numerous sources, including institutional funding, private agencies and federal grants. These include the NIH-funded shared instrumentation grant S10RR025141, and CTSA grants UL1TR002243, UL1TR000445 and UL1RR024975. Genomic data are also supported by investigator-led projects, including U01HG004798, R01NS032830, RC2GM092618, P50GM115305, U01HG006378, U19HL065962 and R01HD074711; and additional funding sources listed at https://victr.vumc.org/biovu-funding/.

The Collaborative Study on the Genetics of Alcoholism (COGA), Principal Investigators B. Porjesz, V. Hesselbrock, A. Agrawal; Scientific Director, A. Agrawal; Translational Director, D. Dick, includes ten different centers: University of Connecticut (V. Hesselbrock); Indiana University (H.J. Edenberg, T. Foroud, Y. Liu, M.H. Plawecki); University of Iowa Carver College of Medicine (S. Kuperman, A. Anderson); SUNY Downstate Health Sciences University (B. Porjesz, J. Meyers); Washington University in St. Louis (L. Bierut, A. Agrawal, S. Hartz); University of California at San Diego (M. Schuckit); Rutgers University (D. Dick, R. Hart, J. Salvatore, J. Tischfield); The Children’s Hospital of Philadelphia, University of Pennsylvania (L. Almasy); Icahn School of Medicine at Mount Sinai (A. Goate, P. Slesinger); and Howard University (D. Scott). Other COGA collaborators include: C. Holzhauer, M. Hesselbrock (University of Connecticut); D. Lai, J. Nurnberger Jr., L. Wetherill, X., Xuei, S. O’Connor, (Indiana University); J. Kramer (University of Iowa), G. Chan (University of Iowa; University of Connecticut); C. Kamarajan, A. Pandey, D.B. Chorlian, P. Barr, S. Kinreich, G. Pandey, Z. Neale, S., C. Chatzinakos, J. Zhang, Saenz deViteri, R. Christian, A. Bingly (SUNY Downstate); G. Pathak (Icahn School of Medicine at Mount Sinai); A. Anokhin, K. Bucholz, F. Dong, A. Hatoum, E. Johnson, V. McCutcheon, J. Rice, S. Saccone (Washington University); F. Aliev, Z. Pang, S. Kuo, S. Brislin, J. Moore (Rutgers University); A. Merikangas (The Children’s Hospital of Philadelphia and University of Pennsylvania); M. Gitik, NIAAA Staff Collaborator. We continue to be inspired by our memories of Henri Begleiter and Theodore Reich, founding PI and Co-PI of COGA, and also owe a debt of gratitude to other past organizers of COGA, including Ting-Kai Li, P. Michael Conneally, Raymond Crowe, and Wendy Reich, for their critical contributions. This national collaborative study is supported by NIH Grant U10AA008401 from the National Institute on Alcohol Abuse and Alcoholism (NIAAA) and the National Institute on Drug Abuse (NIDA).

We thank the Centre for Longitudinal Studies (CLS), University College London Social Research Institute, and the UK Data Service for the use of Millennium Cohort Study (MCS) cohort data and for making these data available, respectively. Neither the CLS nor the UK Data Service bears any responsibility for the analysis or interpretation of these data.

This research was conducted using the UK Biobank Resource under Application Numbers 10279 and 11425; we thank all UK Biobank participants for their contributions.

We would like to thank the research participants and employees of 23andMe Research Institute for making this work possible.

## Author Contributions

T.T.M., K.P.H., and D.M.D. designed the study and jointly oversaw the project. T.T.M. supervised the analytic team. T.T.M., K.P.H., D.M.D., and C.M.W. led drafting of the main text, with substantial input from A.A.P., S.S.R., R.K.L., E.M.T.D., and J.W.S. C.M.W. and T.T.M. led drafting of the Supplementary Information, with contributions from K.P.H., H.E.P., D.L.C., S.S.R., J.D.T., Y.N., N.S.C.-K., M.C., M.R., R.K.L., and E.M.T.D. C.M.W. and T.T.M. prepared the figures. C.M.W. prepared the tables, with assistance from H.E.P., D.L.C., Y.N., and N.S.C.K. C.M.W. served as lead analyst and conducted GWAS analyses, including quality control, meta-analysis, and Genomic SEM, with assistance from T.T.M., D.L.C., P.T.T., and M.C. C.M.W. and T.T.M. led the quasi-replication analyses, with assistance from J.D.T. and Y.N. C.M.W., H.E.P., D.L.C., Y.N., and N.S.C.K. conducted the polygenic index analyses, with assistance from R.K.L., P.B.B., F.A., M.R., and T.T.M. S.S.R. performed the BioVU PheWAS. C.M.W. and T.T.M. led fine-mapping and downstream bioinformatics analyses, with substantial contributions from N.Y.B., K.Y., T.G., T.P., and M.S. J.D.D., M.J., E.C.J., J.G., and H.Z. prepared the leave-one-out GWAS summary statistics. A.A.P., P.B.B., I.D.W., S.J.B., E.M.T.D., and J.d.l.F. provided feedback on study design and interpretation. All authors contributed to and critically reviewed the manuscript.

## Data Availability

All data sources are described in the **Supplementary Information** and listed in the Reporting Summary. No new data were collected for the present study. We analyzed only data from existing studies and cohorts, some of which are available under restricted access to protect participant privacy. The minimum dataset necessary to interpret, verify, and extend our findings (i.e., the externalizing GWAS summary statistics) can be requested at https://externalizing.rutgers.edu/request-data/. These results are based in part on data contributed by 23andMe Research Institute. In accordance with 23andMe’s data-sharing policy, public releases are limited to the top 10,000 SNPs. The full externalizing GWAS summary statistics can be provided to qualified investigators who enter into a data transfer agreement with 23andMe. Upon approval by 23andMe, the Externalizing Consortium will share the complete file. In addition, we will make available a version of the summary statistics that excludes restricted data available for academic use via the Externalizing Consortium website upon publication.

The LD scores and HapMap3 reference data used in the EUR-like Genomic SEM analyses were downloaded from: https://github.com/GenomicSEM/GenomicSEM/wiki. The 1000 Genomes Project Phase 3 v5 data used to calculate LD scores for the Popcorn and AFR-like Genomic SEM analyses were downloaded from: https://www.cog-genomics.org/plink/2.0/resources#phase3_1kg. HapMap 3 SNPs and reference data used for the AFR-like Genomic SEM and PRS-CSx analyses were downloaded from: https://github.com/getian107/PRScsx. Ensembl v110 and GTEx v8 datasets were downloaded from the FUMA website: https://fuma.ctglab.nl/downloadPage. Gene Ontology classifications were downloaded from the Molecular Signatures Database: https://www.gsea-msigdb.org/gsea/msigdb/collections.jsp. Single-cell/single-nucleus RNA sequencing (sc/snRNA-seq) data were downloaded from: https://github.com/tanyaphung/scrnaseq_viewer. BrainSpan data were downloaded from https://www.brainspan.org/static/download. GWAS Catalog results were obtained from: https://www.ebi.ac.uk/gwas/downloads/summary-statistics.

