## Supplementary Information for "Genomic insights into substance use and disinhibitory disorders"

**1. General introduction** **3**
**2. GWAS data acquisition and preprocessing** **4**
2.1. Introduction 4
2.2. Identifying novel GWAS of externalizing-related traits 4
2.3. Revising published GWAS to exclude polygenic prediction cohorts 4
2.4. Quality control of GWAS summary statistics 6
2.5. GWAS meta-analysis 6
**3. Genomic structural equation modeling** **7**
3.1. Introduction 7
3.2. Linkage disequilibrium score regression 7
3.3. Genomic factor analyses 9
3.4. Estimating heterogeneous genetic correlations with QTrait 13
3.5. Ancestry-specific multivariate GWAS of externalizing 17
3.6. Similarity of genetic liability to externalizing across ancestry groups 20
**4. Multi-ancestry GWAS, fine-mapping, and gene prioritization** **22**
4.1. Introduction 22
4.2. Multi-ancestry meta-analysis of externalizing 22
4.3. Fine-mapping putatively causal variants 22
4.4. Gene-based associations and prioritization 23
4.5. Identifying novel effector genes for externalizing 25
4.6. Estimating heterogeneous SNP associations with QSNP. 27
4.7. Evaluating the concordance of EXTEUR and EXTEUR−23andMe SNP-level signals 34
**5. Replication** **36**
5.1. Introduction 36
5.2. Methodology 36
5.3. Results 41
**6. Bioannotation and bioinformatic analyses** **43**
6.1. Introduction 43
6.2. Methods 44
6.2.1. Bioinformatic characterization of genome-wide architecture 44
6.2.2. Bioinformatic characterization of effector genes 48
6.3. Results 49
6.3.1. Bioinformatic characterization of genome-wide architecture 49
6.3.2. Bioinformatic characterization of effector genes 59
**7. Polygenic prediction** **60**

7.1. Introduction 60
7.2. Samples 60
7.2.1. Collaborative Study on the Genetics of Alcoholism (COGA) 61
7.2.2. National Longitudinal Study of Adolescent to Adult Health (Add Health) 61
7.2.3. Adolescent Brain and Cognitive Development (ABCD) Study 62
7.2.4. Millennium Cohort Study (MCS) 63
7.2.5. The UK Biobank (UKB) Siblings 64
7.2.6. Vanderbilt University Medical Center Biobank (BioVU) 65
7.3. Methodology 65
7.3.1. Polygenic index construction 65
7.3.2. Effect estimation framework 66
7.3.3. Cohort-specific analyses 68
7.4. Results 70
7.4.1. COGA 70
7.4.2. Add Health 71
7.4.3. Evaluating the impact of 23andMe data on predictive accuracy 74
7.4.4. ABCD 76
7.4.5. MCS 77
7.4.6. UKB 77
7.4.7. BioVU 78
**8. Deviations from the preregistration 81**
**9. References 83**

### 1. General introduction

We preregistered our plan to extend our prior multivariate genome-wide association study (GWAS) of externalizing[1], henceforth referred to as EXT1, by leveraging new discovery samples with greater statistical power and broader ancestral representation (https://osf.io/7pfgj/). Any deviations from the preregistration are outlined in **Supplementary Section 8**.

EXT1 was a multivariate gene discovery effort of externalizing behaviors, traits, and disorders in individuals whose genomes were most similar to reference samples from Europe (EUR-like). In that study, the externalizing spectrum was measured with seven indicator GWAS: (i) attention-deficit/hyperactivity disorder[2] (ADHD); (ii) problematic alcohol use (ALCP), a meta-analysis of alcohol dependence[3] and AUDIT-P scores[4]; (iii) lifetime cannabis use[5] (CANN), (iv) reverse-coded age at first sexual intercourse[4] (FSEX); (v) number of sexual partners[4] (NSEX); (vi) self-reported risk tolerance[4] (RISK); and (vii) lifetime smoking initiation[6] (SMOK).

Here, we build on EXT1 with three aims: to enhance locus discovery, to deepen understanding of associated biology, and to improve the accuracy and portability of polygenic prediction. Where available, we replace indicator GWAS with better powered summary statistics and add indicators that are theoretically relevant and available in European (EUR-like) or African-like (AFR-like) genetic similarity groups. Analyses are limited to EUR-like and AFR-like samples because no other ancestry group currently has sufficient GWAS data available for multivariate modeling. Throughout, we maintain harmonized trait definitions and quality control to preserve continuity with EXT1 while enabling a broader, cross-ancestry discovery framework.

### 2. GWAS data acquisition and preprocessing

**Section authors:** Camille M. Williams & Travis T. Mallard

#### 2.1. Introduction

This section outlines the procedures used to gather and generate univariate GWAS summary statistics related to externalizing traits, behaviors, and disorders. Our goal was to expand upon our previously published EXT1 GWAS[1] by (i) increasing the sample size for genomic discovery and (ii) considering the inclusion of new theoretically relevant GWAS results available in both EUR- and AFR-like populations. This was accomplished in three basic steps. First, we identified relevant GWAS summary statistics that met our pre-registered inclusion criteria. Second, we either obtained or generated versions of these GWAS summary statistics that excluded the samples selected for downstream analyses (e.g., variant-level replication, polygenic prediction). Third, we subjected all GWAS summary statistics to stringent quality control procedures, yielding the final set of input data for our multivariate genomic analyses.

#### 2.2. Identifying novel GWAS of externalizing-related traits

For the analyses in EUR-like ancestry groups, we identified three new GWAS for the phenotypes included in the EXT1 GWAS with larger sample sizes, including ADHD[7], ALCP[8], and SMOK[9]. We additionally found three externalizing-related GWAS that were not available at the creation of the EXT1 GWAS and were conducted in both EUR- and AFR-like individuals: cannabis use disorder (CUD)[10], opioid use disorder (OUD)[11], and tobacco use disorder (TUD)[12] (**Supplementary Table 1–2**).

#### 2.3. Revising published GWAS to exclude polygenic prediction cohorts

To facilitate downstream polygenic prediction analyses in independent samples, we revised several univariate GWAS to exclude target cohorts. Specifically, we worked with study analysts to generate revised versions of the ALCP, SMOK, and TUD GWAS summary statistics that excluded participants from the UK Biobank (UKB)[13], the National Longitudinal Study of Adolescent to Adult Health (Add Health)[14,15], the Collaborative Studies on the Genetics of Alcoholism (COGA)[16], and Vanderbilt University Medical Center's biobank (BioVU)[17]. However, given that we only planned to use the sibling subsample from UKB for polygenic prediction, we included GWAS of unrelated UKB participants in the discovery-stage analyses. Several of these revised UKB GWAS, which exclude the sibling subsample, were previously conducted and described in EXT1[1]: RISK, NSEX, FSEX, SMOK, and AUDIT-P (i.e., the UKB component of ALCP). Below, we describe the procedures used to conduct a GWAS of TUD in UKB that excluded the sibling subsample (restricted to EUR-like ancestry individuals). Revised UKB GWAS were then meta-analyzed with their respective phenotypic GWAS (**Supplementary Section 2.5**).

We first created a harmonized TUD phenotype using the same case-control criteria from the published TUD[12] GWAS. Cases were defined based on the presence of International Classification of Diseases (ICD)-10 codes F17.0–F17.4, F17.9, or Z72.0, and controls based on the absence of all of these codes. Individuals with the ICD-10 code Z71.6 (tobacco use counseling) were excluded from the control group. As done in EXT1[1], we excluded individuals in the UK Biobank sibling hold-out sample and their close relatives (pairwise KING coefficient ≥ 0.0884). Participants were excluded from analysis if they: had withdrawn consent, exhibited outlier heterozygosity or missingness rates, showed evidence of sex chromosome aneuploidy, self-reported a sex that was discrepant with their genetically inferred sex, or had insufficient data (e.g., missing genetic principal components). To minimize potential bias due to population stratification, we restricted the sample to individuals of genetically inferred European ancestry.

We then used REGENIE[18] v2.2.4, a two-step ridge regression-based linear mixed model optimized for large-scale genetic analyses, to conduct a GWAS of TUD for the typed and imputed genetic data. In Step 1 of the REGENIE pipeline, we used directly genotyped, high-quality variants to fit a null model that accounts for population structure and relatedness, including sex, birth year, genotyping batch, and the first 40 genetic principal components as covariates. Step 1 was completed using a logistic regression model with a block size of 1000. In Step 2 of the REGENIE pipeline, we used the imputed genotype data to test for associations between single-nucleotide polymorphisms (SNPs) and TUD, adjusting for the same covariates as in Step 1. These SNP associations were estimated using approximate Firth logistic regression, applying a Firth/saddle-point approximation correction at $P \leq 0.01$. The resulting GWAS summary statistics were subsequently subjected to quality control procedures and meta-analyzed with the other relevant cohorts, as described in the following section (**Supplementary Section 2.5**).

### 2.4. Quality control of GWAS summary statistics

All GWASs considered for this project included SNPs from the 22 autosomes imputed using a large reference panel matched to the appropriate ancestry group. To be considered for inclusion, we required that each GWAS meet the following minimal quality control (QC) criteria: samples genomes were most similar to reference samples from Europe or Africa; participants had available data for all relevant covariates (e.g., age or birth year, sex, genetic principal components); individuals were successfully genotyped genome-wide (> 95%); and variants and samples passed standard QC filtering and exclusions (e.g., ancestry outliers, sex mismatches).

Prior to analysis, we applied a standardized QC pipeline to all GWAS using EasyQC[19] v23.8, following the procedures outlined in EXT1[1]. This pipeline has been adapted from the Social Science Genetic Association Consortium (SSGAC) protocol and modified to align with the Genomic SEM preprocessing workflow, as previously described[4,20]. Briefly, SNPs were excluded if they: were absent from the reference panel for the relevant ancestry; strand ambiguous; contained missing or invalid values (e.g., $P$ values outside [0,1]); exhibited a low minor allele frequency (MAF < 0.005); exhibited poor imputation quality (INFO < 0.9); were indels, multi-allelic, or non-autosomal variants; had invalid or duplicated chromosomal positions; or could not be aligned to the reference panel. The EUR- and AFR-like samples of the 1000 Genomes Project Phase 3 v5 dataset[21] were used as a reference panel for these QC procedures.

### 2.5. GWAS meta-analysis

Where appropriate, we used METAL[22] v2011-03-25 to meta-analyze the revised indicator GWAS results and our UKB GWAS results that excluded siblings, employing a sample-size weighted approach. For continuous traits, weights were based on the total sample size ($N$). For binary traits, we used the sum of the effective sample size ($\sum N_{\text{eff}}$) as the weights[4,20]. $N_{\text{eff}}$ for each cohort was calculated using the formula: $4 \times v \times (1 - v) \times N$, where $v$ is the sample prevalence. We meta-analyzed GWAS summary statistics for the ALCP, TUD, and SMOK EUR-like indicators and the SMOK AFR-like indicator. To generate the ALCP GWAS for EUR-like individuals, we meta-analyzed the case-control alcohol use disorder (AUD) GWAS[8] with the quantitative trait AUDIT-P GWAS[1], treating the resulting meta-analysis as a quantitative trait for the purposes of weighting and interpretation.

### 3. Genomic structural equation modeling

**Section authors**: Camille M. Williams, Diego Londono-Correa, Elliot M. Tucker-Drob, K. Paige Harden, & Travis T. Mallard

#### 3.1. Introduction

As done in EXT1[1], we used Genomic SEM[23] v0.0.5 to model the variance and covariance structure among indicators and conduct a multivariate GWAS of externalizing. We previously conducted exploratory and confirmatory factor analyses to model the observed covariances among a set of variables related to externalizing traits, behaviors, and disorders, drawing on both theoretical considerations and exploratory data inspection. After testing a series of competing models, we identified a single common factor model with seven externalizing indicator phenotypes as the best-fitting solution. The indicator phenotypes included (i) attention-deficit/hyperactivity disorder (ADHD[2]); (ii) problematic alcohol use (ALCP[3,4]), a meta-analysis of a case-control GWAS (alcohol dependence) and a continuous trait GWAS (Alcohol Use Disorder Identification Test Problems subscale; AUDIT-P); (iii) lifetime cannabis use (CANN[5]), (iv) reverse-coded age at first sexual intercourse (FSEX[4]); (v) number of sexual partners (NSEX[4]); (vi) self-reported risk tolerance (RISK[4]); and (vii) lifetime smoking initiation (SMOK[6]).

Here, we build on our prior study by modeling genetic variances and covariances among an updated set of GWAS indicators with improved statistical power, estimating a single common factor of externalizing liability in both EUR- and AFR-like ancestry groups. After selecting a final set of indicators for the externalizing factor model in EUR-like and AFR-like individuals, we proceeded to: (i) fit ancestry-specific common factor models (**Supplementary Section 3.3)**, (ii) evaluate genetic relationships between externalizing and other complex traits (**Supplementary Section 3.4**), (iii) estimate the effects of individual SNPs on a latent externalizing factor (**Supplementary Section 3.5**), and (iv) examine the degree to which genetic liability to externalizing is similar across ancestry groups (**Supplementary Section 3.6**).

#### 3.2. Linkage disequilibrium score regression

GWAS summary statistics were first processed using the *munge* function from the GenomicSEM[23] package (v0.0.5) in R[24], retaining ancestry-specific HapMap3[25] SNPs outside of the major histocompatibility complex regions with an imputation quality (INFO) score ≥ 0.90

(when available). Alleles with a minor allele frequency (MAF) < 0.01 were excluded from analysis for EUR-like data, while alleles with a MAF < 0.05 were excluded from analysis for AFR-like data. The more conservative threshold in AFR-like data was selected to ameliorate concerns about potential mismatch between the reference panel and the GWAS source data.

We then used GenomicSEM's *ldsc* function to apply multivariable linkage disequilibrium score regression (LDSC) to the munged ADHD, ALCP, CANN, CUD, FSEX, NSEX, RISK, SMOK, OUD, and TUD GWASs from EUR-like individuals, as well as the munged ALCP, CUD, SMOK, OUD, and TUD GWASs from AFR-like individuals. For each indicator, we estimated the SNP-based heritability ($h^2$), evaluated the polygenic signal (e.g., mean $\chi^2$), and assessed the extent of confounding bias from population stratification (quantified by the LDSC intercept and attenuation ratio). We then created genome-wide sampling and covariance matrices as input for downstream modeling analyses. Data from the 1000 Genomes Project Phase 3 v5[21] European and African super populations were used to create LD scores for each ancestral group, filtering for MAF ≥ 0.005. Per-chromosome genotype files (chromosomes 1–22) were generated using PLINK[26] (v1.9.0-b.7.7 and v2.0.0-a.6.12), and LD scores were computed using LDSC[27] v1.0.1 with a 1 Mb window around each SNP.

To estimate SNP heritability on the liability scale, Genomic SEM requires both the sample and population prevalences for binary traits. For these traits, we followed the developers' recommendations[23] to use the $\sum N_{\text{eff}}$ method and set the sample prevalence to 0.5. The exceptions were CANN and RISK, for which we used sample prevalences of 0.265 and 0.260, respectively, to be consistent with EXT1[1]. For population prevalences in the EUR-like GWASs, we used the following values: 0.05 for ADHD[7,28], 0.22 for CANN[5], 0.026 for CUD[29,30], 0.021 for OUD[11,31,32], 0.26 for RISK[4], 0.46 for SMOK[1], and 0.32 for TUD[33]. For the AFR-like GWASs, we used population prevalence estimates of 0.22 for ALCP[8,34], 0.028 for CUD[35–37], 0.012 for OUD [38,39], 0.240 for TUD[33], 0.380 for SMOK[40–42]. Values were set to N/A for continuous traits ALCP, NSEX, and FSEX.

The EUR-like GWASs were modestly to moderately heritable (SNP $h^2$ = 0.062–0.176) and showed strong signal (mean $\chi^2$ = 1.159–5.601). The AFR-like GWASs were also modestly to moderately heritable (SNP $h^2$ = 0.049–0.142) and showed discernible signal (mean $\chi^2$ = 1.029–1.174). The intercepts were close to 1, and the attenuation ratios (defined as (intercept − 1) / (mean $\chi^2$ − 1)) were close to 0, suggesting that there is minimal detectable bias due to population stratification (**Supplementary Tables 3–4**). The multivariable LDSC genetic covariance matrix and its sampling covariance matrix were used for downstream factor modeling.

#### 3.3. Genomic factor analyses

We estimated ancestry-specific common factor models that were informed by EXT1[1]. Specifically, we used the *usermodel* function from GenomicSEM[23] to fit a confirmatory factor model to the multivariable LDSC genetic and sampling covariance matrices. At this stage, the scale of the factor was defined by fixing its variance to unity (i.e., unit variance identification). Model fit was assessed using preregistered thresholds for conventional indices in SEM: the model $\chi^2$ statistic, the Akaike information criterion (AIC), the comparative fit index (CFI), and the standardized root mean square residual (SRMR). All of these indices retain their standard interpretations within the Genomic SEM framework[23], with the exception of the model $\chi^2$ statistic. As $\chi^2$ tests tend to be overpowered and yield significant results in large samples, such as those analyzed in GWAS, the model $\chi^2$ statistic was used as a comparative measure of fit (similar to AIC) rather than as an absolute measure of fit. For CFI and SRMR, values greater than 0.90 and less than 0.08, respectively, were considered indicative of good model fit (i.e., the specified latent variable structure adequately explained the observed covariances among the variables).

***Indicator selection and model fitting.*** Based on our GWAS inclusion criteria (**Supplementary Section 2**), we identified 10 GWASs as potential indicators for the updated EUR-like externalizing factor model ($EXT_{EUR}$): ADHD, ALCP, CANN, CUD, FSEX, NSEX, OUD, RISK, SMOK, and TUD. Similarly, we identified five GWASs for the AFR-like externalizing factor model ($EXT_{AFR}$): ALCP, CUD, OUD, SMOK, and TUD. We considered several criteria when selecting which indicators would be selected for the final factor models, including: the theoretical relevance of each indicator externalizing, the availability of data in both EUR- and AFR-like ancestry groups (**Supplementary Section 2**), the number of non-overlapping samples contributed by each GWAS (**Supplementary Tables 1–2**), the SNP overlap across indicator GWASs, the strength and quality of the GWAS signal, the genetic correlations between indicators as assessed with LDSC within the Genomic SEM framework (**Supplementary Section 3.2)**, and the impact of including or excluding different indicators on the externalizing model parameters and fit statistics.

First, we investigated whether we could replace CANN with CUD in the $EXT_{EUR}$ model, which also included ADHD, ALCP, FSEX, NSEX, RISK, and SMOK. We sought to select a cannabis-related phenotype that was theoretically more closely aligned with externalizing and also available in AFR-like individuals. Notably, CANN was based on a single item assessing lifetime use, whereas CUD reflects problematic use and is conceptually more closely related to other substance use and disinhibitory phenotypes linked to externalizing. Although we preregistered that we would include CANN in the $EXT_{EUR}$ model, we elected to replace it with CUD for the following reasons: First, CUD GWAS data were available for both EUR-like and AFR-like ancestry groups, with a larger sample size of EUR-like participants. Second, compared to CANN, CUD provided a larger number of individuals who were not included in any other indicator GWAS (**Supplementary Table 1**). The CUD GWAS included an additional 24,544 individuals from the Mass General Brigham Biobank (previously known as Partners Biobank), whereas the CANN GWAS only included a total of 4,101 additional unique individuals from the Hospital Universitari Vall d'Hebron (HUVH), TRacking Adolescents' Individual Lives Survey (TRAILS), the UK Household Longitudinal Study (UKHLS), and the Brisbane Longitudinal Twin Study (BLTS) datasets. Note that although the Genomic SEM framework allows for sample overlap across cohorts, there are substantial gains in power when including GWAS with non-overlapping samples[23]. Third, LDSC analyses revealed that although both GWAS had similar SNP coverage, the CUD GWAS had a slightly stronger polygenic signal (mean $\chi^2$ = 1.279) that was less biased by population stratification (intercept = 0.994, s.e. = 0.008) than the CANN GWAS (mean $\chi^2$ = 1.244, intercept = 1.022, s.e. = 0.007). Fourth, CUD was more strongly genetically correlated with other indicators (median $r_g$ = 0.615) compared to CANN (median $r_g$ = 0.403; **Supplementary Table 3**). Fifth, although loadings for the other phenotypes remained largely stable when swapping out CANN for CUD, the loading of CUD on the externalizing factor ($\lambda$ = 0.854, s.e. = 0.024) was substantially higher than that of CANN ($\lambda$ = 0.573, s.e. = 0.028), and the overall model fit improved when using CUD ($\chi^2(14)$ = 446.528, AIC = 474.528, CFI = 0.941, SRMR = 0.067) instead of CANN ($\chi^2(14)$ = 1078.224, AIC = 1106.224, CFI = 0.863, SRMR = 0.094). Although CANN and CUD were not genetically correlated enough to potentially induce rank deficiency in the empirical genetic covariance matrix ($r$ = 0.499, s.e. = 0.044), we replaced CANN with CUD, since it was more relevant to the genetic liability of externalizing and available in both ancestry groups.

We next evaluated whether adding TUD and OUD as indicators in the $EXT_{EUR}$ and $EXT_{AFR}$ models would improve our ability to model a cross-cutting factor of externalizing liability and advance gene discovery. TUD and OUD were considered strong candidates because they are clinical substance use phenotypes that were available in both ancestry groups, but they were ultimately excluded from discovery-stage analyses. Our decision to exclude OUD and TUD from the $EXT_{EUR}$ and $EXT_{AFR}$ models was informed by several points: First, all cohorts included in the OUD GWAS were already included in the other indicator GWAS, and the TUD GWAS only added a modest number of unique individuals from the Pennsylvania Medicine Biobank ($N_{EUR}$ = 28,999; $N_{AFR}$ = 10,088) (**Supplementary Tables 1–2**). Second, when munging the OUD GWAS with the HapMap 3 reference panel, we found that there was poor SNP coverage in both EUR- and AFR-like data ($M$ = 634,788 and $M$ = 301,517, respectively). This problem was also observed when preparing the summary statistics for multivariate GWAS, where we found that OUD had poor SNP coverage and overlap with other indicators in the model. Together, the inclusion of TUD and OUD as indicators would result in losing approximately 37% of SNPs (3,974,858/6,331,678) in our $EXT_{EUR}$ GWAS and approximately 48% of SNPs (3,672,532/7,115,963) in our $EXT_{AFR}$ GWAS. Third, LDSC analyses showed that, although the OUD GWAS was genetically correlated with the established indicator GWAS (median EUR-like $r_g$ = 0.598, median AFR-like $r_g$ = 0.326), it was poorly correlated with TUD in AFR-like samples ($r_g$ = 0.092, s.e. = 0.162). SMOK and TUD were moderately to highly genetically correlated (EUR $r_g$ = 0.833, s.e. = 0.029; AFR-like $r_g$ = 0.641, s.e. = 0.108) and were similarly genetically correlated with other indicators in both ancestry groups (median EUR-like $r_g$ SMOK = 0.612 versus TUD = 0.653, median AFR-like $r_g$ SMOK = 0.359 versus TUD = 0.245). Given the similarity between TUD and SMOK, we considered that including both in the same model might introduce redundant elements and ultimately elected to retain only SMOK as an indicator given its greater utility in advancing gene discovery (**Supplementary Table 3**).

In our final common factor models, we selected ADHD, ALCP, CUD, FSEX, NSEX, RISK, and SMOK as indicators for $EXT_{EUR}$, and ALCP, CUD, and SMOK as indicators for $EXT_{AFR}$. Results for these analyses are discussed in the main text and reported in **Supplementary Table 4**.

***Sensitivity analyses without 23andMe data.*** To provide the community with readily available GWAS summary statistics, we ran analyses that excluded 23andMe data from the

$EXT_{EUR}$ model, henceforth referred to as $EXT_{EUR-23andMe}$. Here, we focus our sensitivity analyses on the $EXT_{EUR}$ model, as 23andMe contributed a large number of unique individuals to the discovery sample. Demonstrating robustness in this setting should, in turn, generalize to analyses in which 23andMe comprised a smaller share of the discovery samples (e.g., $EXT_{AFR}$).

We assessed the impact of removing 23andMe data by comparing model parameters, genetic correlations, and GWAS results in this section, as well as polygenic prediction results in **Supplementary Section 7**. SMOK was the only indicator phenotype that included 23andMe data. Excluding this cohort from the SMOK GWAS reduced the sum of effective sample size from 2,995,104 to 1,180,799 and the mean $\chi^2$ statistic from 5.601 to 2.263. The SNP-based $h^2$ estimate remained stable (0.135 when including 23andMe, 0.132 when excluding 23andMe). Despite the reduction in sample size, the $EXT_{EUR-23andMe}$ model fit the data well ($\chi^2(14) = 403.765$, AIC = 431.765, CFI = 0.940, and SRMR = 0.066), and its fit was similar to that of the full model ($\chi^2(14) = 446.528$, AIC = 474.528, CFI = 0.941, SRMR = 0.067). Factor loadings were virtually identical, with a median absolute difference of 0.008. Overall, we found that the inclusion or exclusion of 23andMe data had a negligible impact on the structure and model fit of $EXT_{EUR}$ (**Supplementary Table 5**).

***Model fitting comparisons to EXT1.*** Finally, we compared our $EXT_{EUR}$ model to EXT1, which differed in terms of sample sizes and one indicator, as described above. The $EXT_{EUR}$ model also differed from EXT1 in that it did not include residual covariances among indicators. Briefly, we found that model fit was similar between the published EXT1 model ($\chi^2(12) = 354.747$, AIC = 386.747, CFI = 0.957, and SRMR = 0.078) and the updated $EXT_{EUR}$ model ($\chi^2(14) = 446.528$, AIC = 474.528, CFI = 0.941, and SRMR = 0.067). Factor loadings were very similar, with a median absolute difference of 0.086. Overall, despite the large increase in sample size from about ~1.5 million to ~3 million individuals and modest changes to the model structure and specification, parameters and fit were concordant and comparable across EXT1 and $EXT_{EUR}$ (**Supplementary Table 6**).

#### 3.4. Estimating heterogeneous genetic correlations with $Q_{Trait}$

Building upon the practices and results of EXT1[1], we used a series of genetic correlation and $Q_{Trait}$ analyses to evaluate the construct validity of our factor in AFR-like and EUR-like ancestry groups. Specifically, we used the $Q_{Trait}$ function[43] from the Genomic SEM[23] v0.0.5

package to (i) estimate ancestry-specific genetic correlations between the EXT factor and other complex traits, and (ii) evaluate whether relationships between model indicators and those other traits plausibly operated via the EXT factor. To identify appropriate GWASs of clinically, socially, or medically relevant traits, we conducted a broad review of the literature and selected those that had a sample size (or effective sample size) $\geq$ 10,000 in EUR-like populations or $\geq$ 8,000 in AFR-like populations. For analyses of EUR-like data, we built upon the list of external phenotypes included in the EXT1 GWAS study. The final list included 96 traits studied in individuals of EUR-like ancestry and 23 traits studied in individuals of AFR-like ancestry.

External trait summary statistics were standardized and aligned using the *munge* function from the Genomic SEM[23] v0.0.5 package. Paralleling the procedures described in **Supplementary Section 3.2**, we set the MAF filter to 0.01 for EUR-like data and 0.05 for AFR-like data, with the INFO filter set to 0.9 for both ancestries. We again used trait-specific population and sample prevalences to accommodate liability-scale transformations for binary traits, using the $\sum N_{\text{eff}}$ approach referenced above. All prevalence values and their sources are provided in **Supplementary Tables 7–10**, with citations provided for relevant GWASs or public health sources, to ensure transparency and replicability. Population prevalences for binary traits were identified using a standardized, reproducible protocol tailored to the type of GWAS:

- *Phenotypes from meta-analyzed GWAS.* When available, the population prevalence reported in the original GWAS manuscript (as used for LDSC-based SNP-heritability estimation) was used. If not reported, we obtained estimates from the Global Burden of Disease (GBD) database (https://vizhub.healthdata.org/gbd-results/), ensuring population-based comparability across traits. If neither source was available, we used values from recent meta-analyses or large-scale epidemiological reviews in the published literature.
- *Phenotypes from UK Biobank (UKB)-only GWAS.* Preference was given to the population prevalence reported in the original GWAS. If unavailable, we used the sample prevalence from UKB, given its population-based sampling frame. If the UKB sample prevalence was clearly unrepresentative, we used the GBD database estimate or, if unavailable, a value from the literature.
- *Phenotypes from Million Veteran Program (MVP)-only GWAS.* We used the prevalence reported in the GWAS publication when available. If unavailable, we relied on GBD

database estimates due to MVP's ascertainment and non-population-based sampling. If neither was available, we referenced published meta-analyses or reviews.

In addition to estimating genetic correlations with these traits, we also used $Q_{\mathrm{Trait}}$ analyses to evaluate whether the factor mediates genetic associations between the external traits and the model indicators. The $Q_{\mathrm{Trait}}$ statistic, or heterogeneity index, is based on $\chi^2$ difference tests comparing two competing models: (1) the common pathway model, where the external correlate predicts only the common factor; and (2) the independent pathways model, where the external correlate predicts the individual indicator phenotypes. The $\chi^2$ difference between these two models is referred to as the $Q_{\mathrm{Trait}}$ statistic[43]. To determine statistical significance for this statistic, we used a Bonferroni-corrected *P* threshold based on the number of external correlates for which the function independently computes the heterogeneity indices. A significant $Q_{\mathrm{Trait}}$ statistic indicates that the association between the external trait and the EXT indicators is not fully explained by the common factor, suggesting trait-specific associations with one or more individual indicators. We also computed the local standardized root mean squared residual (lSRMR), a global effect size index of heterogeneity that quantifies the magnitude of the discrepancies between the empirical LDSC-derived genetic correlations implied by the common pathway model.

Trait correlations were flagged as heterogenous if: (1) the genetic correlation between the external correlate and the common factor exceeded a Bonferroni-corrected *P* threshold, (2) the $Q_{\mathrm{Trait}}$ statistic was significant after Bonferroni correction for the number of external correlates for which the function independently computes the heterogeneity indices, and (3) the lSRMR exceeded both an absolute threshold of 0.10 and 25% of the mean observed genetic correlations between the external correlate and the indicator phenotypes that load on the factor. For heterogeneous traits, we fit a series of follow-up models, iteratively freeing direct paths between the external trait and specific indicators until the model was saturated (i.e., d.f. = 0) or no further significant heterogeneity was detected. If heterogeneity is present, we report the genetic correlation of the follow-up model in the main and supplementary text.

In the present study, we conducted $Q_{\mathrm{Trait}}$ analyses for the $\mathrm{EXT}_{\mathrm{EUR}}$, the $\mathrm{EXT}_{\mathrm{AFR}}$, the $\mathrm{EXT}_{\mathrm{EUR-23andMe}}$, and the EXT1 models. We used ancestry-specific LD scores calculated from the 1000 Genomes Project Phase 3 v5 European and African super populations[21]. For $\mathrm{EXT}_{\mathrm{EUR}}$, analyses were based on the seven indicators (ADHD, ALCP, FSEX, NSEX, CUD, SMOK, RISK) and 96 external traits (**Supplementary Table 7**). For $\mathrm{EXT}_{\mathrm{AFR}}$, analyses were based on three indicators (ALCP, CUD, SMOK) and 23 external traits (**Supplementary Table 8**). $Q_{\text{Trait}}$ analyses for $\text{EXT}_{\text{EUR−23andMe}}$ paralleled $\text{EXT}_{\text{EUR}}$, with the only difference being that SMOK excluded data from 23andMe (**Supplementary Table 9**). For EXT1, we performed analyses using the seven original indicators (ADHD, ALCP, FSEX, NSEX, CANN, SMOK, RISK) and the same external traits included in the $\text{EXT}_{\text{EUR}}$ analyses, except for cannabis initiation, which was an indicator in the EXT1 model(**Supplementary Table 10**).

Note that we found several of the complex traits in AFR-like samples were ultimately unsuitable for $Q_{\text{Trait}}$ analyses. Of the 23 traits identified, 2 had negative heritability estimates, and 7 had non-significant heritability estimates. In EUR-like data, hoarding symptoms was the only trait with a non-significant heritability estimate and, consequently, was excluded from subsequent analyses.

We applied a Bonferroni correction for multiple comparisons based on the number of correlations examined per model ($\text{EXT}_{\text{EUR}}$ and $\text{EXT}_{\text{EUR−23andMe}}$ $P \leq 0.05/95 = 5.26\text{e-}4$, EXT1 $P \leq 0.05/94 = 5.32\text{e-}4$, and $\text{EXT}_{\text{AFR}}$ $P \leq 0.05/14 = 3.57\text{e-}3$).

***$\text{EXT}_{\text{EUR}}$ $Q_{\text{Trait}}$ analyses.*** We found that the $\text{EXT}_{\text{EUR}}$ factor was genetically correlated with 74% (70/95) of the external traits related to clinical, health, and social outcomes, with coefficients ranging from −0.688 (age of smoking initiation) to 0.899 (antisocial behavior) after adjusting for multiple comparisons (**Supplementary Table 7**). Of these associations, 41% (29/70) were heterogeneous according to the $Q_{\text{Trait}}$ statistics. Via the $Q_{\text{Trait}}$ function, we proceeded to fit a series of follow-up models for each of these traits, iteratively freeing direct paths between the trait and indicators until no further significant heterogeneity was detected. This resulted in a median absolute change of 0.026 in the genetic correlation estimates. The maximum decrease in coefficients was observed for household income (0.092), and the maximum increase was observed for sensation seeking (0.094). The $Q_{\text{Trait}}$ results suggest that while factor-trait correlations are largely unchanged after freeing a specific path, ADHD shares unique pleiotropic relationships with several phenotypes that are not fully explained by the EXT factor.

The $\text{EXT}_{\text{EUR}}$ factor was associated with a broad range of clinically and socially relevant traits, including lower educational achievement and socioeconomic status. Specifically, $\text{EXT}_{\text{EUR}}$ was most positively genetically associated with a range of externalizing, social, and substance use phenotypes, including antisocial behavior ($r_g = 0.899$, s.e. = 0.067), opioid use disorder ($r_g = 0.836$, s.e. = 0.043), maternal smoking around birth ($r_g = 0.816$, s.e. = 0.024), a substance use latent factor

($r_g$ = 0.763, s.e. = 0.025), Townsend deprivation index ($r_g$ = 0.744, s.e. = 0.049), borderline personality disorder ($r_g$ = 0.712, s.e. = 0.030), stress-related disorder ($r_g$ = 0.686, s.e. = 0.047), and suicide attempt ($r_g$ = 0.665, s.e. = 0.035). The $EXT_{EUR}$ factor was most negatively genetically associated with age of smoking initiation ($r_g$ = −0.661, s.e. = 0.027), mother's age at death ($r_g$ = −0.608, s.e. = 0.073), father's age at death ($r_g$ = −0.478, s.e. = 0.035), household income ($r_g$ = −0.344, s.e. = 0.018), and educational attainment ($r_g$ = −0.350, s.e. = 0.017).

***$EXT_{AFR}$ $Q_{Trait}$ analyses.*** The $EXT_{AFR}$ factor was significantly positively correlated with depression ($r_g$ = 0.737, s.e. = 0.226) and tobacco use disorder ($r_g$ = 0.806, s.e. = 0.111) after correction for multiple comparisons (**Supplementary Table 8**). There were no significant $Q_{Trait}$ relationships, suggesting that correlations between the external traits and the EXT indicators (ALCP, CUD, and SMOK) were well accounted for by the EXT factor. Opioid use disorder, post-traumatic stress disorder, number of drinks per week, and schizophrenia were also positively correlated over 0.5; however, these associations had large standard errors and did not reach statistical significance.

***$EXT_{EUR-23andMe}$ $Q_{Trait}$ analyses.*** When conducting analyses for the $EXT_{EUR-23andMe}$ model, we found similar genetic correlations and $Q_{Trait}$ results to those reported for $EXT_{EUR}$ (**Supplementary Table 9**). Genetic correlations between the externalizing factor and external traits were virtually identical between the $EXT_{EUR}$ and the $EXT_{EUR-23andMe}$ model (correlation of coefficients: $r$ = 0.998, s.e. = 0.006). All significant factor-trait correlations in the $EXT_{EUR}$ model were also significant in the $EXT_{EUR-23andMe}$ model. The $Q_{Trait}$ results were also highly concordant between the $EXT_{EUR-23andMe}$ and $EXT_{EUR}$ models. Although the proportion of significant $Q_{Trait}$ relationships in the $EXT_{EUR-23andMe}$ model was comparable to that of the $EXT_{EUR}$ model (41%, 29/70), bipolar disorder and attentional symptoms had a significant $Q_{Trait}$ statistic in the $EXT_{EUR}$ model, whereas carbohydrate consumption and the psychopathology factor had significant $Q_{Trait}$ statistics in the $EXT_{EUR-23andMe}$ model. Although the $Q_{Trait}$ $P$ value was significant for both of these phenotypes in the $EXT_{EUR}$ model, the lSRMR either did not exceed an absolute threshold of 0.10 or 25% of the mean observed genetic correlations between the external correlate and the indicator phenotypes. Collectively, the $Q_{Trait}$ results suggest that the $EXT_{EUR}$ and the $EXT_{EUR-23andMe}$ factors measure the same underlying construct.

***EXT1 $Q_{Trait}$ analyses.*** When conducting genetic correlation and $Q_{Trait}$ analyses for the EXT1 factor, we found that results were similar to those reported for the $EXT_{EUR}$ model. Genetic correlations between the externalizing factor and external traits were highly similar across the $EXT_{EUR}$ and EXT1 models (correlation of coefficients: $r = 0.948$, s.e. = 0.033). The $Q_{Trait}$ results showed greater variability between $EXT_{EUR}$ and EXT1, though, with the EXT1 factor being the more genetically heterogeneous factor (**Supplementary Table 10**). Of the significantly associated external trait correlations, 41% (29/70) were heterogeneous for the $EXT_{EUR}$ factor compared to 71% (46/65) for the EXT1 factor according to the $Q_{Trait}$ analyses.

#### 3.5. Ancestry-specific multivariate GWAS of externalizing

Prior to estimating ancestry-specific SNP associations with the EXT factor, we first used the *sumstats* function from GenomicSEM[23] to align effect alleles across indicators and standardize effect estimates and standard errors. To maximize coverage for discovery of high-quality common variants, we excluded SNPs with a MAF < 0.005 or an INFO score < 0.60 (when this information was available). This yielded a final set of 6,331,678 SNPs for analysis in the EUR-like data and 7,115,963 SNPs for analysis in the AFR-like data. We then estimated the SNP effects on the $EXT_{EUR}$ **(Figure S1)** and $EXT_{AFR}$ (**Figure S2**) latent genetic factors using diagonally weighted least squares (DWLS) and ancestry-matched LD scores from the 1000 Genomes Project Phase 3 v5 European and African super populations[21].

The estimated sample size ($\widehat{N}$) of the factor was calculated using the approach described by Mallard and colleagues[44], averaging $1/((2 \times MAF \times (1 - MAF)) \times s.e.^2)$ across SNPs, restricted to variants with $0.10 \leq MAF \leq 0.40$ for more stable estimates. The $\widehat{N}$ of the $EXT_{EUR}$ GWAS was 3,943,075, compared to 1,492,085 for the EXT1 GWAS. In line with our model fitting results described above, we found that the genetic correlation between $EXT_{EUR}$ genomic factor and EXT1 was indistinguishable from unity ($r_g = 0.990$, s.e. = 0.036).

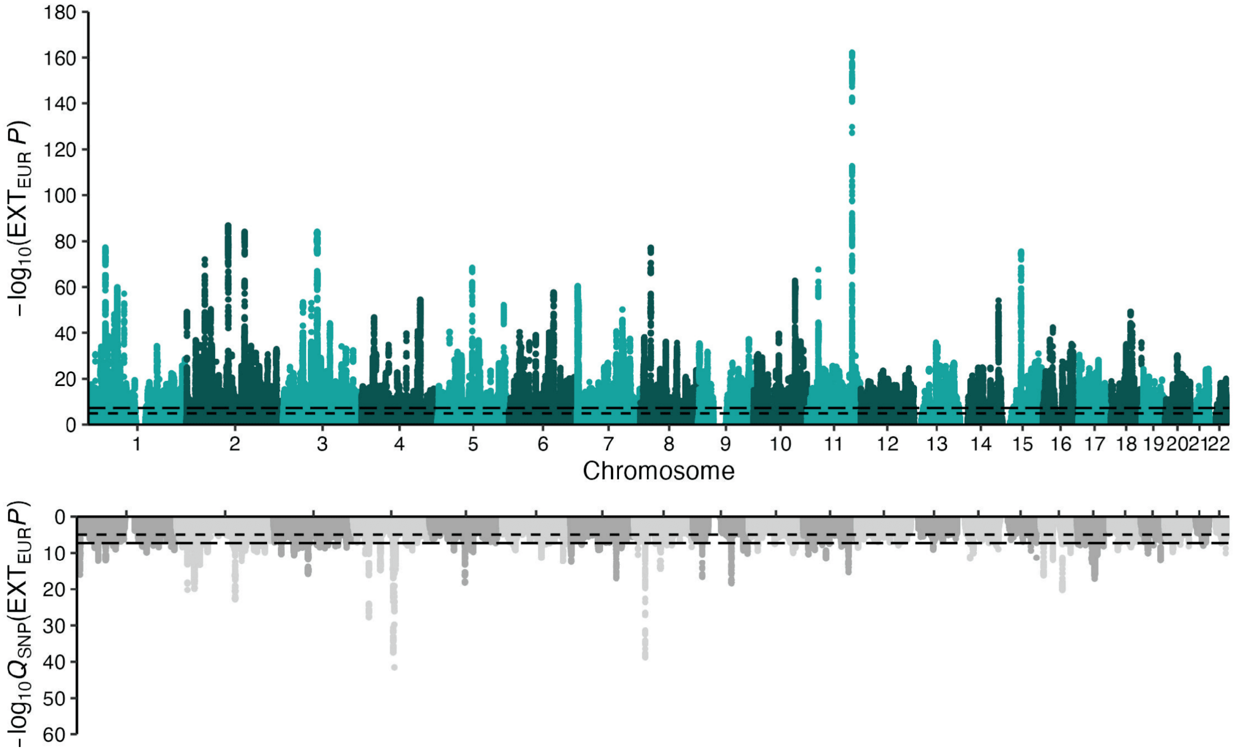


***Figure S1. Multivariate genome-wide association analysis of externalizing in individuals from European-like (EUR) ancestry.*** Scatterplot of $-\log_{10}$($P$ for two-sided $Z$ test) for weighted least-squares regression to estimate GWAS associations (top) and $-\log_{10}$($P$ for one-sided $\chi^2$ test with 7 – 1 d.f.) for $Q_{\text{SNP}}$ tests of heterogeneity (bottom) for EXT estimated with the Genomic SEM[23] v0.0.5 package. The line with small dashes corresponds to $-\log_{10}$(1e-5) and the line with wider dashes to $-\log_{10}$(5e-8).

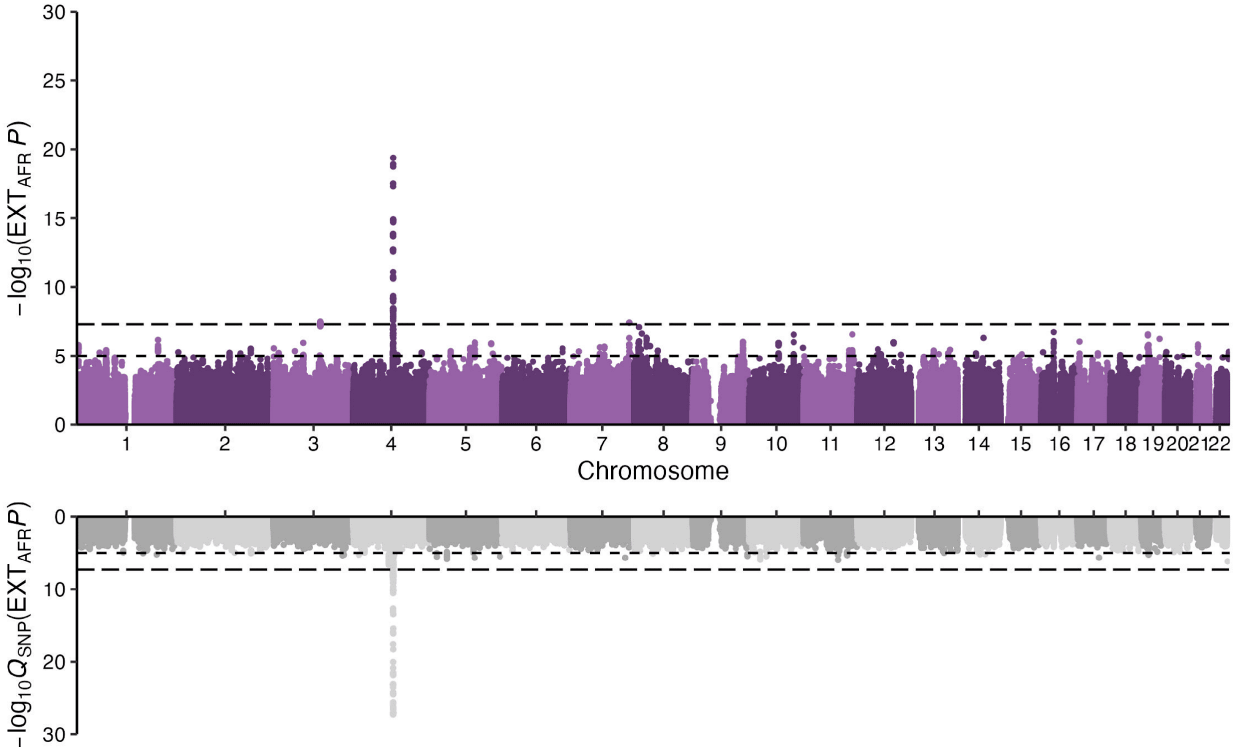


***Figure S2. Multivariate genome-wide association analysis of externalizing in individuals from African-like (AFR) ancestry.*** Scatterplot of $-\log_{10}$($P$ for two-sided $Z$ test) for weighted least-squares regression to estimate GWAS associations (top) and $-\log_{10}$($P$ for one-sided $\chi^2$ test with 3 – 1 d.f.) for $Q_{SNP}$ tests of heterogeneity (bottom) for EXT estimated with the Genomic SEM[23] v0.0.5 package. The line with small dashes corresponds to $-\log_{10}$(1e-5) and the line with wider dashes to $-\log_{10}$(5e-8).

#### 3.6. Similarity of genetic liability to externalizing across ancestry groups

Prior to conducting downstream bioinformatic and polygenic index analyses across ancestry groups, we first sought to test whether genetic liability for externalizing was similar across those groups. We evaluated the cross-ancestry similarity through a series of three analyses. First, we compared the $EXT_{EUR}$ and $EXT_{AFR}$ factor model parameters. Second, we assessed whether the genetic correlations between EXT and related complex traits were consistent across ancestry groups. Third, we directly estimated the cross-ancestry genetic correlation between $EXT_{EUR}$ and $EXT_{AFR}$.

***Cross-ancestry model comparisons.*** To facilitate cross-ancestry comparisons of joint genetic architecture, we created a truncated version of the $EXT_{EUR}$ model that mirrored the configuration of the $EXT_{AFR}$ model (i.e., using only ALCP, CUD, and SMOK as indicators). All indicators in the truncated model loaded significantly onto the externalizing factor, with loadings ranging from 0.729 (s.e. = 0.022) to 0.916 (s.e. = 0.029). Moreover, we found that this truncated $EXT_{EUR}$ model factor was correlated at unity with the full $EXT_{EUR}$ GWAS ($r_g = 1$, s.e. = 0.029), suggesting that the identified polygenic signal is consistent even when estimated from three indicators. As with the $EXT_{AFR}$ model, we found that CUD had the highest loading in the truncated $EXT_{EUR}$ model (**Supplementary Table 3**).

***Similarity of factor-trait genetic correlations across ancestries.*** As described in **Supplementary Section 3.4**, we estimated genetic correlations between the externalizing factor and other complex traits that were clinically, socially, or medically relevant to externalizing. Importantly, this was done for both $EXT_{EUR}$ and $EXT_{AFR}$, allowing us to quantify the extent to which genetic correlations with other traits were similar across ancestry groups.

Here, we used an errors-in-variables approach to calculate a disattenuated correlation coefficient to quantify the similarity of those estimates. This approach corrected for attenuation due to measurement error, defined here as the sampling variability of the genetic correlation estimates. Specifically, we computed the disattenuated correlation by taking the cross-trait covariance of the observed pairs and dividing it by the product of the error-corrected standard deviations in each ancestry, obtained by subtracting the average squared standard error from the corresponding across-trait variance. Confidence intervals were obtained by a nonparametric bootstrap over traits.

We found that the correlation between the externalizing factor and 14 external traits was highly similar across ancestry groups (disattenuated $r$ = 0.950, 95% CI: 0.816–1.127). These results suggest that, despite large standard errors in the AFR-like estimates, the pattern of associations between the externalizing and other complex traits is broadly consistent across ancestry groups.

***Cross-ancestry genetic correlation.*** As a final test, we estimated the cross-ancestry genetic correlation $EXT_{EUR}$ and $EXT_{AFR}$ using Popcorn[45] v1.1. Specifically, we used Popcorn to calculate cross-ancestry LD scores and estimate the genetic correlation between the $EXT_{EUR}$ and $EXT_{AFR}$ GWAS summary statistics. Cross-ancestry LD scores were computed for 4,607,783 SNPs that were available in both of the 1000 Genomes Project Phase 3 v5 European and African super populations[21], had a MAF $\geq$ 0.01, and were not in the major histocompatibility complex region. Popcorn's regression-based model was then used to estimate the cross-ancestry genetic correlation between the $EXT_{EUR}$ GWAS from the seven indicator model and the $EXT_{AFR}$ GWAS, using 1,364,903 SNPs present in both $EXT_{EUR}$ and $EXT_{AFR}$ after filtering on MAF $\geq$ 0.01. Specifically, we calculated both the genetic-impact correlation, representing the correlation between ancestry-specific allele variance-normalized SNP effect sizes, and the genetic-effect correlation, reflecting the correlation between per-allele SNP effect sizes. The genetic impact correlation was 1.409 (s.e. = 0.296, $P$ = 0.168), and the genetic effect correlation was 1.475 (s.e. = 0.318, $P$ = 0.136). Note that the genetic correlation estimates are not constrained between −1 and +1, and that the $P$ reported is for a test that the genetic correlation is less than 1.

Collectively, the results provided strong evidence that $EXT_{EUR}$ and $EXT_{AFR}$ indexed the same cross-cutting liability for disinhibition with similar genetic architectures. Based on this, we proceeded with cross-population analyses, including fine-mapping, bioinformatics, and polygenic prediction.

### 4. Multi-ancestry GWAS, fine-mapping, and gene prioritization

Section authors: Camille M. Williams, K. Paige Harden, Maia Choi, and Travis T. Mallard

#### 4.1. Introduction

After establishing that the genetic architecture of externalizing was similar across ancestry groups, we (i) conducted a cross-ancestry meta-analysis of externalizing (**Supplementary Section 4.2**); (ii) leveraged both ancestry-specific GWAS for fine-mapping analyses (**Supplementary Section 4.3**) and gene prioritization analyses (**Supplementary Section 4.4**); (iii) identified novel effector genes (**Supplementary Section 4.5**), (iv) examined heterogeneity in the SNP level effects of genome-wide associations, fine-mapped variants, and their corresponding effector genes (**Supplementary Section 4.6**), and (v) evaluated the concordance of SNP-level associations after removing 23andMe data (**Supplementary Section 4.7**)

#### 4.2. Multi-ancestry meta-analysis of externalizing

Using METAL[22] v2011-03-25, we conducted a fixed-effects sample-size-weighted meta-analysis of the $EXT_{EUR}$ and $EXT_{AFR}$ GWAS summary statistics. The meta-analysis weights were set to the estimated sample size ($\widehat{N}$), which was calculated using the approach described by Mallard and colleagues[44], averaging $1/((2 \times MAF \times (1 - MAF)) \times s.e.^2)$ across SNPs, restricted to variants with $0.10 \leq MAF \leq 0.40$ for more stable estimates. Meta-analyzed results were primarily used to define approximately independent regions harboring significant associations across ancestry groups in the fine-mapping analyses (**Methods**), for data visualization purposes (**Figure 3**), and in the binomial sign concordance test (**Supplementary Section 5**).

#### 4.3. Fine-mapping putatively causal variants

We used the $EXT_{EUR}$ and $EXT_{AFR}$ GWAS results to identify putatively causal variants associated with externalizing. Specifically, we used SuSiEx[46] v1.1.2 to perform cross-population fine-mapping within each unified locus (**Methods**). We prioritized variants based on their posterior inclusion probability (PIP) values, defining candidates at two confidence thresholds: PIP ≥ 0.5 for moderate confidence and PIP ≥ 0.95 for high confidence. We identified 647 putative causal SNPs across 485 regions with a moderate level of confidence, and 214 SNPs across 181 regions with high confidence (**Supplementary Table 11**). Inspection of $Q_{SNP}$ signals for these variants revealed that only 1% (5/485) of moderate-confidence loci, and a single high-confidence locus, harbored a causal variant with significantly heterogeneous effects .

#### 4.4. Gene-based associations and prioritization

***Ancestry-specific gene-based association testing.*** We used MAGMA[47] v1.10 to conduct gene-based association tests, mapping SNP-based signals to protein-coding genes within and across ancestry groups. Consistent with best practices, we used the SNP-wise mean model to map SNP-level signals to genes on autosomal chromosomes based on physical location, using gene boundaries defined by Ensembl build 110 (19,429 genes). Gene-based association analyses were conducted separately in EUR-like and AFR-like data with ancestry-matched LD reference data from the 1000 Genomes Project Phase 3 v5 European and African super populations[21], yielding ancestry-specific gene-level *Z* statistics and gene-gene correlation matrices. To benchmark gains attributable to our increased statistical power, we also used this pipeline to re-analyze our EXT1[1] GWAS. A Bonferroni correction was used to control for multiple testing, dividing the standard alpha (0.05) by the number of genes tested in each gene-based GWAS.

Briefly, we found that 16% (2,971/18,259) of genes were significant in the gene-based GWAS of $EXT_{EUR}$, representing a roughly threefold increase compared to EXT1, in which 5% (943/18,318) of genes were significant (**Supplementary Table 14**). Only four genes out of 18,685 were significantly associated in the gene-based GWAS of $EXT_{AFR}$, two of which were significant in the gene-based GWAS of $EXT_{EUR}$. Importantly, we found that 97% (911/943) of the significant genes in the EXT1 analysis were also significant in the better powered $EXT_{EUR}$ analysis.

***Multi-ancestry gene-based association testing.*** To calculate multi-ancestry gene-based association results, we used MAGMA's standard implementation of Stouffer's method to meta-analyze signals across populations. As there is no sample overlap across ancestral populations, this simplified to

$$Z_i^{meta} = \frac{\sum_k \; w_k Z_i^{(k)}}{\sqrt{\sum_k \; w_k^2}},$$

where the meta-analytic Z statistic for gene *i* is an average across *k* populations, weighted by the square root of the corresponding sample size. Genes present in either population were allowed to enter the meta-estimate. Similarly, we used MAGMA's default meta-analytic procedures to generate a multi-ancestry gene-gene correlation matrix as

$$R_{ij}^{meta} = \frac{\sum_k \; w_{k,i} w_{k,j} R_{ij}^{(k)}}{\sum_k \; w_{k,i} w_{k,j}},$$

where the correlation between genes $i$ and $j$ is an average across $k$ populations with $w$ sample size weights. As expected, the multi-ancestry results were highly similar but not identical to the $EXT_{EUR}$ results: Approximately 15% (2,898/18,813) of genes were significant in the gene-based GWAS of $EXT_{META}$, and 94% (2,791/2,971) of $EXT_{EUR}$ significant genes were also significant in the $EXT_{META}$ analysis.

***Multimodal gene prioritization.*** We used FLAMES[48] v1.1.2, a novel framework for predicting the most likely effector gene within a locus, to identify the specific genes underlying externalizing liability. FLAMES is a multi-modal method that requires three inputs: PIP values from a fine-mapping analysis (here, the SuSiEx results described above), gene-based test statistics from MAGMA (described above), and polygenic prioritization scores from PoPS[49].

Prior to effector gene prioritization, we therefore used PoPS v0.2 to calculate polygenic prioritization scores for each gene in the multi-ancestry MAGMA results. Briefly, PoPS uses gene-level associations to learn the joint polygenic enrichments of gene features derived from cell-type-specific gene expression, biological pathways, and protein-protein interactions, and then uses these enrichments to calculate a priority score for every protein-coding gene in the feature space. Following the suggestions of the FLAMES developers, we used default parameters and the default feature set for all PoPS analyses. We also performed this analysis excluding pathway-based features (i.e., pathway-naïve PoPS) to ensure that any downstream analysis of FLAMES prioritized genes would not be susceptible to false positives caused by non-independence of training and testing data.

Following the recommendations of FLAMES developers, we also included GTEx-based tissue-specific weights to identify the most likely effector genes underlying externalizing. We also repeated gene prioritization using the pathway-naïve PoPS scores, as described above. All FLAMES analyses were employed using standard procedures and default settings.

Across the 1,294 genomic regions associated with externalizing, FLAMES identified 961 unique effector genes with a cumulative precision $\geq 0.75$ and 239 genes with a cumulative precision $\geq 0.999$ (**Supplementary Table 24**). Overall, the pathway-informed and pathway-naive effector genes identified by FLAMES were largely consistent. When running FLAMES with pathway-naive PoPS scores, we re-identified 922 (96%) of the 961 effector genes identified in the full pathway-informed analysis. The cumulative precision scores ranged from 0.757 – 0.894 for the 39 genes identified as effector genes in the pathway-informed analysis but not in the naive pathway analysis (median = 0.822). The cumulative precision scores ranged from 0.751 – 0.900 for the 39 genes identified as effector genes in the pathway-naive analysis but not in the pathway-informed analysis (median = 0.809).

#### 4.5. Identifying novel effector genes for externalizing

To identify novel genes related to externalizing, we examined whether pathway-informed FLAMES effector genes had previously been associated with externalizing-related traits in the GWAS Catalog[50]. Keywords used to search the GWAS Catalog included cannabis, smoking, externalizing, alcohol, risk, conduct disorder, alcohol, methamphetamine, nicotine, illegal, disinhibition, opioid, tobacco, alcohol use disorder, oppositional defiant disorder, ADHD, cigarettes, irritable mood, sex, sexual, alcoholic, drug, antisocial behavior, substance use, and aggressive.

We downloaded the genetic associations file of all traits related to externalizing from the GWAS Catalog on November 17th, 2025, including:

- aggressive behavior (22 associations, EFO_0003015),
- nicotine dependence (147 associations, EFO_0003768),
- attention deficit hyperactivity disorder (2,501 associations, EFO_0003888),
- drug dependence (1,021 associations, EFO_0003890),
- risk factor (147 associations, EFO_0003919),
- conduct disorder (71 associations, EFO_0004216),
- heroin dependence (11 associations, EFO_0004240),
- smoking behavior (1,652 associations, EFO_0004318),
- alcohol drinking (1,524 associations, EFO_0004329),
- methamphetamine dependence (3 associations, EFO_0004701),
- alcohol and nicotine codependence (10 associations, EFO_0004776),
- alcohol withdrawal (1 association, EFO_0004777),
- nicotine use (26 associations, EFO_0005430),
- illegal drug consumption (30 associations, EFO_0005431),
- non-substance related disinhibited behaviour (24 associations, EFO_0005432),

- opioid dependence (338 associations, EFO_0005611),
- smoking initiation (4,937 associations, EFO_0005670),
- cigarettes per day measurement (741 associations, EFO_0006525),
- smoking status measurement (3,068 associations, EFO_0006527),
- behavioural disinhibition measurement (161 associations, EFO_0006946),
- drug use measurement (3,653 associations, EFO_0007010),
- cannabis dependence (152 associations, EFO_0007191),
- cannabis use (71 associations, EFO_0007585),
- cannabis use initiation (14 associations, EFO_0007586),
- longitudinal alcohol consumption measurement (50 associations, EFO_0007645),
- oppositional defiant disorder dimensions in attention-deficit hyperactivity disorder (34 associations, EFO_0007679),
- alcohol dependence measurement (308 associations, EFO_0007835),
- ADHD symptom measurement (67 associations, EFO_0007860),
- cannabis dependence measurement (37 associations, EFO_0008457),
- risk-taking behaviour (1,060 associations, EFO_0008579),
- alcohol exposure measurement (8 associations, EFO_0009113),
- tobacco smoke exposure measurement (428 associations, EFO_0009115),
- nicotine dependence symptom count (295 associations, EFO_0009262),
- maximum cigarettes per day measurement (98 associations, EFO_0009264),
- alcohol use disorder measurement (408 associations, EFO_0009458),
- irritability measurement (67 associations, EFO_0009594),
- age at first sexual intercourse measurement (328 associations, EFO_0009749),
- opioid use measurement (23 associations, EFO_0009937),
- bitter alcoholic beverage consumption measurement (97 associations, EFO_0010092),
- opioid overdose severity measurement (6 associations, EFO_0010140),
- cocaine use disorder (10 associations, EFO_0010445),
- cocaine use measurement (12 associations, EFO_0010553),
- opioid use disorder (118 associations, EFO_0010702),
- age at initiation of smoking (65 associations, EFO_0021784),

- borderline personality disorder (28 associations, HP_0012076),
- addictive alcohol use (21 associations, HP_0030955),
- alcohol abuse (164 associations, MONDO_0002046),
- substance abuse (755 associations, MONDO_0002491),
- substance-related disorder (3,431 associations, MONDO_0002494),
- alcohol dependence (299 associations, MONDO_0007079),
- alcohol-related disorders (809 associations, MONDO_0021698),
- social inhibition quality (584 associations, OBA_1000278),
- aggressive behavior quality (148 associations, OBA_1000376),
- alcohol consumption quality (3,933 associations, OBA_1000840),
- sexual activity behaviour attribute (181 associations, OBA_2045293), and
- smoking behavior trait (4,318 associations, OBA_2050116).

There were no available associations for the age of onset of alcohol dependence (OBA_2001016), oppositional defiant disorder (HP_0010865), irritability (HP_0000737), and drug misuse (EFO_0011049). We then compared these genes to our effector gene list. Non-overlapping genes were classified as novel externalizing-related genes.

A total of 9,020 distinct genes were associated with one or more externalizing-related traits in the GWAS Catalog. Of the 961 effector genes linked to externalizing liability in the present paper, 207 (22%) had not previously been associated with any externalizing-related trait, behavior, or disorder in the GWAS Catalog, including 28 genes with a high cumulative precision score at unity (> .999; **Supplementary Table 24**).

#### 4.6. Estimating heterogeneous SNP associations with $Q_{SNP}$.

We conducted $Q_{SNP}$ analyses to evaluate heterogeneity in the SNP-level effects across externalizing phenotypes for fine-mapped variants and their associated effector genes. The $Q_{SNP}$ approach compares model fit between two scenarios: one where the SNP influences the factor and another where it affects each indicator or trait individually. This comparison helps determine if SNP associations act through the latent factor or are better explained by indicator-specific effects. For more information, refer to the original Genomic SEM paper[23].

For each multivariate GWAS, we first calculated the ratio of GWAS mean $\chi^2$ to $Q_{SNP}$ mean $\chi^2$, broadly quantifying the strength of the polygenic signal relative to heterogeneity (i.e., signal-to-noise ratio). We found that this ratio was 2.804 (5.862/2.091) for $EXT_{EUR}$ and 1.104 (1.160/1.051) for $EXT_{AFR}$, indicating that there was more polygenic signal than heterogenous signal for both GWAS. When investigating the specific significant $Q_{SNP}$ signals for each GWAS, we found that 0.162% (10,238/6,331,678) of SNPs had a significant $Q_{SNP}$ $P \leq$ 5e-8 in the $EXT_{EUR}$ GWAS, and 0.002% (108/7,115,963) of SNPs had a $Q_{SNP}$ $P \leq$ 5e-8 in the $EXT_{AFR}$ GWAS. We then examined whether any of these heterogeneous signals overlapped with the putative causal variants identified via fine-mapping. Among the 647 SNPs fine-mapped with moderate confidence (PIP ≥ 0.5), we found that 9 (1%) had a significant $Q_{SNP}$ signal in the $EXT_{EUR}$ GWAS (rs4702, rs2970931, rs62250713, rs1383722, rs34269355, rs4385369, rs1155397, rs2565059, and rs2974454) and 1 (< 1%) had a significant $Q_{SNP}$ signal in the $EXT_{AFR}$ GWAS (rs2066702). Among the 214 SNPs fine-mapped with very high confidence (PIP ≥ 0.95), we found that 5 (2%) had a significant $Q_{SNP}$ signal in the $EXT_{EUR}$ GWAS (rs4702, rs62250713, rs4385369, rs1155397, rs2565059) and no significant $Q_{SNP}$ signals in the $EXT_{AFR}$ GWAS.

As reported above, we used FLAMES to identify the most likely effector genes underlying externalizing liability. One component of FLAMES involves the integration of PIP values from fine-mapped credible sets, helping to connect putative causal variants to putative causal genes. However, it stands to reason that prioritized genes that map to $Q_{SNP}$ signals may not truly operate via the shared pathway—or that there is at least a nontrivial indicator-specific effect. Accordingly, we tested whether prioritized genes were linked to fine-mapped variants exhibiting significantly heterogeneous effects (**Supplementary Table 24**), thereby better distinguishing shared-factor signals from indicator-specific signals and refining effector gene prioritization.

The AFR $Q_{SNP}$ SNP signal (rs2066702) did not map onto any effector gene. Eight out of nine $EXT_{EUR}$ $Q_{SNP}$ variants with a PIP ≥ 0.5 mapped onto effector genes (rs2974454 did not map onto any gene). For each of these eight genes, we concisely summarize the patterns of association among fine-mapped variants below:

- ***CADM2***. We identified 84 variants across 5 credible sets mapping to the Cell Adhesion Molecule 2 (*CADM2*) gene. Only one of these sets, consisting of a single SNP (rs62250713; PIP ≥ 0.95), contained a significant $EXT_{EUR}$ $Q_{SNP}$ signal.
- ***FURIN.*** The $EXT_{EUR}$ $Q_{SNP}$ variant rs4702 (PIP ≥ 0.95) was the only variant mapping onto the Furin, Paired Basic Amino Acid Cleaving Enzyme (*FURIN*) gene.

- ***CHRNA2.*** For the Cholinergic Receptor Nicotinic Alpha 2 Subunit (*CHRNA2*) gene, five variants across two credible sets were identified, and all had significant $\text{EXT}_{\text{EUR}}$ $Q_{\text{SNP}}$ $P$ values.
- ***FOXP2.*** There were 13 variants across 3 credible sets mapping to the Forkhead Box P2 (*FOXP2*) gene. Two of the three credible sets included $\text{EXT}_{\text{EUR}}$ $Q_{\text{SNP}}$ signals: one set consisting of a single variant with a PIP ≥ 0.95 (rs1155397), and another with 11 SNPs. Across these 12 significant $Q_{\text{SNP}}$ variants, examination of the SNP-trait associations as a function of the unstandardized factor loadings revealed that the SNP effect on SMOK was generally inconsistent with the shared pathway model (**Figure S3**). The third credible set included a single variant (rs6466499) that was consistent with the common factor model (i.e., non-significant $Q_{\text{SNP}}$ $P$) with a PIP ≥ 0.95, which was notably independent of the $Q_{\text{SNP}}$ signal rs1155397 ($R^2 < 0.001$, $P = 0.369$). These findings suggest that there is likely more than one causal variant in the locus mapping to *FOXP2*, with a mixture of shared and trait-specific effects.

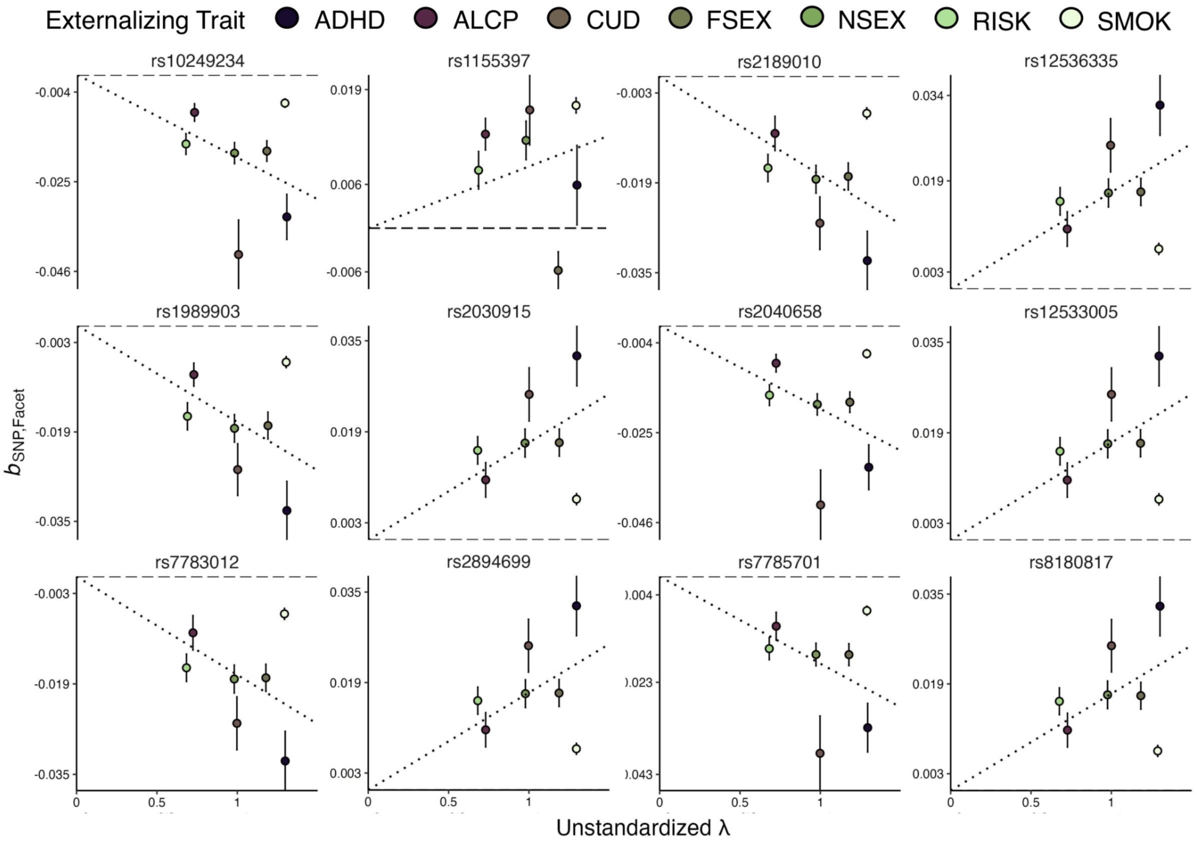


***Figure S3. Univariate SNP effects as a function of factor loadings for all significant FOXP2 loci with significant $Q_{SNP}$ associations*** Scatter plots depicting the indicator GWAS estimates for all significant $Q_{\text{SNP}}$ associations as a function of unstandardized factor loadings. Error bars represent standard errors of the beta coefficient. The dashed line represents the line of best fit, as estimated with linear regression with the intercept fixed to zero. Deviations from this line lend insight into how significant $Q_{\text{SNP}}$ $P$ arises. For example, the positive effect of rs8180817 (bottom right) on SMOK is inconsistent with the common factor model, driving the significant $Q_{\text{SNP}}$ $P$.

- ***MAD1L1***. For the Mitotic Arrest Deficient 1 Like 1 (*MAD1L1*) gene, two variants comprised their own credible set, one of which (rs4385369; PIP ≥ 0.95) had a significant $EXT_{EUR}$ $Q_{SNP}$ signal. Follow-up examination indicated that the positive effect of rs4385369 on RISK was inconsistent with the common factor model (**Figure S4a)**. Although the variant (rs35273555; PIP ≥ 0.95) in the second credible set was consistent with the common factor model (i.e., non-significant $Q_{SNP}$ *P*), it was not independent of the $Q_{SNP}$ signal rs4385369 ($R^2 = 0.865$, $P < 1.00e\text{-}4$).
- ***DPP4***. For the Dipeptidyl Peptidase 4 (*DPP4*) gene, two variants—each forming its own credible set—were identified. Both exhibited significant $Q_{SNP}$ *P* values in the $EXT_{EUR}$ GWAS (additionally, only rs2970931 had a PIP ≥ 0.5).
- ***TTC29***. There were 28 variants across 2 credible sets that mapped to the Tetratricopeptide Repeat Domain 29 (*TTC29*) gene. All four SNPs from one set had significant $EXT_{EUR}$ $Q_{SNP}$ signals (including rs34269355 with a PIP ≥ 0.5). Across these SNPs, the null-to-negative effects on RISK and NSEX appeared to be generally inconsistent with the common factor model (**Figure S4b**). The 24 remaining SNPs from the second set did not have significant $Q_{SNP}$ *P* values, suggesting that there may be a mixture of causal variants associated with *TTC29* that operate through the common factor.
- ***CENPC***. There were 55 variants across 3 credible sets that mapped to the Centromere Protein C (*CENPC*) gene. All 30 SNPs from 2 of the sets had significant $EXT_{EUR}$ $Q_{SNP}$ signals, with rs1383722 being the only one with PIP ≥ 0.5. Follow-up inspection suggested that some SNP effects were incompatible with the common factor pathway (**Figure S5**). However, the third set, which consisted of 25 SNPs, did not contain any significant $Q_{SNP}$ signal, suggesting that only a subset of causal variants in the same locus mapping to the *CENPC* gene are inconsistent with the common factor model.

Collectively, these results suggest that the vast majority of prioritized genes are plausible effector genes for the transdiagnostic externalizing factor. *FURIN, CHRNA2, MAD1L1,* and *DPP4* appear to map to putative causal variants with significantly heterogeneous or trait-specific effects.

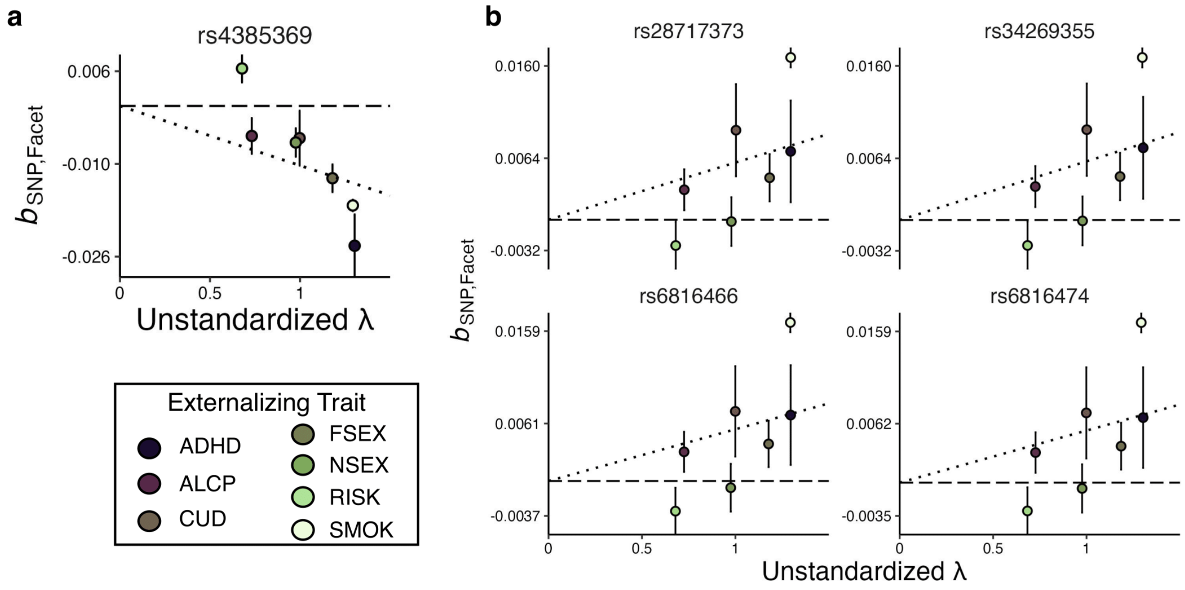


***Figure S4. Univariate SNP effects as a function of factor loadings for all (a) MAD1L1 and (b) TTC29 loci with significant $Q_{SNP}$ associations*** Scatter plots depicting the indicator GWAS estimates for all significant $Q_{SNP}$ associations as a function of unstandardized factor loadings. Error bars represent standard errors of the beta coefficient. The dashed line represents the line of best fit, as estimated with linear regression with the intercept fixed to zero. Deviations from this line lend insight into how significant $Q_{SNP}$ $P$ arises. For example, the larger positive effect of rs4385369 (top left) on RISK is inconsistent with the common factor model, driving the significant $Q_{SNP}$ $P$.

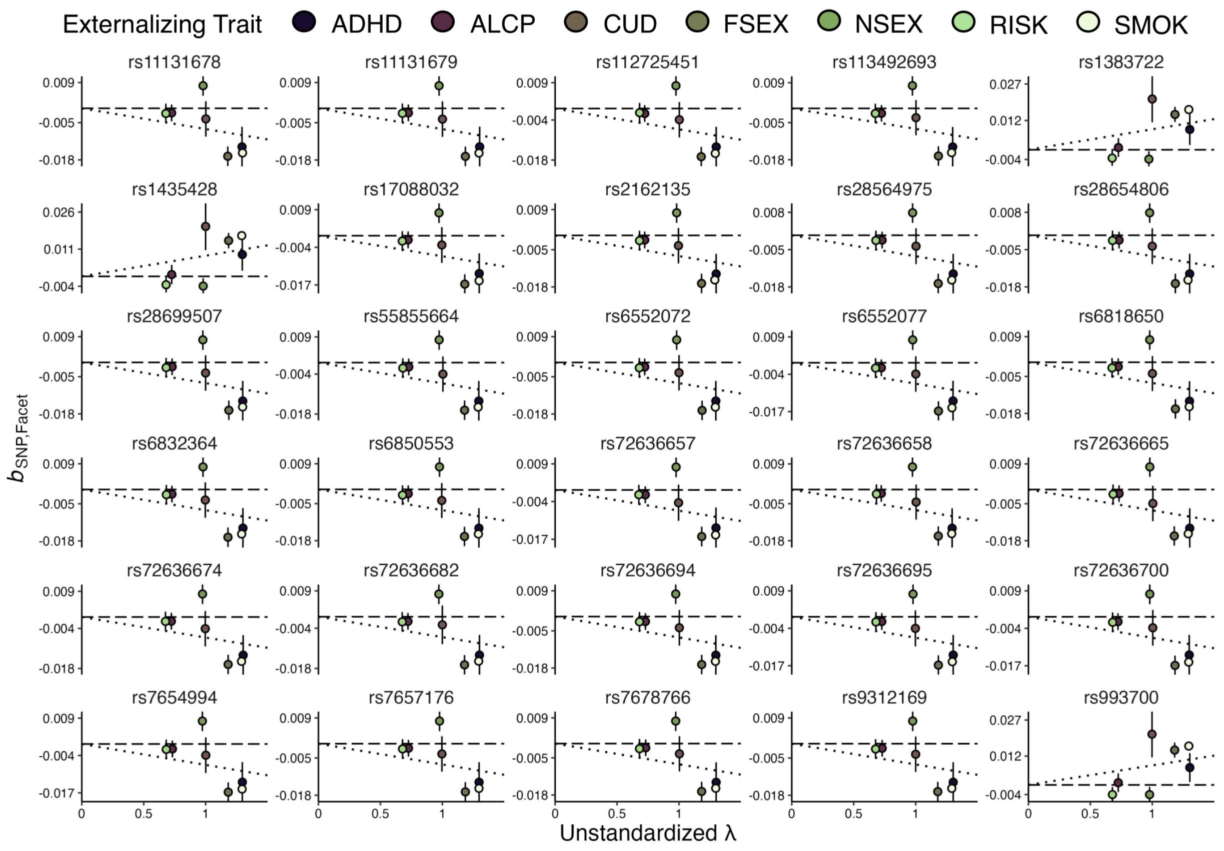


***Figure S5. Univariate SNP effects as a function of factor loadings for all CENPC loci with significant $Q_{SNP}$ associations*** Scatter plots depicting the indicator GWAS estimates for all significant $Q_{SNP}$ associations as a function of unstandardized factor loadings. Error bars represent standard errors of the beta coefficient. The dashed line represents the line of best fit, as estimated with linear regression with the intercept fixed to zero. Deviations from this line lend insight into how significant $Q_{SNP}$ *Ps* arise. For example, the larger positive effect of rs993700 (bottom right) on CUD is inconsistent with the common factor model, driving the significant $Q_{SNP}$ *P*.

### 4.7. Evaluating the concordance of $EXT_{EUR}$ and $EXT_{EUR-23andMe}$ SNP-level signals

To evaluate whether SNP-level signals were consistent across the $EXT_{EUR}$ and $EXT_{EUR-23andMe}$ GWAS results, we quantified the sign and effect size concordance between the two GWAS as recommended[51]. For this analysis, we focused on SNPs with a *P* value ≤ 1e-5 in the $EXT_{EUR}$ GWAS, as well as the 637 fine-mapped SNPs that had a PIP value ≥ 0.5 and a non-significant $Q_{SNP}$ statistic. Five SNPS (rs9265727, rs1330177, rs1330180, rs1997902, and rs134242) were not present in the $EXT_{EUR-23andMe}$ GWAS and were replaced with proxy SNPS (rs9265646, rs7850744, rs1934208, rs6044082, and rs134267, respectively).

Overall, the SNP-level signals were highly consistent across $EXT_{EUR}$ and $EXT_{EUR-23andMe}$ GWAS results. Although the $\hat{N}$ of the $EXT_{EUR-23andMe}$ GWAS was lower for the $EXT_{EUR-23andMe}$ GWAS ($\hat{N}$= 1,582,347) compared to the $EXT_{EUR}$ GWAS ($\hat{N}$= 3,943,075), the $EXT_{EUR}$ genomic factor and the $EXT_{EUR-23andMe}$ GWAS were genetically correlated at unity ($r_g$ = 1, s.e. = 0.028). We found that 100% of the 637 fine-mapped SNPs exhibited concordant signs across the two GWAS. The beta coefficients of these SNPs were highly correlated (*r* = 0.983, s.e. = 0.007) (**Figure S6a**), and only 17% (107/637) of these SNPs were identified as outliers (i.e., SNPs with an $EXT_{EUR-23andMe}$ estimate outside the 95% confidence interval of the $EXT_{EUR}$ estimate). Of the 381,281 SNPs with a $P \leq$ 1e-5 in the $EXT_{EUR}$ GWAS, 99.96% (381,105/381,281) were sign-concordant in the $EXT_{EUR-23andMe}$ GWAS. The beta coefficients for these SNPs were also highly correlated (*r* = 0.973, s.e. = 0.0004) (**Figure S6b**), and only 18% (69,123/381,281) of these SNPs were outliers.

For both SNP subsets, we observed a general attenuation of the *Z* statistics, as expected when down-sampling, since the *Z* statistic is a function of both sample size and the magnitude of the GWAS effect estimate (**Figure S6b**; **Figure S6d**).

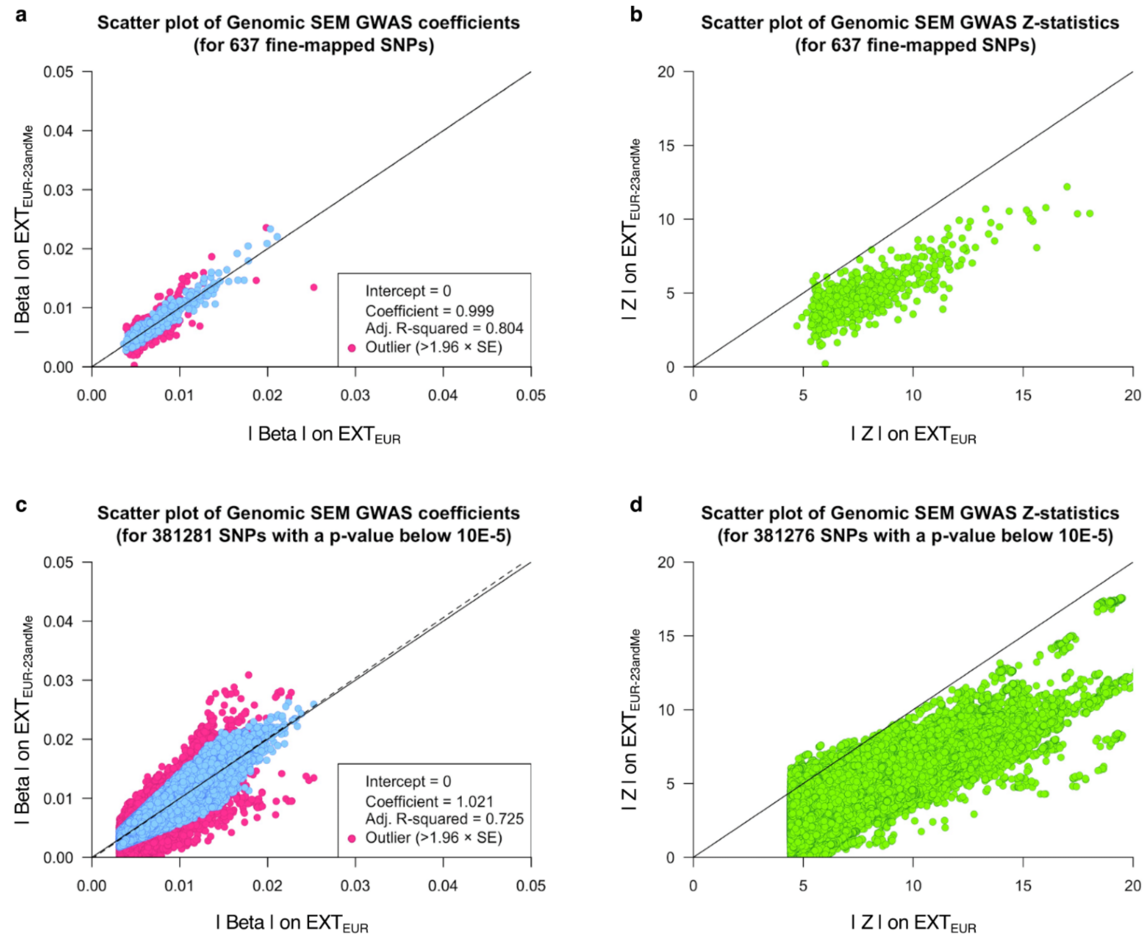


***Figure S6. Scatter plots of multivariate Genomic SEM GWAS coefficients and Z statistics of a subset of SNPs.*** The left panel displays the absolute value of the GWAS coefficients ("Beta") of the multivariate GWAS on externalizing ($\text{EXT}_{\text{EUR}}$) against their estimates from the analogous down-sampled analysis ($\text{EXT}_{\text{EUR-23andMe}}$), with corresponding *Z* statistics in the right panel. Because the Z-statistic is a function of both sample size and the size of the GWAS coefficient, general attenuation of the *Z* statistic is to be expected when down-sampling. This attenuation is noticeable by visual inspection of the scatter plots: the green dots for the *Z* statistics fall systematically below the diagonal line, while the blue dots for the coefficients (for which we expect little to no attenuation) are dispersed around the diagonal, suggesting concordance in coefficients. In panels **a** and **c**, there are a total of 107 (out of 637) and 69,123 (out of 381,281) outliers for which the GWAS coefficient fell outside the 95% confidence interval of the full-data estimate. Panels **a** and **c** report the results of a linear regression analysis of the observations in the figure. A diagonal solid line with a slope of 1 and an intercept of 0 is plotted for reference. Dashed lines represent fitted regression lines.

### 5. Replication

**Section authors:** Camille M. Williams, Justin D. Tubbs, Yuchen Ning, & Travis T. Mallard

#### 5.1. Introduction

In this section, we report a series of quasi-replication analyses similar to those described in EXT1[1], but aligned to a multi-ancestry context. The primary goal is to evaluate whether prioritized variants (here, the fine-mapped putative causal SNPs) show evidence of association in an independent sample withheld from discovery-stage analyses. This strategy enhances statistical power for replication by explicitly focusing on the most robust and relevant GWAS signals, thereby reducing the multiple testing burden. It is particularly useful when equally large, independent GWAS for the target trait are unavailable, or when direct replication is complicated by phenotype availability.

Here, we tested whether statistical signals in the fine-mapped variants—the SNPs with putative causal effects across ancestral groups—replicated in a multi-ancestry GWAS of externalizing disorders in the All of Us Research Program [52,53](AoU v8). To accomplish this, we (i) conducted ancestry-specific confirmatory factor analyses of electronic health record (EHR) defined externalizing disorders in AoU; (ii) performed genome-wide association analyses of the resultant externalizing factor scores within each ancestry; (iii) evaluated the genetic correlation between ancestries and cohorts (i.e., discovery and replication); (iv) meta-analyzed AoU GWAS results across ancestry groups; and (iv) performed a series of analyses to evaluate the replication record, including tests of joint enrichment, sign concordance, nominal significance, and Bonferroni-adjusted significance. Throughout this section, we refer to the putative causal SNPs fine-mapped via SuSiEx as the discovery set, and the randomly sampled LD-pruned and variance-matched SNPs as the background set.

#### 5.2. Methodology

We selected two sets of SNPs that were strongly associated with externalizing based on PIPs from SuSiEx[46]: 647 putatively causal SNPs with a PIP ≥ 0.5 (moderate confidence), and 214 putatively causal SNPs with a PIP ≥ 0.95 (high confidence). After excluding $Q_{SNP}$ hits, these PIP ≥ 0.50 and PIP ≥ 0.95 discovery SNP sets were reduced to 637 and 209 SNPs, respectively. We aligned the $EXT_{META}$ GWAS results to the 1000 Genomes Project Phase 3 v5 European and

African super populations[21] and then subset the $EXT_{META}$ GWAS to each of these discovery SNP sets to conduct quasi-replication analyses separately for SNPs with moderate to high confidence of causality.

To evaluate whether these SNP sets were robustly associated with clinical conditions related to externalizing, we conducted a GWAS of externalizing disorders. Specifically, we focused on clinical conditions on the externalizing spectrum in the AoU dataset to further investigate the clinical relevance and potential utility of externalizing SNP associations.

The AoU is a longitudinal cohort study in the U.S., launched in 2015, with health and genetic data from ~450,000 diverse participants. It includes survey data collected by the program (e.g., demographics and physical measurements), EHR data, and biospecimen data (e.g., genomics). Clinical conditions were defined using ICD-9/10 codes for participants with EHR data:

- Alcohol use disorder (AUD): ICD-9 (303, 303.0, 303.00, 303.01, 303.02, 303.03, 303.9, 303.90, 303.91, 303.92, 303.93, 305.0, 305.00, 305.01, 305.02, 305.03) and ICD-10 (F10, F10.1, F10.10, F10.11, F10.12, F10.120, F10.121, F10.129, F10.13, F10.130, F10.131, F10.132, F10.139, F10.14, F10.15, F10.150, F10.151, F10.159, F10.18, F10.180, F10.181, F10.182, F10.188, F10.19, F10.2, F10.20, F10.21, F10.22, F10.220, F10.221, F10.229, F10.23, F10.230, F10.231, F10.232, F10.239, F10.24, F10.25, F10.250, F10.251, F10.259, F10.26, F10.27, F10.28, F10.280, F10.281, F10.282, F10.288, F10.29)
- Antisocial personality disorder (ASPD): ICD-9 (301.7) and ICD-10 (F60.2)
- Attention-deficit/hyperactivity disorder (ADHD): ICD-9 (314, 314.0, 314.00, 314.01, 314.1, 314.2, 314.8, 314.9) and ICD-10 (F90, F90.0, F90.1, F90.2, F90.8, F90.9)
- Borderline personality disorder (BPD): ICD-9 (301.83, 301.3) and ICD-10 (F60.3)
- Cannabis use disorder (CUD): ICD-9 (304.3, 304.30, 304.31, 304.32, 304.33, 305.2, 305.20, 305.21, 305.22, 305.23) and ICD-10 (F12, F12.1, F12.10, F12.11, F12.12, F12.120, F12.121, F12.122, F12.129, F12.13, F12.15, F12.150, F12.151, F12.159, F12.18, F12.180, F12.188, F12.19, F12.2, F12.20, F12.21, F12.22, F12.220, F12.221, F12.222, F12.229, F12.23, F12.25, F12.250, F12.251, F12.259, F12.28, F12.280, F12.288, F12.29)
- Cocaine use disorder (CocUD): ICD-9 (304.2, 304.20, 304.21, 304.22, 304.23, 305.6, 305.60, 305.61, 305.62, 305.63) and ICD-10 (F14, F14.1, F14.10, F14.11, F14.12, F14.120, F14.121, F14.122, F14.129, F14.13, F14.14, F14.15, F14.150, F14.151, F14.159, F14.18, F14.180, F14.181, F14.182, F14.188, F14.19, F14.2, F14.20, F14.21, F14.22,

F14.220, F14.221, F14.222, F14.229, F14.23, F14.24, F14.25, F14.250, F14.251, F14.259, F14.28, F14.280, F14.281, F14.282, F14.288, F14.29)

- Conduct disorder (CD): ICD-9 (312, 312.0, 312.00, 312.01, 312.02, 312.03, 312.1, 312.10, 312.11, 312.12, 312.13, 312.2, 312.20, 312.21, 312.22, 312.23, 312.4, 312.8, 312.81, 312.82, 312.89, 312.9) and ICD-10 (F91.0, F91.1, F91.2, F91.8, F91.9)
- Gambling disorder (GD): ICD-9 (312.31) and ICD-10 (F63.0)
- Histrionic personality disorder (HPD): ICD-9 (301.5, 301.50, 301.59) and ICD-10 (F60.4)
- Intermittent explosive disorder (IED): ICD-9 (312.34, 312.35) and ICD-10 (F63.81)
- Narcissistic personality disorder (NPD): ICD-9 (301.81) and ICD-10 (F60.81)
- Opioid use disorder (OUD): ICD-9 (304.0, 304.00, 304.01, 304.02, 304.03, 305.5, 305.50, 305.51, 305.52, 305.53) and ICD-10 (F11, F11.1, F11.10, F11.11, F11.12, F11.120, F11.121, F11.122, F11.129, F11.13, F11.14, F11.15, F11.150, F11.151, F11.159, F11.18, F11.181, F11.182, F11.188, F11.19, F11.2, F11.20, F11.21, F11.22, F11.220, F11.221, F11.222, F11.229, F11.23, F11.24, F11.25, F11.250, F11.251, F11.259, F11.28, F11.281, F11.282, F11.288, F11.29)
- Oppositional defiant disorder (ODD): ICD-9 (313.81) and ICD-10 (F91.3)
- Tobacco use disorder (TUD): ICD-9 (305.1) and ICD-10 (F17, F17.2, F17.20, F17.200, F17.201, F17.203, F17.208, F17.209, F17.21, F17.210, F17.211, F17.213, F17.218, F17.219, F17.22, F17.220, F17.221, F17.223, F17.228, F17.229, F17.29, F17.290, F17.291, F17.293, F17.298, F17.299)

Cases were defined as individuals with at least one relevant ICD-9 or ICD-10 code to maximize statistical power for gene discovery, and control subjects were defined as individuals who lacked the ICD-9/10 code corresponding to the target disorder. We restricted the sample to participants with reported sex that matched their genetically inferred sex, who had available birth year data, who self-identified as either “White” or “Black or African American”, and who were genetically most similar to either the African or the European 1000 Genome Project superpopulation. Inferred ancestry labels were generated by the All of Us Research Program Genomics Investigators and described in detail in a flagship publication[53]. Briefly, a random forest classifier was trained using data from the Human Genome Diversity Project and 1000 Genomes reference panels. This classifier was subsequently applied to AoU participants, showing high concordance between inferred ancestry and self-reported ethnicity. Participants whose self-reported race did not match their genetically inferred ancestry were excluded.

Due to the limited number of cases ($N$ < 100) for some of the clinical conditions, conducting univariate GWASs of these disorders to input in a multivariate GWAS of externalizing in Genomic SEM was not feasible. Instead, we estimated externalizing factor scores based on these externalizing spectrum disorders and conducted a GWAS of these scores. In light of the small number of cases for several personality and disruptive behavior disorders, particularly among participants from AFR-like ancestry (**Supplementary Table 12)**, we combined CD, ODD, and IED into a single disruptive behavior disorder (DBD) variable, and ASPD, BPD, HPD, and NPD into a single personality disorder variable reflecting cluster B personality disorders. The gambling disorders phenotype had fewer than 100 cases across ancestry groups and was excluded from the factor analyses.

We used the *lavaan* package[54] v0.6-20 to conduct confirmatory factor analyses and estimate factor scores for 201,904 EUR-like and 61,975 AFR-like individuals. Specifically, we used weighted least squares means and variance adjusted (WLSMV) estimation to fit ancestry-specific factor models where all eight clinical indicators loaded onto a common externalizing factor. The WLSMV estimator uses diagonally weighted least squares to estimate the model parameters and the full weight matrix to compute robust standard errors, and a mean- and variance-adjusted test statistic. We assessed model fit on the basis of the CFI, the Tucker-Lewis index (TLI), the SRMR, and the root mean square error of approximation (RMSEA), with CFI and TLI ≥ .95, SRMR ≤ 0.08, and RMSEA ≤0.05 values indicative of good fit[55]. We computed ancestry-specific factor scores using the Empirical Bayes Modal (EBM) approach with the *lavPredict* function. The EBM method weights indicators based on factor loadings and measurement error and then combines these weighted observations with a Bayesian prior on the latent factors to create factor scores. The results of the confirmatory factor analysis are reported in **Supplementary Table 12**.

GWASs on the externalizing factor in AoU ($EXT_{AoU}$) were performed separately in EUR- and AFR-like samples. First, subjects were excluded from analysis if the AoU Genomics Investigators flagged them as outliers on quality-control metrics or if their self-reported sex was inconsistent with their genetically inferred sex. Data were further restricted to the maximally independent set of unrelated samples, as provided by the AoU Genomics Investigators. Subsequently, within-sample ancestry-specific genetic principal components (PCs) were calculated for EUR- and AFR-like participants using PLINK[26] v2.0.0-a.6.12. Linear association tests were performed using PLINK v2.00a5.11LM for each SNP available in the Allele Count/Allele Frequency (ACAF) threshold callset provided by AoU with an ancestry-specific minor allele frequency greater than 0.001. Covariates included sex, year of birth (YOB), $YOB^2$, sex*YOB, and the first 10 ancestry-specific PCs. This resulted in 201,904 EUR-like and 61,974 AFR-like individuals with quality-controlled genomic data and complete phenotypic data (including covariates). Following genome-wide association analyses, SNP-level results were lifted over from CRCh38 to GRCh37 using R, with > 99.99% of SNPs successfully transferred.

Prior to meta-analyzing the $EXT_{AoU}$ GWAS results, we estimated the ancestry-specific genetic correlations between each $EXT_{AoU}$ GWAS and the discovery EXT factors using Genomic SEM[23]. Briefly, we found that the externalizing factor score was highly genetically correlated with the $EXT_{AoU}$ GWAS in both EUR-like individuals ($r_g$ = 0.840, s.e. = 0.025) and AFR-like individuals ($r_g$ = 0.899, s.e. = 0.142). Given these findings, we proceeded with our quasi-replication analyses and conducted a fixed-effect sample-size-weighted meta-analysis of the EUR- and AFR-like $EXT_{AoU}$ GWASs using METAL v2011-03-25[22].

Using PLINK v2.0.0-a.6.12, we generated a set of LD-pruned SNPs from the 1000 Genomes Project Phase 3 v5 European and African super populations[21]. SNPs present in both populations were jointly pruned to remove variants in high linkage disequilibrium across the combined sample, followed by separate re-pruning within each population to eliminate residual population-specific LD. We prepared background SNP sets by subsetting the $EXT_{AoU}$ GWAS to the LD-pruned set and removing all fine-mapped variants. For each fine-mapped SNP, we then sampled 250 background SNPs matched on effective genotype variance estimated from the $EXT_{AoU}$ GWAS, allowing an initial tolerance of 0.02 that could be expanded up to three times if insufficient matches were available. To compute the effective genotype variance for each SNP, we first estimated the genotype variance within each ancestry as 2p(1−p), where p is the effect allele frequency in $EXT_{AoU}$ GWAS samples, and then calculated a sample-size–weighted average of the genotype variance across the two ancestries. Sampling was performed on SNPs with adequate sample size ($N \geq 80\%$ of the maximum $N$ in the GWAS) were included, ensuring that differences in association signals were not driven by allele frequency or imputation quality. The background PIP ≥ 0.50 SNP set included 64,387 SNPs, and the background PIP ≥ 0.95 SNP set included 64,395 SNPs.

To evaluate whether fine-mapped SNPs showed stronger associations in AoU than expected by chance, we conducted one-sided non-parametric Mann–Whitney U tests (Wilcoxon rank-sum tests) using the *wilcox.test* function from the stats R package[56]. This test compares the distribution of *P* values for the fine-mapped SNPs to those from randomly sampled background SNPs in order to assess whether fine-mapped SNPs are more strongly associated (i.e., have smaller *P* values).

We used the *binom.test* function from the stats R package[56] to perform a binomial sign concordance test. This test compares the directions of the *Z* statistics for the fine-mapped SNPs from the $EXT_{META}$ GWAS with those from the $EXT_{AoU}$ GWAS. Under the null hypothesis, we expect approximately 50% of SNPs to show concordant effect directions by chance. A significant result ($P \leq 0.05$) indicates that the direction of effects is more concordant between the $EXT_{AoU}$ GWAS and the discovery SNP sets, providing support for replication.

To rule out the scenario that sign concordance is driven by a large number of non-significant Z-statistics, we tested the significance of the discovery hit sets in the $EXT_{AoU}$ GWAS. We extracted *P* values from the discovery SNPs in the $EXT_{AoU}$ GWAS and assessed whether a larger-than-empirically-expected proportion (determined using the randomly sampled *P* values) of these SNPs reached nominal significance ($P \leq 0.05$) as well as significance after Bonferroni correction ($P \leq 0.05$/number of SNPs in discovery set).

#### 5.3. Results

We modeled a latent phenotypic externalizing factor using ICD-9/10 diagnoses from electronic health data in AoU for individuals with EUR-like (N = 201,904) and AFR-like (N = 61,975) ancestry with genetic data (**Supplementary Table 12)**. The single externalizing factor model fit the data well in the EUR-like ($\chi^2(20) = 690.099$, CFI = 0.994, TLI = 0.992, RMSEA = 0.013, SRMR = 0.061) and AFR-like ($\chi^2(20) = 484.933$, CFI = 0.994, TLI = 0.992, RMSEA = 0.019, SRMR = 0.089) ancestry groups. Factor parameters were similar across ancestry groups (*r* = 0.953, s.e. = 0.063). Loadings ranged from 0.299 (AFR-like) and 0.412 (EUR-like) for ADHD to 0.897 (AFR-like) and 0.904 (EUR-like) for Cocaine Use Disorder.

Three fine-mapped SNPs from the discovery sets were missing from the $EXT_{AoU}$ GWAS results: one in the PIP ≥ 0.50 set, and two in the PIP ≥ 0.95 set (**Supplemental Table 13)**. As the number of missing fine-mapped variants was small (i.e., one with a PIP ≥ 0.5 and two with a PIP

≥ 0.95), we elected to forgo LD-proxy substitution, ensuring that replication analyses were explicitly based on putative causal SNPs.

The 634 moderate-confidence putatively causal SNPs from the $EXT_{META}$ GWAS showed strong evidence of replication across all tests (**Supplemental Table 13)**. The Mann–Whitney test rejected the null hypothesis of no enrichment (one-sided $P$ = 2.58e-55). The sign concordance test revealed that 547 out of 634 SNPs (86%) had the same direction of effect in the $EXT_{AoU}$ GWAS (one-sided $P$ = 1.96e-82). The nominal significance test showed that 212 SNPs (33%) reached nominal significance (one-sided $P$ = 1.83e-84), and 15 SNPs (2%) were significant after Bonferroni correction ($P$ = 1.77e-23), with both results exceeding the expected proportion under the null hypothesis.

Strong replication was also observed for the 207 high-confidence putatively causal SNPs from the $EXT_{META}$ GWAS across all tests (**Supplemental Tables 13)**. The Mann–Whitney test rejected the null hypothesis of no enrichment (one-sided $P$ = 9.26e-24). The sign concordance test revealed that 183 out of 207 SNPs (88%) had the same direction of effect in the $EXT_{AoU}$ GWAS (one-sided $P$ = 1.72e-31). The nominal significance test showed that 75 SNPs (36%) reached nominal significance (one-sided $P$ = 1.10e-84), and 9 SNPs (4%) were significant after Bonferroni correction ($P$ = 2.31e-12), with both results exceeding the expected proportion under the null hypothesis.

### 6. Bioannotation and bioinformatic analyses

**Section authors**: Camille M. Williams, K. Paige Harden, & Travis T. Mallard

#### 6.1. Introduction

To contextualize the polygenic signals associated with externalizing, we implemented an ancestry-aware bioannotation and bioinformatics pipeline to identify relevant tissues, developmental epochs, cell types, and molecular pathways. Briefly, we leveraged gene-based association statistics from MAGMA[47] (**Supplementary Section 4.4**), prioritized genes from FLAMES[48] (**Supplementary Section 4.4**), and complementary multi-omic resources spanning spatial, temporal, and cellular resolution. These inputs supported a set of genome-wide and prioritized-gene analyses within the MAGMA, SynGO[57], and DRUGSETS[58] frameworks (**Figure S7**).

Throughout, we prioritize robust, generalizable inference and therefore report results informed by multiple ancestries. Where informative, we also report ancestry-specific results from the externalizing GWAS in EUR-like and AFR-like populations, as well as a re-analysis of the EXT1 GWAS[1] to enable commensurable comparison to our prior work.

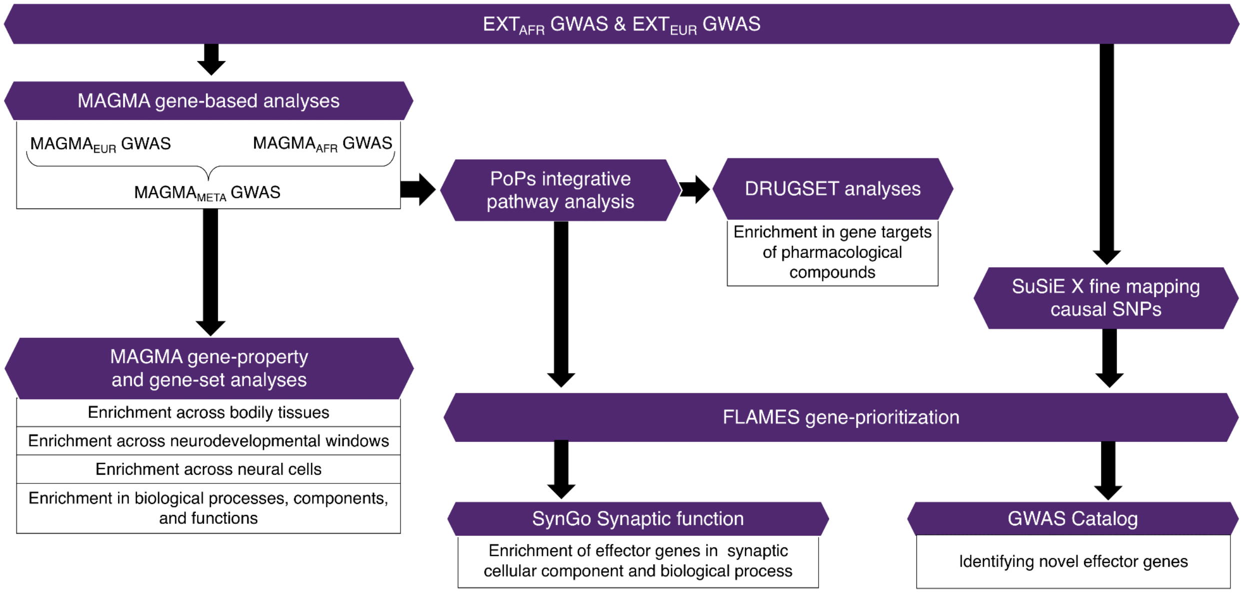


***Figure S7. Overview of bioannotation and bioinformatic analyses.*** European (EUR) and African (AFR) ancestry-specific GWAS served as input for cross-ancestry fine-mapping with SuSiEx and ancestry-specific gene-level analyses with MAGMA. MAGMA gene-based results were meta-analyzed to generate a cross-ancestry MAGMA META GWAS (**Supplementary Section 4.4**), which was subsequently used for gene property and gene-set enrichment analyses, as well as the integrative pathway analysis with PoPS. PoPS results were used as input in the DRUGSET analyses (**Supplementary Sections 6.2.1 and 6.3.1**). FLAMES combined fine-mapping, integrative pathway, and tissue-specific gene property results to prioritize causal "effector" genes for externalizing across ancestries (**Supplementary Section 4.4**). Effector genes were subsequently analyzed using SynGO to examine synaptic enrichment (**Supplementary Sections 6.2.2 and 6.3.2**), and the GWAS Catalog was used to evaluate whether these genes had previously been associated with externalizing-related phenotypes (**Supplementary Section 4.5**).

### 6.2. Methods

#### 6.2.1. Bioinformatic characterization of genome-wide architecture

To interrogate the biological underpinnings of externalizing, we used MAGMA[47] (v1.10) to conduct a series of gene property and gene-set enrichment analyses based on the MAGMA-derived gene-level test statistics (**Supplementary Section 4.4**). These analyses were designed to determine when, where, and in which cellular contexts externalizing-linked genes tended to be expressed, providing insight into their functional biology. Below, we concisely describe each analysis, as well as the relevant data.

***Enrichment across bodily tissues.*** We performed a gene property analysis to assess the relationship between polygenic signal for externalizing and 54 tissue-specific gene expression profiles measured in GTEx v8[59] (obtained via FUMA[60]; https://fuma.ctglab.nl/home). For each tissue, we tested for enrichment while including the gene's mean expression across all tissues as a covariate (in addition to default covariates). We applied a Bonferroni correction to account for multiple testing across the 54 tissues.

***Enrichment across neurodevelopmental windows.*** We conducted MAGMA gene property analyses with BrainSpan[61] data to assess whether the polygenic signal for externalizing was associated with gene expression during specific neurodevelopmental windows. BrainSpan provides spatiotemporal expression profiles across 11 defined developmental windows. For each window, we tested for enrichment while including each gene's mean expression across all windows as a covariate (in addition to MAGMA's default gene covariates); we also report results without this adjustment. We applied a Bonferroni correction to account for multiple testing across the 11 windows.

***Enrichment across neural cells.*** We conducted MAGMA gene property analyses across six human brain single-cell/single-nucleus RNA sequencing (sc/snRNA-seq) datasets spanning early prenatal development through older adulthood, testing whether polygenic signal for externalizing was associated with cell-resolved expression patterns in brain tissues. The gene expression profiles were prepared by the FUMA development team[62] (https://github.com/tanyaphung/scrnaseq_viewer) and are succinctly described below. For each dataset, we tested for enrichment while including an additional covariate for each gene's mean expression across all cell types in that dataset. We applied a Bonferroni correction to account for multiple testing within each of the six datasets.

*Early prenatal (whole brain).* We selected a scRNA-seq dataset with single-cell transcriptomic profiles collected from 26 brain specimens spanning 5–14 weeks post-conception, dissected into 111 distinct biological samples[63]. This dataset includes one level of cell type granularity across 10 regions (brain, cerebellum, diencephalon, forebrain, head, hindbrain, medulla, midbrain, pons, and telencephalon), grouped into 11 functional classes, including glioblasts, oligodendrocytes, radial glial cells, neurons, neuroblasts (labeled sensu vertebrata), neuronal intermediate progenitor cells (IPCs) (labeled sensu nematoda and protostomia), erythrocytes, macrophages, fibroblasts, neuroplacodal cells, and neural crest cells.

*Early prenatal (embryonic midbrain).* We analyzed scRNA-seq data from the Linnarsson GSE76381 dataset[64], which profiles human embryonic ventral midbrain cells collected between 6–11 weeks of gestation. The dataset includes a wide range of neuronal and non-neuronal cell types, such as the oculomotor and trochlear nucleus, serotonergic neurons, medial neuroblasts, dopaminergic neuroblasts, dopaminergic neurons, red nucleus, GABAergic neurons, lateral neuroblasts, mediolateral neuroblasts, neuronal progenitors, and progenitor cells from the medial floorplate, lateral floorplate, midline, and basal plate. It also includes radial glia-like cells, microglia, endothelial cells, pericytes, ependymal cells, and oligodendrocyte precursor cells.

*Mid-to-late prenatal.* We selected a scRNA-seq dataset with single-cell transcriptomic profiles from 14–25 weeks post-conception, sampled in 13 individuals[65]. This dataset features two levels of cell-type granularity across six neocortical areas (parietal cortex, prefrontal cortex, primary motor cortex, primary somatosensory cortex, primary visual cortex, and temporal cortex). At the first level of granularity (Level 1), cell types are grouped into 10 major functional classes: radial glial cells, GABAergic interneurons, microglial cells, progenitor cells, cerebral endothelial cells, blood vessel endothelial cells, oligodendrocyte precursor cells, Cajal-Retzius cells, glutamatergic neurons, and unknown cells. At Level 2, these classes are further divided into 209 transcriptionally distinct subtypes.

*First trimester to adolescence.* We analyzed single-nucleus chromatin accessibility and transcriptomic data from 38 human neocortical samples, spanning the first trimester to adolescence[66]. Samples were taken from multiple regions, including the prefrontal cortex; the primary visual cortex; Brodmann areas 9, 10, and 17; the neocortex; telencephalon; and forebrain. Cell types were classified at three levels of granularity. At the broader level (Level 1), cells were grouped into six major classes: glia, immune, neuron, progenitor, vascular, and unknown. These classes were further divided at Level 2 into 12 subtypes, including astrocytes, Cajal-Retzius cells, GABAergic neurons, glutamatergic neurons, excitatory neuron (EN) IPCs, glial IPCs, microglia, oligodendrocytes, oligodendrocyte precursor cells (OPCs), radial glia, vascular cells, and unknown cells. These subtypes were further refined into 29 more specific categories (Level 3), such as distinct subtypes of GABAergic and glutamatergic neurons.

*Adulthood (grey matter)*. We analyzed snRNA-seq data from postmortem brain tissue collected from three human donors aged 18 to 68 years, with no known history of neuropsychiatric or neurological conditions[67]. Tissues were enriched for neurons from approximately 100 anatomically defined locations across the cerebral cortex, hippocampus, cerebral nuclei, hypothalamus, thalamus, midbrain, pons, cerebellum, medulla, and spinal cord. Cell types were annotated at two levels of granularity. At the broadest level (Level 1), there were 13 major cell classes, including neurons, astrocytes, Bergmann glial cells, CNS macrophages, choroid plexus epithelial cells, endothelial cells, ependymal cells, fibroblasts, leukocytes, oligodendrocytes, oligodendrocyte precursor cells, pericytes, and vascular-associated smooth muscle cells. At Level 2, these classes were further divided into 31 more specific regional subtypes (e.g., mammillary body cells, hippocampal dentate gyrus cells).

*Middle and older adulthood (white matter).* We analyzed snRNA-seq data from post-mortem white matter samples of 20 human donors (10 young adults aged 30–45 years and 10 older adults aged 60–75 years[68]). Tissues were collected from the primary motor cortex (Brodmann area 4, BA4), the arbor vitae of the cerebellum (CB), and the cervical spinal cord (CSC), including the fasciculi cuneatus and gracilis. Cell types were annotated at three levels of granularity. At the broader level (Level 1), classes included astrocytes, endothelial/pericyte cells, microglia/macrophages, neurons, excitatory neurons, inhibitory neurons, RELN+ neurons, oligodendrocytes, oligodendrocyte precursor cells, and unidentified cells. At Level 2, these ten categories were further resolved into 15 subtypes, including astrocytes, capillary endothelial cells, CNS macrophages, cerebellar granule cells, committed oligodendrocytes, arterial endothelial cells, GABAergic neurons, glutamatergic neurons, leukocytes, microglial cells, mural cells, neurons, oligodendrocytes, oligodendrocyte precursor cells, and vascular-associated smooth muscle cells. These subtypes were further classified into 60 categories of cell types at Level 3, including specific types of excitatory and inhibitory neurons.

***Enrichment in biological processes, components, and functions***. We used a competitive gene-set enrichment analysis to test whether MAGMA-derived gene-level signals for externalizing were enriched in ontological gene sets. Specifically, we used the 2025 C5 Gene Ontology (GO) classifications from the Molecular Signatures Database[69] (MSigDB). These 10,480 C5 GO gene sets reflect various biological processes, molecular functions, and cellular components linked to gene products. Analyses employed MAGMA's default gene covariates (e.g., gene size, SNP density). Statistical significance was assessed using a Bonferroni correction across the 10,480 gene sets tested.

***Enrichment in gene targets of pharmacological compounds.*** We used DRUGSETS[58] to test whether the polygenic signal for externalizing was enriched among the protein targets of approved or investigational medications. Specifically, we applied an adjusted version of DRUGSETS, which implements MAGMA's competitive gene-set framework to test whether medications whose protein targets (or well-curated interactors) are enriched in a given GWAS. In this adjusted version, MAGMA $Z$ statistics were replaced with PoPS[49] scores, a gene-level metric that integrates GWAS associations with functional genomic features to prioritize genes.

DRUGSETS tests for enrichment in 1,201 compound-level drug-gene sets based on known drug-gene interactions in the DGIdb[70] and the Drug Repurposing Hub[71]. For each drug, we ran competitive gene-set analyses on the $EXT_{META}$, $EXT_{EUR}$, and $EXT_{AFR}$ results, conditioning on the union of all drug-target genes to account for properties shared across pharmacological targets. Statistical significance was evaluated with Bonferroni thresholds reflecting the number of drug gene sets tested in each analysis: 1,171 for $EXT_{META}$ ($P \leq 0.05/1{,}171 = 4.23e\text{-}5$), 1,163 for $EXT_{EUR}$ ($P \leq 0.05/1{,}163 = 4.29e\text{-}5$), and 1,169 for $EXT_{AFR}$ ($P \leq 0.05/1{,}169 = 4.27e\text{-}5$).

To contextualize compound-level signals, we used DRUGSETS' built-in grouping function to aggregate drug-level results by Anatomical Therapeutic Chemical (ATC) level III class, clinical indication, and mechanism of action. The ATC Level III class is a standardized system dividing drugs by their primary therapeutic or pharmacological class at the third specificity level. The clinical indication refers to the primary medical use or prescribed reason for each drug, and the mechanism of action corresponds to the molecular or cellular mechanism through which the drug exerts its effect. Within each grouping, a multiple linear regression model was fit with the drug-level gene-set *t* statistic as the outcome and indicators for group membership as predictors (restricting to groups with ≥ 5 compounds). A Bonferroni correction was used to account for multiple testing within each grouping (ATC: $P \leq 5.56e\text{-}4$; indication: $P \leq 4.24e\text{-}4$; MOA: $P \leq 6.33e\text{-}4$).

#### 6.2.2. Bioinformatic characterization of effector genes

We used SynGO[57] release 1.2—an evidence-based, expert-curated ontology focused specifically on synaptic structure and function—to characterize synaptic biology linked to the effector genes prioritized by FLAMES. In contrast to broad GO collections, SynGO provides granular, synapse-focused annotations supported by multiple forms of evidence. Here, we tested whether prioritized genes were over-represented in synaptic cellular component and biological process gene sets, using brain-expressed genes as the background set and applying a high stringency evidence filter. Importantly, to ensure that our results were not contaminated by data leakage (i.e., non-independence of training and testing data), we used the FLAMES results based on pathway-naïve PoPS scores. For a term to be considered, we required that a minimum of three genes overlap with the annotation. Statistical significance was set as a Benjamini-Hochberg false discovery rate (FDR) of 1% (the default for SynGO).

### 6.3. Results

#### 6.3.1. Bioinformatic characterization of genome-wide architecture

***Enrichment across bodily tissues.*** Of the 54 tissues available in the GTEx v8 dataset, the polygenic signal of $EXT_{META}$ was enriched in 14 central nervous system tissues: the amygdala, anterior cingulate cortex, basal ganglia, brain cortex, cerebellar hemisphere, cerebellum, B9 frontal cortex, hippocampus, hypothalamus, nucleus accumbens, caudate and putamen (basal ganglia), spinal cord, substantia nigra, and pituitary. This pattern was observed with and without adjustment for average gene expression across all tissues. We observed the same enrichment pattern for both $EXT_{EUR}$ (correlation of coefficients: $r = .999$, s.e. = 0.007) and EXT1 (correlation of coefficients: $r = .988$, s.e. = 0.021). Enrichment was limited for $EXT_{AFR}$, with significant signals restricted to the hypothalamus and substantia nigra (**Supplementary Table 15**), though this presumably reflects limited statistical power as opposed to regional specificity.

***Enrichment across neurodevelopmental windows.*** When not adjusting for mean gene expression across windows, we observed significant enrichment for $EXT_{META}$ and $EXT_{EUR}$ across all 11 windows in the BrainSpan data (**Supplemental Table 16**). For both $EXT_{META}$ and $EXT_{EUR}$, some of the strongest signals were observed during the early mid-prenatal ($P_{META}$ = 8.43e-17, $P_{EUR}$ = 7.27e-21) and late mid-prenatal ($P_{META}$ = 2.21e-18, $P_{EUR}$ = 6.34e-20) windows. There were no significant enrichment patterns for $EXT_{AFR}$ after correcting for multiple testing.

After including gene-level average expression as a covariate, we found that the polygenic signal for externalizing was only enriched during early prenatal ($P_{META}$ = 1.85e-5, $P_{EUR}$ = 4.94e-6), early mid-prenatal ($P_{META}$ = 1.20e-7, $P_{EUR}$ = 2.90e-8, $P_{AFR}$ = 1.73e-3), and late mid-prenatal ($P_{META}$ = 2.15e-7, $P_{EUR}$ = 2.02e-8, $P_{AFR}$ = 5.15e-4) windows. Patterns of enrichment were very similar between $EXT_{META}$, $EXT_{EUR}$, and EXT1.

***Enrichment across neural cells.*** With respect to cell-type expression profiles, we observed widespread enrichment of polygenic signal for externalizing in neurons—especially GABAergic and glutamatergic neurons—and in oligodendrocyte precursor cells beginning very early in development (early post-conception) and persisting through adulthood. Generally, these cell-specific patterns did not exhibit much regional or spatial specificity, as enrichment patterns tended to be broadly similar throughout the brain. While analyses were conducted across multiple scales of cellular organization, we focus here on the resolution within each dataset that maximized biological interpretability on a systems-level scale. Specifically, we describe the sc/snRNA-seq enrichment results across neurodevelopmental windows and brain regions, spannning early prenatal brain development[63] (Level 1), including the embryonic midbrain[64]; mid-to-late prenatal brain development[65](Level 1); brain development from the first trimester to adolescence[66] (Level 2); adulthood grey matter[67] (Level 1), and white matter[68] (Level 2). Below, we report the results for $EXT_{META}$. Full results are reported after Bonferroni correction across all cell types within each dataset (**Supplementary Tables 17–22**).

*Early prenatal (whole brain).* Between 5.5–15 weeks post-conception, the polygenic signal for externalizing showed the most temporally and regionally consistent pattern of enrichment in neurons and vertebrate neuroblasts (**Figure S8**). Vertebrate neuroblasts are a distinct class of progenitor cells that can proliferate and differentiate into a variety of neuronal and glial cell types. Enrichment of externalizing liability in other cell types during this early developmental period was more localized and time-specific. For example, signal was only enriched in oligodendrocytes at 12 weeks post-conception, albeit across multiple brain regions (**Supplementary Table 17**).

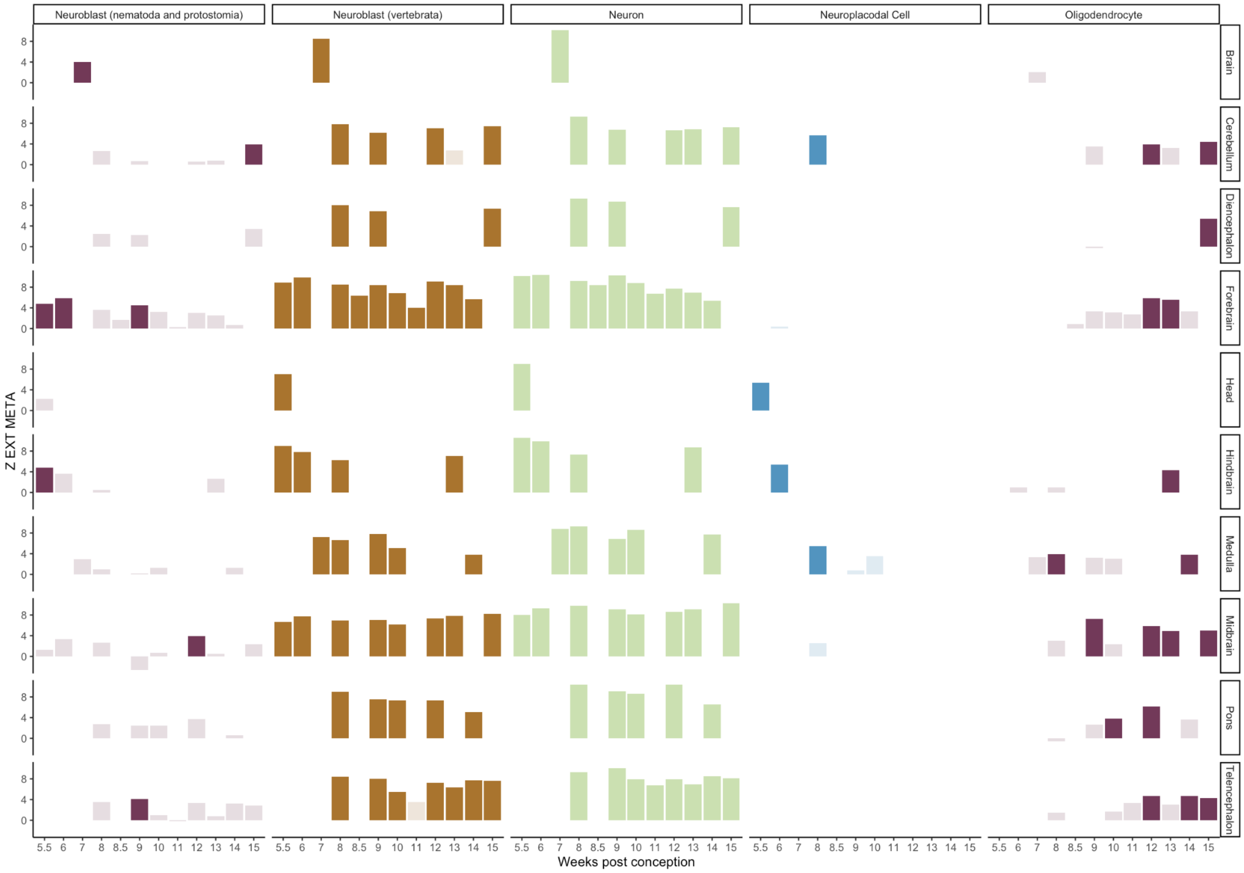


***Figure S8. Enrichment of externalizing liability in specific cell types across brain regions during early prenatal brain development.*** The $y$-axis reflects the $Z$ statistic from the enrichment test. Darker shades indicate associations that remain significant after Bonferroni correction. Only cell types with at least one significant association are shown.

*Early prenatal (embryonic midbrain)*. Focusing specifically on the ventral midbrain cells collected between 6- and 11-weeks of gestation, externalizing liability was significantly enriched in dopaminergic neurons (DA0–DA2), serotonergic neurons (Sert), GABAergic neurons (Gaba), GABAergic neuroblasts (NbGaba), mediolateral neuroblasts (NbML5), oculomotor and trochlear nucleus cells (OMTN), and the red nucleus (RN) (**Supplementary Table 18**).

*Mid-to-late prenatal.* From 14 to 25 weeks post-conception, externalizing liability was significantly in glutamatergic neurons across multiple brain regions (**Figure S9**;

Supplementary Table 19**). In contrast, GABAergic interneurons and progenitor cells exhibited more restricted temporal and spatial enrichment patterns, visible at 14 or 16 weeks in a few regions and then re-emerging at 22 or 25 weeks. Other enrichment patterns displayed more temporal and spatial specificity. For instance, forebrain radial glial cells were only enriched in visual cortical regions at 17 weeks.

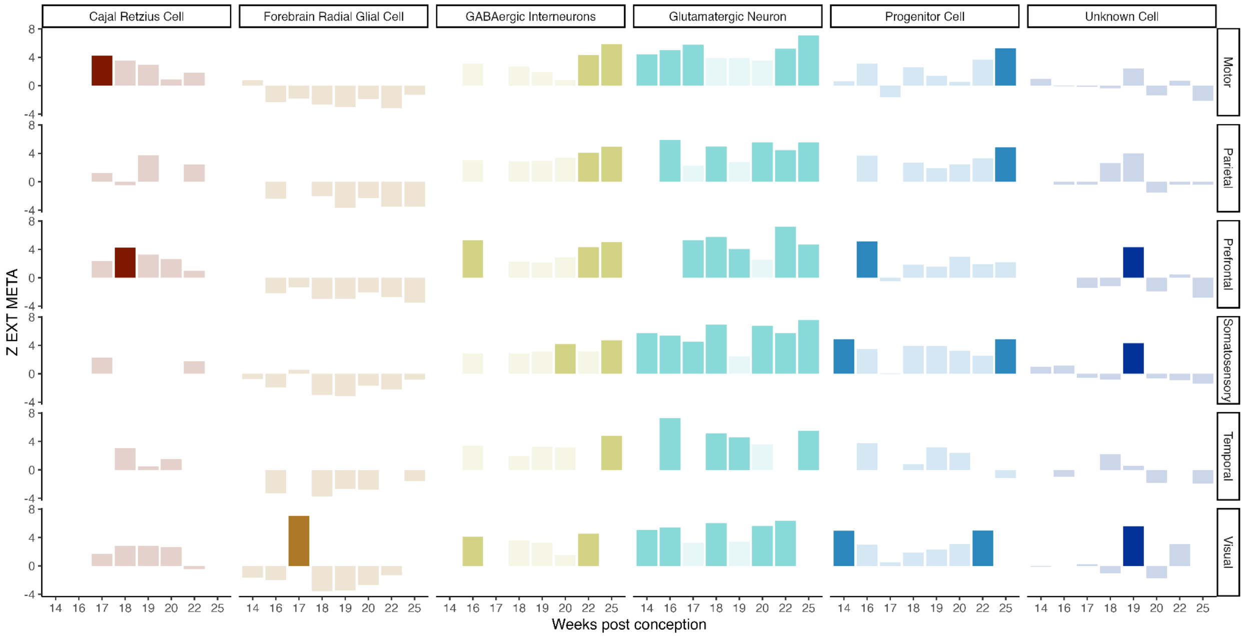


***Figure S9. Enrichment of externalizing liability in specific cell types across brain regions during mid-to-late prenatal brain development.*** The $y$-axis reflects the $Z$ statistic from the enrichment test. Darker shades indicate associations that remain significant after Bonferroni correction. Only cell types with at least one significant association are shown.

*First trimester to adolescence.* Zooming out to consider the human neocortex from the first trimester to adolescence, externalizing liability was significantly enriched in Cajal-Retzius cells, GABAergic neurons, and glutamatergic neurons, with widespread enrichment across the brain and consistent throughout cortical development (**Figure S10**; **Supplementary Table 20**). A more specific enrichment pattern was observed for intermediate progenitor cells committed to the excitatory neuron lineage (IPC EN), restricted to Brodmann area 10 during the second trimester. Oligodendrocyte precursor cells were also enriched across development in a few distinct brain regions.

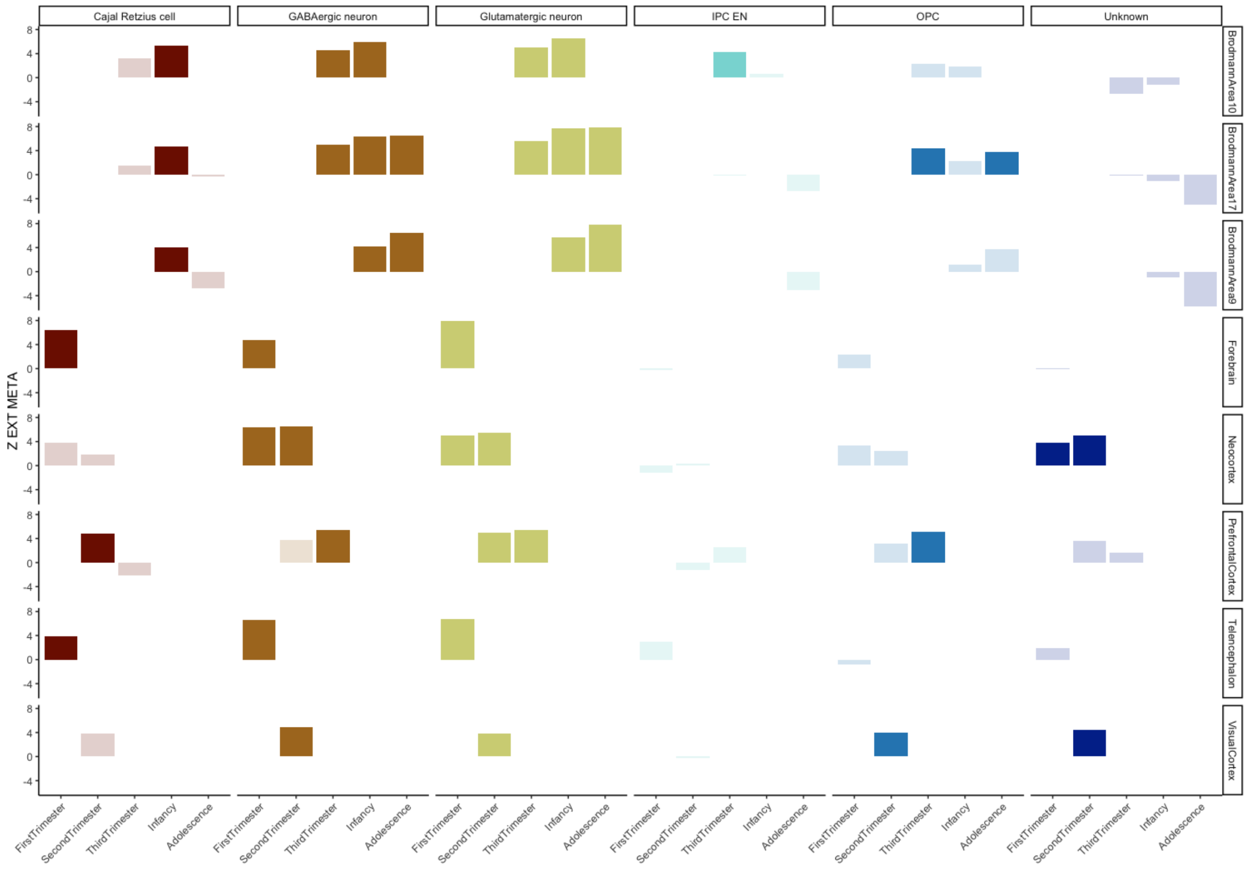


***Figure S10. Enrichment of externalizing liability in specific cell types across brain regions from the first trimester to adolescence.*** The *y*-axis reflects the *Z* statistic from the enrichment test. Darker shades indicate associations that remain significant after Bonferroni correction. Only cell types with at least one significant association are shown. IPC EN = intermediate progenitor excitatory neuron, OPC = oligodendrocyte precursor cells.

*Adulthood (grey matter)*. Finally, when considering grey matter development spanning the entire adult life course, we observed enrichment of externalizing liability in both neurons and oligodendrocyte precursor cells across all investigated brain regions (**Figure S11; Supplementary Table 21**), with stronger effects in neurons.

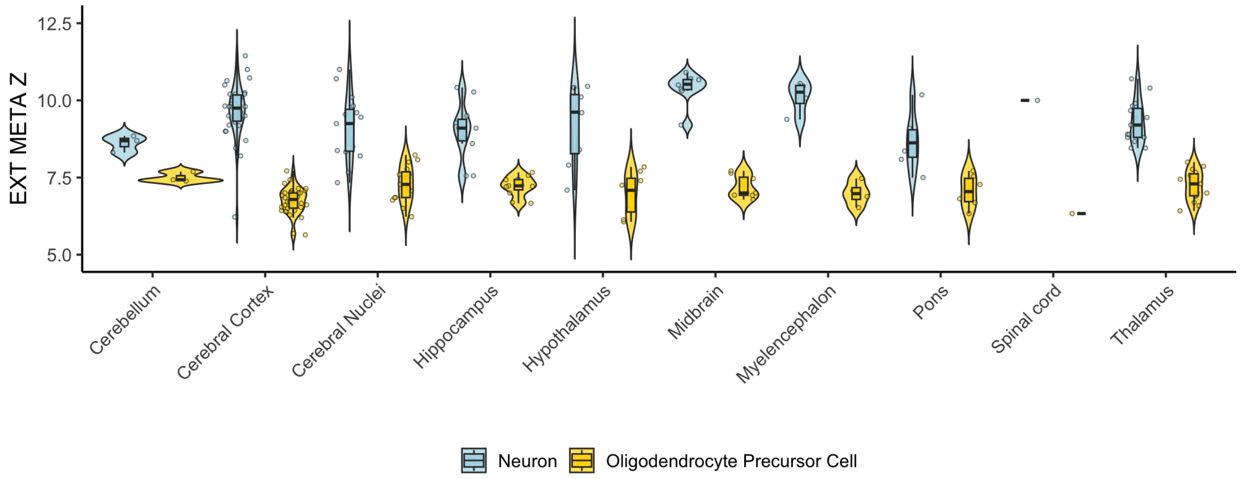


***Figure S11. Enrichment of externalizing liability in specific cell types across grey matter regions.*** The *y*-axis reflects the *Z* statistic from the enrichment test. Each violin plot represents the distribution of enrichment scores for a given cell type across regions, with the central bar indicating the median. All tested cell types showed significant enrichment.

*Adulthood (white matter)*. When considering white matter in middle- and older-adulthood, we observed enrichment of externalizing liability across the primary motor cortex (Brodmann area 4), the arbor vitae of the cerebellum, and the cervical spinal cord, including the fasciculi cuneatus and gracilis (**Figure S12**; **Supplementary Table 22**). Across these white matter regions, consistent enrichment was detected in cerebellar granule cells, GABAergic neurons, glutamatergic neurons, and oligodendrocyte precursor cells. In contrast, enrichment for committed oligodendrocytes and neurons showed age specificity. Oligodendrocyte enrichment was prominent in middle adulthood (ages 30–45) but not in late adulthood (ages 60–75). Neuronal enrichment was not observed in the cervical spinal cord in middle adulthood.

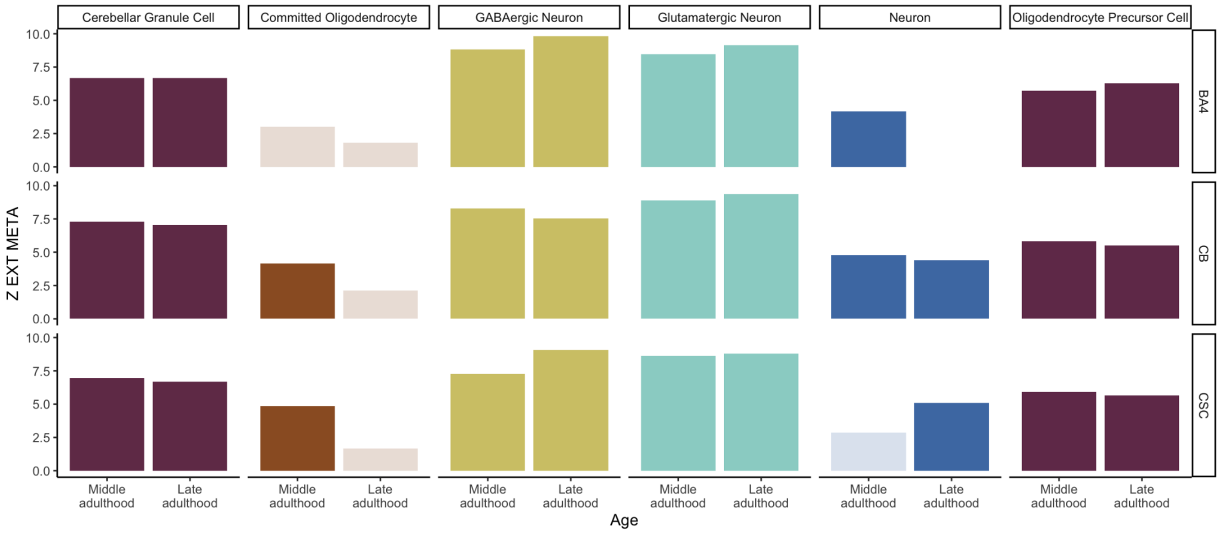


***Figure S12. Enrichment of externalizing liability in specific cell types across white matter brain regions from middle to late adulthood.*** The *y*-axis reflects the *Z* statistic from the enrichment test. Darker shades indicate that associations remain significant after Bonferroni correction. BA4 = Brodmann area 4 (i.e., the primary motor cortex) CB = the arbor vitae of the cerebellum CSC = the cervical spinal cord, including the fasciculi cuneatus and gracilis.

*Convergence across GWAS results*. Patterns of enrichment were highly consistent across the $EXT_{META}$ and $EXT_{EUR}$ results (correlation of coefficients: $r$ = 0.995, s.e. = 0.001). Of the 6,798 associations tested, 883 (13%) were significant in both analyses. An additional 73 associations were uniquely significant for $EXT_{META}$, while 36 were uniquely significant for $EXT_{EUR}$. Although not significant across both $EXT_{META}$ and $EXT_{EUR}$ results, the 73 and 36 associations were highly correlated (correlation of coefficients: $r$ = 0.963, s.e. = 0.032, and $r$ = 0.983, s.e. = 0.031, respectively). Results for $EXT_{META}$ and $EXT_{AFR}$ were also broadly similar (correlation of coefficients: $r$ = 0.766, s.e. = 0.008). However, only 13 associations reached significance after Bonferroni correction in $EXT_{AFR}$. There were no unique associations for the $EXT_{AFR}$ results.

Enrichment results for $EXT_{EUR}$ and EXT1 were very similar as well (correlation of coefficients: $r$ = 0.980, s.e. = 0.002), with 714 (11%) associations significant in both datasets. Notably, $EXT_{EUR}$ identified 205 additional significant enrichment signals, and none were unique to EXT1.

***Enrichment in biological processes, components, and functions***. Both the $EXT_{META}$ and $EXT_{EUR}$ MAGMA gene-level results showed enrichment in C5 GO gene sets related to neurogenesis, neuron differentiation, generation, and development, neuronal projection, chromatin, the synapse, synaptic assembly and organization, the postsynapse, and neuronal structures (axon, dendritic tree, and somatodendritic compartment; **Figure S13**; **Supplementary Table 23**).

The $EXT_{META}$ results were also significantly enriched in gene sets related to the distal axon, spine formation, and neurofibrillary tangle assembly. These associations were of similar magnitude, but below the significance threshold, for the $EXT_{EUR}$ results.

The $EXT_{EUR}$ results were also significantly enriched in gene sets related to RNA metabolic processes, RNA polymerase transcription, axon development, synaptic signaling, and central nervous system development. Although these associations were not significant in the $EXT_{META}$ results, the $EXT_{META}$ and $EXT_{EUR}$ MAGMA gene-set results were generally highly consistent (correlation of coefficients: $r = 0.946$, s.e. = 0.003).

Enrichment results for $EXT_{EUR}$ and EXT1 were strongly correlated (correlation of coefficients: $r = 0.733$, s.e. = 0.007), and 11 of the 19 $EXT_{EUR}$ significant gene sets were also significant for EXT1. Compared to the $EXT_{META}$ results, EXT1 results were not significantly enriched for gene-sets related to neuron structure (e.g., axon, distal axon, neuron spine) and instead showed significant enrichment for gene sets related to regulatory processes (e.g., regulation of synapse assembly, regulation of transcription by RNA polymerase II). No significant enrichment results were observed for the $EXT_{AFR}$ MAGMA results.

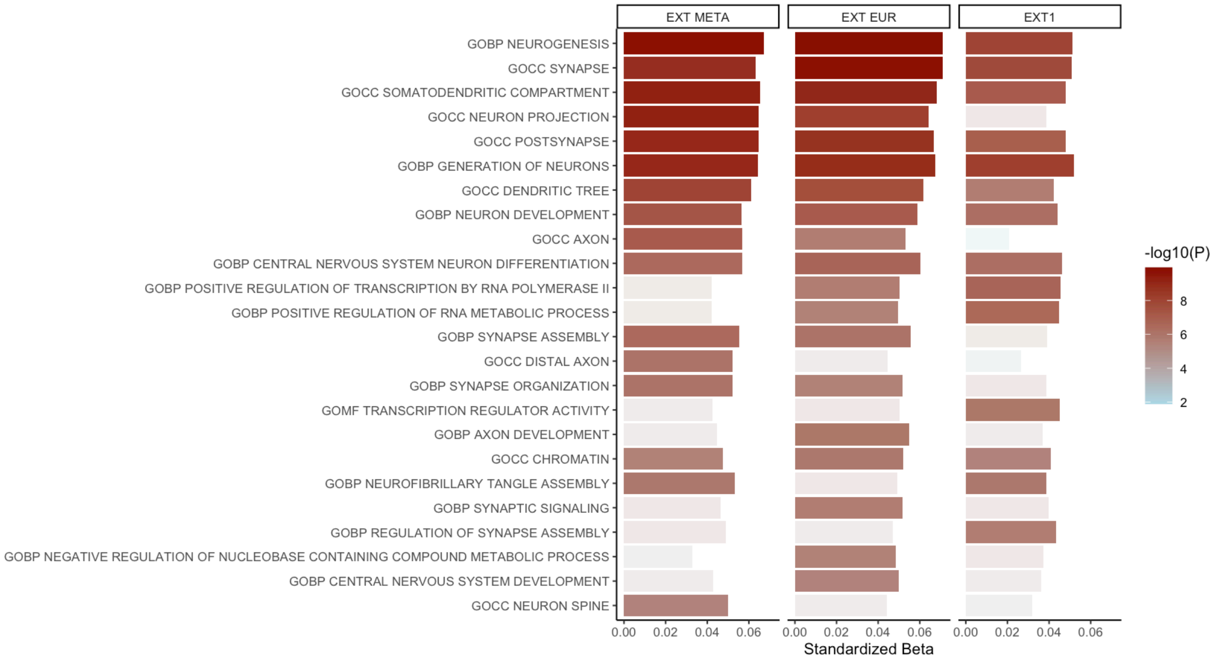


***Figure S13. Patterns of gene set enrichment across externalizing GWASs.*** Significantly enriched gene sets from the C5 Gene Ontology collection were identified by applying MAGMA to multiple externalizing GWASs. Results are shown for the meta-analysis combining EUR- and AFR-like ancestry groups ($EXT_{META}$), the European-only analysis ($EXT_{EUR}$), and the prior externalizing GWAS (EXT1). No significant gene set enrichments were found in the $EXT_{AFR}$ analysis. A Bonferroni correction was applied based on the number of gene sets tested per GWAS. The color scale represents the $-\log_{10}(P)$ of enrichment; lighter shaded bars indicate non-significant associations.

***Enrichment in gene targets of pharmacological compounds.*** DRUGSETS compound-level analyses revealed significant enrichment across multiple drug-gene sets, with consistent patterns observed for both $EXT_{META}$ and $EXT_{EUR}$, with $EXT_{AFR}$ showing greater variability (**Supplementary Table 26**). In total, 73 compounds were significantly enriched in the $EXT_{META}$ results, 71 in the $EXT_{EUR}$ results, and 56 in the $EXT_{AFR}$ results. Of the compounds significantly enriched in the $EXT_{META}$ results, 88% (64/73) were also significant in $EXT_{EUR}$, 34% (25/73) in $EXT_{AFR}$, and 26% (19/73) in both $EXT_{EUR}$ and $EXT_{AFR}$. Six compounds were only significant in $EXT_{EUR}$, although they were close to the significance threshold for the $EXT_{META}$ results ($P = 4.27e\text{-}5$). One compound (ribociclib) was significant in both $EXT_{EUR}$ and $EXT_{AFR}$ but not in $EXT_{META}$, with the META $P$ ($P$ = 6.13e-5) near the Bonferroni threshold ($P$ = 4.27e-5). In contrast, 30 compounds were uniquely implicated in the $EXT_{AFR}$ results. Some of these compounds approached significance in $EXT_{META}$, but others were clearly non-significant ($P$ > 0.05). These included oncology drugs (acalabrutinib, axitinib, bicalutamide, nintedanib, tivozanib), antihypertensives (irbesartan, quinapril), and an anti-inflammatory immunosuppressant (methylprednisolone).

$EXT_{META}$ and $EXT_{EUR}$ signals were largely similar when grouping by mechanism of action, clinical indication, and ATC classifications, while the $EXT_{AFR}$ results again showed more variability, consistent with the compound-level analyses (**Supplementary Table 27**). When compounds were grouped by their mechanism of action, we identified nine significant mechanisms enriched for $EXT_{META}$, all of which were also significantly enriched for $EXT_{EUR}$. These included: dopamine receptor antagonists, GABA receptor antagonists, histone deacetylase (HDAC) inhibitors, RAF inhibitors, dopamine receptor agonists, potassium channel blockers, progesterone receptor agonists, calcium channel blockers, and glutamate receptor antagonists. Two of these mechanisms were also significant in $EXT_{AFR}$ results: dopamine receptor antagonists and agonists.

Pathways were enriched in the $EXT_{META}$ analyses for drugs used to treat schizophrenia ($P$ = 2.71e-12), depression ($P$ = 3.58e-4), psychosis ($P$ = 1.94e-7), seizures ($P$ = 8.25e-5), anesthetics ($P$ = 1.37e-4), melanoma ($P$ = 1.77e-10), and induce muscle relaxation (muscle relaxants, $P$ = 1.29e-4). These drug pathways were also enriched in the $EXT_{EUR}$ results, except for the drug pathways linked to depression. Three of these pathways, drugs related to schizophrenia, psychosis, and muscle relaxants, were also significantly enriched in the $EXT_{AFR}$ results.

At the level of ATC classifications, the $EXT_{META}$ results showed enrichment for antipsychotics (N05A, $P$ = 7.07e-13), anesthetics (N01A, $P$ = 1.17e-8), antiepileptics (N03A, $P$ = 2.38e-7), and dopaminergic agents (N04B, $P$ = 1.47e-5). These pathways were also enriched in the $EXT_{EUR}$ results, and only two of these, N05A and N04B, were enriched in the $EXT_{AFR}$ results.

At the level of mechanisms of action, ancestry-specific results for $EXT_{EUR}$ included serotonin-norepinephrine reuptake inhibitors ($P$ = 5.203e-4) and drugs for chronic obstructive pulmonary disease ($P$ = 3.33e-4). We observed enriched signals for drugs used to treat epilepsy ($P$ = 2.41e-4) and for ATC classes related to endocrine therapy (L02A, $P$ = 7.17e-5) and muscle relaxants (M03A, $P$ = 2.33e-5). These were close to the significance threshold in the $EXT_{META}$ results (median $P$ = 8.01e-3), except for L02A ($P$ = 3.71e-2).

Ancestry-specific results for $EXT_{AFR}$ included estrogen receptor agonists (*P* = 1.33e-4), acetylcholine receptor antagonists (*P* = 6.82e-5), adrenergic receptor agonists (*P* = 1.43e-4), and VEGFR inhibitors (*P* = 1.26e-5). We observed enriched pathways for drugs used to treat menopause (*P* = 2.86e-9), Parkinson's disease (*P* = 1.88e-6), heartburn (*P* = 2.08e-4), and breast cancer (*P* = 7.03e-5). Except for menopause (*P* = 1.30e-3) and Parkinson's disease (*P* = 4.72e-3), these groupings did not approach nominal significance in the $EXT_{META}$ results, which may reflect the noisier polygenic signal in $EXT_{AFR}$.

#### 6.3.2. Bioinformatic characterization of effector genes

We found that 238 of 961 (25%) effector genes were robustly linked to synaptic structure or function, mapping to a SynGO annotation under the high-stringency experimental evidence filter. Among these 238 genes, 200 were linked to synaptic cellular component annotations, and 145 were linked to synaptic biological process annotations, with 107 genes mapping to both. After testing for enrichment—requiring ≥ 3 overlapping genes for inclusion—we found that 22 cellular component terms and 16 biological process terms were enriched at 1% FDR (the SynGO default). Full results are available in **Supplemental Table 25**.

### 7. Polygenic prediction

**Section authors**: Holly E. Poore, Camille M. Williams, Diego Londono-Correa, Yuchen Ning, Natasia S. Courchesne-Krak, Matthew Rosenblatt, Sandra Sanchez-Roige, K. Paige Harden, Richard Karlsson Linnér, & Travis T. Mallard

#### 7.1. Introduction

We evaluated the predictive power of the externalizing polygenic index (PGI) across six independent samples with varied demographic characteristics and contexts. Specifically, we assessed how strongly the PGI was associated with outcomes across four categories: clinical presentations, health risk behaviors, personality, and social and medical correlates. We first describe the six independent samples used. We then describe the methodology employed to generate and test the externalizing PGI across contexts, including the estimation of population and direct effects. Finally, we conclude with a summary of the significant results in each cohort.

#### 7.2. Samples

We selected six independent samples for polygenic prediction: (i) the Collaborative Study of the Genetics of Alcoholism[16] (COGA), (ii) the National Longitudinal Study of Adolescent Health[14,15] (Add Health), (iii) the Adolescent Brain and Cognitive Development Study[72] (ABCD), (iv) the Millennium Cohort Study[73] (MCS), (v) the sibling subset of UK Biobank[13] (UKB), and (vi) the Vanderbilt University Medical Center biobank[17] (BioVU). Collectively, these samples cover a wide range of ages (early childhood to late adulthood), recruitment strategies (e.g., population representative versus clinically referred samples), and ancestries (including both EUR-like and AFR-like individuals). Except for BioVU, each sample also includes at least a subset of related individuals for whom familial structure is known, allowing for the estimation of population and direct effects.

For each sample, we selected traits for analysis from four phenotypic domains: clinical presentations (e.g., measures of psychiatric symptoms or diagnoses); health risk behaviors (e.g., measures of substance use and risky sex); social and medical consequences (e.g., measures of educational, social, and medical problems); and personality traits (e.g., measures of temperament). These variables, and the preprocessing steps involved in their preparation, are described in **Supplementary Tables 28–33**.

#### 7.2.1. Collaborative Study on the Genetics of Alcoholism (COGA)

COGA[16] is a U.S.-based, multi-site, family study of the etiology of alcohol use disorders and related conditions. Individuals in treatment for alcohol use disorder and their families, as well as community-based comparison families, were recruited beginning in 1991 and followed continuously until present day. Data were collected over three waves. For this study, we used data from all waves for EUR-like ($N_{max}$ = 7,483) and AFR-like ($N_{max}$ = 3,358) participants for whom genotype and phenotypic data were available. Measures from COGA spanned three domains:

- ***Clinical presentations:*** Interview- and self-report measures were used to assess DSM-IV-defined alcohol, cannabis, and other substance use disorder (combined abuse and dependence criteria), as well as attention-deficit/hyperactivity disorder, conduct disorder/antisocial personality disorder (depending on the age of the participant), and suicidality (ideation and attempt). The Fagerström Test for Nicotine Dependence[74] was used to measure symptoms of nicotine dependence.
- ***Health risk behaviors:*** Self-report measures were used to assess substance initiation and involvement (alcohol, cannabis, opioids), patterns of heavy and/or episodic use, and early/risky sexual behavior. Achenbach Self-Report scales were used to measure rule-breaking and aggressive tendencies[75].
- ***Social and medical consequences:*** Self-report measures were used to assess criminal justice involvement (ever arrested, number of times arrested, ever convicted of a felony, ever incarcerated), sexual health and reproductive outcomes (number of live births, number of pregnancies, lifetime diagnosis of sexually transmitted infections), and socioeconomic outcomes (educational attainment, household income, full-time employment).

#### 7.2.2. National Longitudinal Study of Adolescent to Adult Health (Add Health)

Add Health[14,15] is a nationally representative, longitudinal study of young people in the U.S. Participants were 11- to 18-years-old at Wave 1 (1994-1995) and were 35- to 42-years-old at Wave 5 (2016-2018), with data collection ongoing. For this study, we used data from Waves 1-4 for participants of EUR- ($N_{max}$ = 5,645) and AFR-like ($N_{max}$ = 2,025) ancestry for whom genotype and phenotypic data were available. Measures from Add Health spanned three broad domains:

- ***Clinical presentations:*** Self-report measures were used to assess lifetime and current experiences of clinically significant psychiatric problems and substance use, including symptom-level measures of DSM-IV[76]-defined attention-deficit/hyperactivity disorder and alcohol, cannabis, and other substance use disorder (combined abuse and dependence criteria). A self-report measure was used to assess whether participants had ever received a formal diagnosis of attention-deficit/hyperactivity disorder. The Fagerstrom Test for Nicotine Dependence[74] was used to assess symptoms of nicotine dependence.
- ***Health risk behaviors:*** Self-report measures were used to lifetime and recent substance use (alcohol, tobacco, cannabis, and other illicit substances including sedatives, tranquilizers, stimulants, painkillers, steroids, cocaine, crystal meth), risky sexual behavior (age at first sexual intercourse, condom use), and rule-breaking and aggressive behaviors (e.g., vandalism, carrying/using a weapon, skipping school).
- ***Social/medical consequences:*** Self-report measures were used to assess indicators of consequences across sexual/reproductive (number of live births, number of pregnancies, lifetime diagnosis of sexually transmitted infections), educational (problems in school, attitudes toward school, ever expelled/held back/suspended, college completion, educational attainment, GPA), socioeconomic (personal and household income, net worth, debt-to-asset ratio, months insured during past year, ever fired from job, current unemployment), and criminal justice (ever arrested, ever convicted, ever incarcerated) domains.

##### 7.2.3. Adolescent Brain and Cognitive Development (ABCD) Study

The ABCD Study[72] is a multi-site study of U.S. adolescents, ages 9-10 at baseline, with participants assessed every 6 months and neuroimaging occurring every 2 years through age 18. For this study, we used baseline data for participants of EUR- ($N_{max}$ = 5,686) and AFR-like ($N_{max}$ = 1,923) ancestry for whom genotype and phenotypic data were available. Measures from ABCD spanned three broad domains:

- ***Clinical presentations:*** Parent-report measures from the Child Behavior Checklist[77] (CBCL) were used to measure the child's behavioral and emotional problems. This included DSM-oriented symptom scales for attention-deficit/hyperactivity disorder, conduct disorder, and total externalizing problems, as well as additional item/subscale scores indexing the child's aggressive behavior, attention problems, impulsivity, rule-breaking behavior, and social problems.

- ***Health risk behaviors:*** Youth self-report measures were used to measure participant experimentation with substance use and endorsement of deviant peer influences. Substance use measures included a lifetime measure of any alcohol use (even sips) and a lifetime measure of being offered tobacco.
- ***Personality:*** Youth self-report measures from the Behavioral Inhibition System/Behavioral Activation System (BIS/BAS)[78] Scales were used to assess motivational tendencies related to sensitivity to punishment and reward (i.e., the Drive, Fun Seeking, and Reward Responsiveness subscales). Similar self-report measures from the Urgency, Premeditation (lack of), Perseverance (lack of), Sensation Seeking, and Positive Urgency (UPPS-P) Impulsive Behavior Scale for children[79] were used to measure five distinct dimensions of impulsivity.

#### 7.2.4. Millennium Cohort Study (MCS)

The MCS[73] is a nationally representative birth cohort following children born in the U.K. in ~2000–2002 with repeated assessments through age 17. The original cohort comprised approximately 18,827 children, with follow-up assessments conducted at 9 months and 3, 5, 7, 11, 14, and 17 years. Genotype data were available for approximately 21,192 individuals, including both cohort members and their parents. For this study, we restricted the sample to genotyped individuals of EUR-like ancestry with phenotypic data suitable for population-level ($N_{max}$ = 8,043) and parent-child trio-based analyses (trio $N_{max}$ = 2,799). Measures from MCS were taken at age 17 unless otherwise indicated and spanned two broad domains, as summarized below.

- ***Clinical presentations:*** Parent-, teacher-, and youth-report measures from the Strengths and Difficulties Questionnaire[80] (SDQ) were used to measure hyperactivity/inattention and conduct problems. We also created a composite externalizing symptom score from these subscales. Data were standardized within each wave and informant type and then averaged to derive composite scores per wave.
- ***Health risk behaviors:*** Youth self-report surveys were used to assess lifetime initiation of alcohol use, cigarette/e-cigarette use, cannabis use, as well as lifetime sexual activity.

#### 7.2.5. The UK Biobank (UKB) Siblings

The UKB[13] is a population-based cohort with more than 500,000 participants recruited between 2006 and 2010. For validation, we created a UKB sibling hold-out sample for PGI analyses, as previously reported[1]. We constructed a subsample of approximately 40,000 individuals from ~19,000 families and withheld them from the current discovery GWAS. We further ensured that participants in the UKB sibling subsample had no other close relatives remaining in the discovery sample (KING coefficient < 0.0442 for all pairwise comparisons). This design supported both large-scale population-level and within-family tests ($N_{\max}$ = 39,640). Measures from the UKB sibling hold-out sample spanned four broad domains:

- ***Clinical presentations:*** Problematic alcohol use measured from ICD-9/10 diagnoses reported in EHR data and medical conditions disclosed during an interview
- ***Health risk behaviors:*** Self-report measures indexed lifetime smoking initiation, cigarettes per day, drinks per week, lifetime cannabis initiation, number of sexual partners, age at first sexual intercourse, and automobile speeding propensity.
- ***Social/medical consequences:*** Self-report and registry-linked sociodemographic indicators were used to measure educational attainment, fluid intelligence, household income, overall health, housing tenure, and neighborhood deprivation (Townsend Deprivation Index). Reproductive outcomes were also measured via self-report questionnaires, including the number of children ever born (both sexes), children fathered (males), and, for females, live births, age at first birth, and teenage conception.
- ***Personality:*** Self-report measures indexed general risk tolerance (1-item), neuroticism, and subjective well-being. For neuroticism, we analyzed both the total score and individual items from the Eysenck Personality Questionnaire-Revised Short Form's Neuroticism scale[81], such as those measuring tendencies to be highly strung, feel worried, and ruminate after embarrassment.

#### 7.2.6. Vanderbilt University Medical Center Biobank (BioVU)

BioVU[17] is a U.S.-based biobank of EHR data from the Vanderbilt University Medical Center on more than two million patients spanning 1990 to 2025. For this study, we used data from participants of EUR-like ($N_{\max}$ = 66,184) and AFR-like ($N_{\max}$ = 12,250) ancestries for whom genotype and phenotypic data were available. Measures from BioVU spanned two broad domains: clinical presentations and social/medical consequences.

- ***Clinical presentations and social/medical consequences.*** EHR data were used to construct case/control "phecodes" based on ICD-9/10 codes spanning: psychiatric and substance-related disorders, as well as broader medical conditions and sequelae. Phecodes were organized in the following established categories: infectious diseases, neoplasms, endocrine/metabolic diseases, hematopoietic diseases, mental disorders, neurological disorders, sense organ disorders, circulatory system disorders, respiratory diseases, digestive diseases, genitourinary diseases, pregnancy complications, dermatologic disorders, musculoskeletal disorders, congenital anomalies, disease symptoms, and injuries/poisonings. Further information regarding phecode definitions, mapping, and analytic procedures, including multiple-testing thresholds and sensitivity analyses, is detailed in the BioVU phenome-wide association study (PheWAS) **Supplementary Section 7.3.3**.

### 7.3. Methodology

We define predictive power as the incremental variance explained ($\Delta R^2$) achieved by adding the PGI to a baseline regression model with covariates. All association tests were conducted within-ancestry; no cross-ancestry pooling of individual-level data was performed. Across five of the six cohorts, we implemented a harmonized protocol that quantified both population-level and direct genetic effects. BioVU is the only cohort for which we did not follow this protocol, as it was used only for estimating population-level effects of the externalizing PGI across the medical phenome.

#### 7.3.1. Polygenic index construction

To maximize information from both EUR- ($\widehat{N}$ = 3,943,075) and AFR-like ($\widehat{N}$ = 303,641) samples, we estimated posterior SNP effect sizes using PRS-CSx[82] v1.1.0, a Bayesian approach that integrates GWAS results across multiple ancestries while accounting for population-specific LD. All PRS-CSx analyses used population-specific LD reference data from the EUR- and AFR-like samples of the 1000 Genomes Project Phase 3 v5 dataset[21], restricted to HapMap3 SNPs. As we sought to generate a single set of weights to apply across multiple cohorts, we did not prespecify a global shrinkage parameter; instead, we opted to learn this parameter automatically from the discovery summary statistics. We also employed the `meta` approach within PRS-CSx, which integrated population-specific posterior SNP effects via an inverse-variance-weighted meta-analysis within the Gibbs sampler. This yielded a final set of 1,133,211 high-quality SNP-level weights for the $EXT_{META}$ GWAS, 1,031,200 for the $EXT_{EUR}$ GWAS, 1,031,126 for the $EXT_{EUR-23andMe}$ GWAS, and 552,102 for the $EXT_{AFR}$ GWAS, suitable for constructing externalizing PGIs across cohorts.

Using PLINK[83] v1.9/2, we calculated four versions of the externalizing PGI: the cross-ancestry index ($PGI_{META}$, our primary PGI) and three ancestry-specific indices ($PGI_{EUR}$, $PGI_{EUR-23andMe}$, $PGI_{AFR}$). These individual-level PGIs were subsequently standardized to a zero mean and unit variance within each ancestry in each sample. This process ensured that cross-cohort differences in predictive performance reflect substantive between-cohort differences (*e.g.*, composition, ascertainment) rather than differences in PGI construction.

#### 7.3.2. Effect estimation framework

Within each cohort and ancestry group, we implemented a three-stage protocol designed to quantify population-level predictive power, validate signals in family subsamples, and estimate direct genetic effects. All analyses were conducted in R[24] and used a common set of covariates tailored to the standardized implementation: sex, age (omitted where invariant, e.g., MCS), and at least the first 10 genetic principal components for all cohorts except the UK Biobank, which included the first 20. Cohort-specific covariates (e.g., site or batch) were added where appropriate. Standard errors were clustered at the family level for all cohorts that included siblings.

In the *population-level step*, each outcome $Y$ was regressed on the externalizing PGI $S$ and covariates $X$, as

$$Y = X\theta + S\beta + \epsilon$$

and predictive power was summarized as $\Delta R^2 = R^2(\text{PGI + covariates model}) - R^2(\text{covariates-only model})$. To quantify uncertainty in $\Delta R^2$, we used a nonparametric bootstrap with 200 repetitions to form 95% confidence intervals. Resampling respected dependence structure: in samples with related individuals, bootstrap draws were taken at the family level so that sibling sets (or trios) remained intact within resamples, and then all members of a selected family were included together. This approach preserves the within-family variation that later identifies direct effects while providing stable uncertainty quantification for $\Delta R^2$ contrasts.

In the *family-validation step*, outcomes that were significant after Bonferroni correction (within each cohort and ancestry) were re-estimated in the family-restricted subsample using specifications that did not yet absorb family effects (i.e., siblings without family fixed effects, trios without parental PGIs). This gatekeeping step ensured that signals were sufficiently stable under reduced sample size and the different endorsement patterns before proceeding to within-family identification.

In the *direct-effect step*, we estimated within-family models that disambiguated the effect of the PGI from population stratification, assortative mating, dynastic effects, and shared environment. In sibling designs (ABCD, Add Health, COGA, UKB), identification is derived from within-family contrasts

$$Y = X\theta + S\beta + D\alpha + \epsilon$$

with a dummy intercept α for each family. In trio designs (MCS), we jointly included the child's PGI and both parental PGIs so that $\beta$ captures the direct effect of transmitted alleles conditional on parental genotype.

To formally test attenuation when adjusting for family effects, we contrasted the PGI coefficient from the family-restricted model without family effects (or without parental PGIs in trios) against the coefficient from the within-family model. The difference,

$$\Delta\hat{\beta} = \hat{\beta}_{withFFE} - \hat{\beta}_{withoutFFE}$$

was paired with a closed-form standard error derived from the joint covariance of the stacked estimators (matrix solution following the approach described in EXT1[1]), using the residual variance estimate from the relevant baseline. We then calculated $Z$ as $\Delta\beta$ / s.e.($\Delta\beta$) and applied a Bonferroni-adjusted $|Z|$ threshold based on the number of within-family associations under consideration. Only outcomes that (i) were significant in the population models and (ii) persisted in the family-restricted validation step were carried forward to within-family models, thereby controlling multiple testing burden at the direct-effect stage and focusing inference on robust signals. Unless otherwise specified, we evaluated statistical significance using a Bonferroni correction within each cohort, ancestry, and analysis.

Across stages, effects were estimated with ordinary least squares for both continuous and binary outcomes (i.e., a linear probability model). This choice is deliberate for three reasons. First, it yields a single, comparable estimand (the mean change in the outcome per 1-SD increase in the PGI) across continuous and binary traits and permits a coherent definition of $\Delta R^2$—a core quantity in this section—without resorting to pseudo-$R^2$ measures with different scales. Second, it facilitates within-family identification: a fixed-effects logit model in this context is susceptible to severe bias (i.e., the "incidental parameter problem") and drops families with no within-family variation in the outcome, whereas the linear model retains all informative families and cleanly nests the with "versus without family effects" contrast required for a formal test of attenuation. Third, in large samples with moderate event rates, the linear probability model provides a close approximation to marginal effects from nonlinear probability models and avoids the non-collapsibility of the odds ratio that can complicate comparisons across nested specifications.

#### 7.3.3. Cohort-specific analyses

In addition to the standardized protocol described above, we conducted several analyses that exploited cohort-specific design and measurement features. Below, we describe these cohort-specific procedures, each implemented to probe the utility and robustness of the externalizing PGI across settings.

***Modeling latent externalizing factors in Add Health and COGA.*** In Add Health and COGA, we evaluated the ability of the externalizing PGI to predict a latent externalizing factor with indicators matching the discovery model as closely as possible. These models were estimated using M*plus*[84] version 8, which allowed us to fit confirmatory factor models with indicators that have different scales of measurement (*i.e.*, dichotomous and continuous variables). Model fit was assessed on the basis of the CFI, TLI, RMSEA, and SRMR, with values of CFI and TLI ≥ .95, SRMR ≤ 0.08, and RMSEA ≤0.05 indicating good fit[55].

Model fit and factor loadings from ancestry-stratified models are presented in **Supplementary Table 28**. Note that models were tested separately for participants of EUR- and AFR-like ancestry groups due to evidence of metric invariance (i.e., unequal factor loadings) across these groups. For the purposes of within-family analyses, which require the addition of family fixed effects in a manner that is not compatible with M*plus*, we estimated externalizing factor scores via the *FSCORES* function. These factor scores represent factor loading-weighted sums of the seven indicator phenotypes for each individual, yielding an observed measure suitable for the previously described effect estimation procedures.

***Evaluating the impact of 23andMe data in Add Health and COGA.*** To assess whether the inclusion of 23andMe data influenced downstream results, we re-estimated the multivariate discovery GWAS excluding that data. As only one indicator (SMOK) contained 23andMe data in the EUR-like ancestry group, removing this data did not meaningfully affect the factor model fit or factor loadings, and the identified polygenic architecture remained robust and stable across model configurations (**Supplementary Section 3**). We subsequently calculated the externalizing PGI using the down-sampled GWAS summary statistics (i.e., $PGI_{EUR-23andMe}$) and repeated the PGI analyses in Add Health and COGA. We compared estimates between the $PGI_{EUR}$ and $PGI_{EUR-23andMe}$ and formally evaluated their concordance.

***Phenome-wide association analyses in BioVU.*** In BioVU, we conducted a PheWAS to characterize associations between the externalizing PGIs and medical outcomes, as measured by EHR data (i.e., phecodes; case/control phenotypes derived from ICD-9/10 data). Phecode mapping followed Phecode Map 1.2[85–87] (Phecode Map 1.2 https://phewascatalog.org), and models were implemented in the PheWAS R package[88] (v0.12). Phecodes were only selected for analysis if they had ≥ 100 cases (with cases also requiring ≥ 2 qualifying ICD codes), resulting in a total of 1,440 phecodes available in EUR-like participants and 614 phecodes available in AFR-like participants. For each phecode, we fit a logistic regression of case status on the PGI, adjusting for sex, the median age of EHR encounters, and 10 ancestry PCs.

*Age-stratified analyses.* To examine developmental patterning of associations, we repeated the PheWAS with the $EXT_{META}$ GWAS in the EUR-like sample within six age bins defined by age at ICD code occurrence in the electronic health records: 0–11 (childhood; *n* = 7,042–8,443), 12–18 (adolescence; *n* = 5,886–6,893), 19–25 (young adulthood; *n* = 6,241–7,211), 26–40 (adulthood; *n* = 13,261–15,437), 41–60 (midlife; *n* = 26,269–31,346), and 61–100 (older adulthood; *n* = 22,405–26,819). A single individual was included in multiple age groups if relevant ICD codes were documented at different time points. We did not conduct age-stratified PheWAS analyses within the AFR-like sample due to limited sample sizes. To formally test whether associations varied across age groups in EUR-like participants, we first focused our attention on phecodes that were FDR-significant in at least two age bins. Among the 486 phenotypes meeting this criterion, we used the metafor[89] v4.8-0 R package to calculate Cochran's Q across age bins, which indexes heterogeneity in a fixed-effects inverse-variance weighting meta-analysis framework, applying the same Benjamini-Hochberg FDR adjustment as before.

*Sensitivity analyses*. We conducted a series of sensitivity analyses using the $PGI_{META}$ in the EUR-like BioVU subsample, adding covariates for the Area Deprivation Index[90] (i.e., socioeconomic disadvantage in a particular region of interest), a global comorbidity score[91] (i.e., an inverse-normal transformed count of the top 600 phecodes), and substance use disorder phecodes (alcohol 316; tobacco 317; other substances 318), and report results that survived an FDR-significance threshold.

#### 7.4. Results

We estimated cross-ancestry and ancestry-specific associations between the externalizing PGI and outcomes across four domains measured in six independent cohorts. Across cohorts, $PGI_{META}$ explained more variance than ancestry-specific scores, with gains over $PGI_{AFR}$ generally larger than gains over $PGI_{EUR}$. As described in the standardized protocol, reported $\Delta R^2$ values reflect incremental variance explained over covariates, and statistical significance was evaluated following Bonferroni correction within cohort and ancestry unless otherwise noted.

##### 7.4.1. COGA

***EUR-like ancestry.*** $PGI_{META}$ and $PGI_{EUR}$ were associated with 26 of 30 outcomes (non-significant: number of live births, full-time employment, alcohol initiation, lifetime sexually transmitted infection; **Supplementary Table 29**). Effect estimates were essentially identical for the two PGIs (correlation of coefficients: $r$ = 1), and differences between the PGIs in variance explained were trivial (median $\Delta R^2$ difference = 0.05%, range −0.1% to 0.1%). $PGI_{META}$ predicted the latent externalizing factor ($\beta$ = 0.352, $\Delta R^2$ = 11.9%) and the factor score ($\beta$ = 0.248, $\Delta R^2$ = 8.2%), a 29-33% improvement over the EXT1 PGI (**Supplementary Table 34**).

With respect to individual phenotypes, the largest associations were observed for FTND symptoms ($\Delta R^2$ = 7.7%), lifetime smoking initiation ($\Delta R^2$ = 6.7%), educational attainment ($\Delta R^2$ = 5.9%), age at first sex ($\Delta R^2$ = 5.5%), antisocial behavior ($\Delta R^2$ = 4.8%), AUD symptoms ($\Delta R^2$ = 4.3%), lifetime other substance initiation ($\Delta R^2$ = 4.1%), and rule-breaking behavior ($\Delta R^2$ = 3.8%). Additional clinical presentations included ADHD symptoms ($\Delta R^2$ = 2.5%), other SUD symptoms ($\Delta R^2$ = 2.2%), OUD symptoms ($\Delta R^2$ = 2.0%), suicide attempt ($\Delta R^2$ = 2.0%), CUD symptoms ($\Delta R^2$

= 1.8%), and suicidal ideation ($\Delta R^2$ = 1.7%). Notable health-risk behavior associations included opioid initiation ($\Delta R^2$ = 2.9%), cannabis initiation ($\Delta R^2$ = 2.6%), violence/aggression ($\Delta R^2$ = 2.6%), and number of sexual partners ($\Delta R^2$ = 1.4%). Social and medical consequences included ever being arrested ($\Delta R^2$ = 2.5%), household income ($\Delta R^2$ = 2.4%), ever being incarcerated ($\Delta R^2$ = 1.5%), ever being convicted ($\Delta R^2$ = 1.0%), and number of arrests ($\Delta R^2$ = 0.9%).

In sibling analyses, a subset of associations remained significant after including family fixed effects: the externalizing factor score, ASPD/CD symptoms, AUD symptoms, educational attainment, ever smoker, and FTND symptoms. Among these, only educational attainment and smoking initiation showed significant attenuation compared to family-unadjusted estimates. The remaining direct-effect estimates did not significantly differ from the family-unadjusted estimates.

***AFR-like ancestry.*** $PGI_{META}$ was associated with 10 outcomes and $PGI_{AFR}$ with 7 outcomes (correlation of coefficients: $r$ = 0.953, s.e. = 0.017; **Supplementary Table 29**). Differences in $\Delta R^2$ were small (median 0.2%, range −0.2% to 1.8%), with $PGI_{META}$ providing the largest improvement for AUD symptoms (+1.0%) and ASPD/CUD symptoms (+0.8%). $PGI_{META}$ predicted the externalizing latent factor ($\beta$ = 0.192, $\Delta R^2$ = 2.6%) and factor score ($\beta$ = 0.137, $\Delta R^2$ = 1.5%). Additional associations included AUD symptoms ($\Delta R^2$ = 1.3%), ASPD/CD symptoms ($\Delta R^2$ = 1.2%), FTND symptoms ($\Delta R^2$ = 0.8%), smoking initiation ($\Delta R^2$ = 0.6%), other substance initiation ($\Delta R^2$ = 0.5%), age at first sex ($\Delta R^2$ = 0.4%), cannabis initiation ($\Delta R^2$ = 0.4%), opioid initiation ($\Delta R^2$ = 0.3%), and number of sexual partners ($\Delta R^2$ = 0.2%). $PGI_{AFR}$ was associated with ever being convicted ($\Delta R^2$ = 0.4%). None of these associations were significant at the family-validation step.

#### 7.4.2. Add Health

***EUR-like ancestry.*** $PGI_{META}$ and $PGI_{EUR}$ were associated with 38 of 40 outcomes (non-significant: current employment, condom use in the last 12 months; **Supplementary Table 30**). Effect estimates were virtually identical for the two PGIs (correlation of coefficients: $r$ = 1), and differences between the PGIs in variance explained were trivial (median $\Delta R^2$ difference = 0.04%, range −0.04% to 0.1%). $PGI_{META}$ predicted the latent externalizing factor ($\beta$ = 0.359, $\Delta R^2$ = 12.6%) and the factor score ($\beta$ = 0.241, $\Delta R^2$ = 8.4%), a 24-26% improvement over the EXT1 PGI (**Supplementary Table 34**).

Among individual phenotypes, the largest associations were observed for lifetime smoking initiation ($\Delta R^2$ = 7.1%), education and school variables ($\Delta R^2$ = 6.0% for college completion, $\Delta R^2$ = 5.4% for educational attainment, $\Delta R^2$ = 4.7% for GPA, and $\Delta R^2$ = 3.9% for ever being suspended from school), age at first sexual intercourse ($\Delta R^2$ = 5.6%), and lifetime cannabis initiation ($\Delta R^2$ = 3.8%). Within variables related to clinical disorders/symptoms, $PGI_{META}$ explained the most variance in symptoms of ADHD ($\Delta R^2$ = 2.4%), FTND symptoms ($\Delta R^2$ = 1.7%), and other SUD symptoms ($\Delta R^2$ = 1.5%) and somewhat less variance in AUD and CUD symptoms ($\Delta R^2$ = 0.6% and 0.4%, respectively). Of the health risk variables, $PGI_{META}$ explained the most variance in smoking and cannabis initiation and age at first sexual intercourse (discussed above) followed by aggression ($\Delta R^2$ = 2.7%), other substance use ($\Delta R^2$ = 2.2%), number of sexual partners ($\Delta R^2$ = 1.6%), rule-breaking behavior ($\Delta R^2$ = 1.2%), and alcohol initiation ($\Delta R^2$ = 0.4%). Of the social/medical consequences variables, $PGI_{META}$ explained the most variance in the school related variables discussed above, but was significantly associated with other school variables, including ever held back a grade ($\Delta R^2$ = 1.6%), school problems ($\Delta R^2$ = 1.5%), positive attitude toward school ($\Delta R^2$ = 1.4%), ever skipped school ($\Delta R^2$ = 1.3%), and ever expelled ($\Delta R^2$ = 1.0%). It was also significantly associated with other socioeconomic variables, including individual income ($\Delta R^2$ = 2.3%), household income ($\Delta R^2$ = 1.7%), net worth ($\Delta R^2$ = 0.9%), and debt-to-asset ratio ($\Delta R^2$ = 0.6%). Finally, it was significantly associated with variables related to contact with the criminal justice system, including ever arrested ($\Delta R^2$ = 2.3%), ever incarcerated ($\Delta R^2$ = 2.0%), number of times arrested ($\Delta R^2$ = 2.0%), and ever convicted ($\Delta R^2$ = 1.1%).

We examined whether these associations were robust to genetic confounding by testing whether significant population-level associations remained significant after adjusting for within-family effects. Only a subset of these associations remained significant in the sibling sample, and of these, only one phenotype retained significance after controlling for family fixed effects (**Supplementary Table 30**). Specifically, both $PGI_{META}$ and $PGI_{EUR}$ still predicted the externalizing factor score after adjusting for family fixed effects, and the betas of these associations were not significantly attenuated (attenuation ratios = 0.771 and 0.752, respectively).

***AFR-like ancestry.*** In the AFR-like ancestry subsample, $PGI_{META}$ was significantly associated with 20 phenotypes, and $PGI_{AFR}$ was associated with 11 phenotypes out of 39 outcomes (Other SUD symptoms, which had fewer than 150 observations, were excluded; **Supplementary Table 30**). Although the beta coefficients of the $PGI_{META}$ and $PGI_{AFR}$ associations were highly correlated (correlation of coefficients: $r$ = 0.958, s.e. = 0.014), we found more pronounced differences in $\Delta R^2$ between $PGI_{META}$ and $PGI_{AFR}$ compared to $PGI_{META}$ and $PGI_{EUR}$. Differences in $\Delta R^2$ ranged from −0.1% to 2.1% (median $\Delta R^2$ difference = 0.3%), with the most substantial improvements observed for the externalizing factor (difference in $\Delta R^2$ = 2.1%), lifetime smoking initiation (difference in $\Delta R^2$ = 1.4%), the externalizing factor (difference in $\Delta R^2$ = 1.3%), ever arrested (difference in $\Delta R^2$ = 1.0%), and having completed college (difference in $\Delta R^2$ = 1.0%).

Both $PGI_{META}$ and $PGI_{AFR}$ were significantly associated with the latent externalizing factor ($\beta_{META}$ = 0.207, $\Delta R^2$ = 3.4%; $\beta_{AFR}$ = 0.123, $\Delta R^2$ = 2.1%) and the externalizing factor score ($\beta_{META}$ = 0.127, $\Delta R^2$ = 2.1%, $\beta_{AFR}$ = 0.075, $\Delta R^2$ = 1.3%). Both PGIs were also significantly associated with ever being arrested ($\Delta R^2_{META}$ = 2.2%, $\Delta R^2_{AFR}$ = 1.2%), ever being incarcerated ($\Delta R^2_{META}$ = 2.1%, $\Delta R^2_{AFR}$ = 1.8%), smoking initiation ($\Delta R^2_{META}$ = 2.0%, $\Delta R^2_{AFR}$ = 0.6%), personal income ($\Delta R^2_{META}$ = 1.9%, $\Delta R^2_{AFR}$ = 0.1%), number of times arrested ($\Delta R^2_{META}$ = 1.8%, $\Delta R^2_{AFR}$ = 0.9%), college completion ($\Delta R^2_{META}$ = 1.6%, $\Delta R^2_{AFR}$ = 0.7%), educational attainment ($\Delta R^2_{META}$ = 1.6%, $\Delta R^2_{AFR}$ = 0.7%), ever convicted ($\Delta R^2_{META}$ = 1.2%, $\Delta R^2_{AFR}$ = 0.6%), and rule-breaking behavior ($\Delta R^2_{META}$ = 0.6%, $\Delta R^2_{AFR}$ = 0.5%). $PGI_{META}$ was also associated with number of pregnancies ($\Delta R^2$ = 0.6%), cannabis initiation ($\Delta R^2$ = 0.6%), ever being held back in school ($\Delta R^2$ = 0.6%), ever being suspended ($\Delta R^2$ = 0.6%), being fired from work ($\Delta R^2$ = 0.8%), GPA ($\Delta R^2$ = 0.8%), having health insurance ($\Delta R^2$ = 0.8%), household income ($\Delta R^2$ = 1.0%), and ever using a weapon ($\Delta R^2_{META}$ = 1.2%). Only three of these associations remained significant in the sibling sample, and none remained significant after adjusting for family fixed effects (**Supplementary Table 30**).

#### 7.4.3. Evaluating the impact of 23andMe data on predictive accuracy

To evaluate the degree to which including 23andMe data in our discovery analyses affected polygenic performance, we compared four versions of the EXT PGI: $PGI_{EUR}$ (i.e., based on the present $EXT_{EUR}$ results), $PGI_{EUR-23andMe}$, $PGI_{EXT1}$ (i.e., based on our prior EXT GWAS[1]), and $PGI_{EXT1-23andMe}$[51]. We found that $Z$ statistics and incremental $R^2$ values were correlated at near unity between the $PGI_{EUR}$ and $PGI_{EXT1}$ (correlation of $Z$: $r$ = 0.996, s.e. = 0.001; correlation of $R^2$: $r$ = 0.985, s.e. = 0.004), between the $PGI_{EUR}$ and $PGI_{EUR-23andMe}$ (correlation of $Z$: $r$ = 0.996, s.e. < 0.001; correlation of $R^2$: $r$ = 0.983, s.e. = 0.004), and between the $PGI_{EUR-23andMe}$ and $PGI_{EXT1-23andMe}$ (correlation of $Z$: $r$ = 0.996, s.e. = 0.001; correlation of $R^2$: $r$ = 0.984, s.e. = 0.004).

However, we found that incremental $R^2$ values systematically laid above the identity line, indicating that PGIs based on larger discovery GWAS generally explained a larger proportion of variance in outcomes, as expected (**Supplementary Table 34**, **Figure S14**).

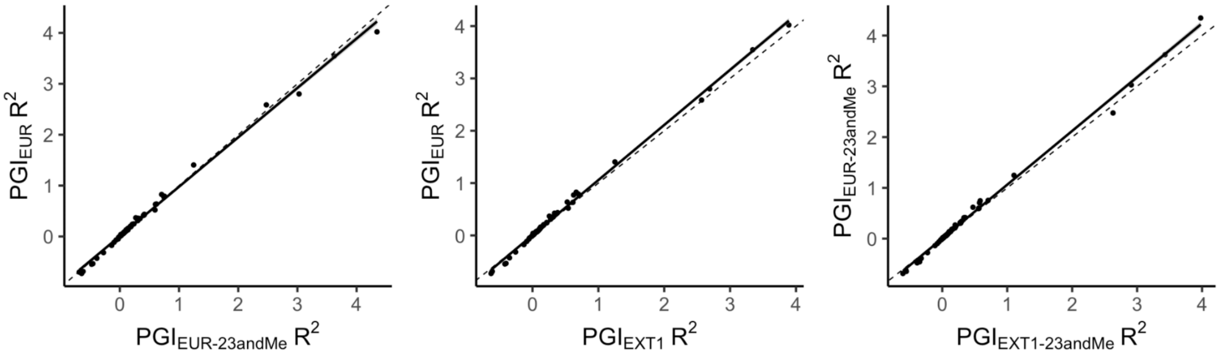


**Figure S14. Incremental variance explained for EXT PGIs derived from alternative discovery GWAS.** Scatter plots depict the correlations between incremental $R^2$ values of the associations between different versions of the EXT PGI and phenotypes in EUR-like participants from COGA and Add Health. $PGI_{EUR}$ denotes the European-ancestry externalizing GWAS from the current study, and $PGI_{EXT1}$ denotes the previous European-ancestry externalizing GWAS[1]. The $PGI_{EUR\text{-}23andMe}$ was generated from the current study after removing 23andMe samples, and $PGI_{EXT1\text{-}23andMe}$ corresponds to a version of the previous externalizing GWAS excluding 23andMe data[51]. The dashed line represents the identity line (intercept = 0, slope = 1), and the solid line represents the line of best fit. Each point corresponds to the incremental $R^2$ for a single outcome.

#### 7.4.4. ABCD

***EUR-like ancestry***. $PGI_{META}$ and $PGI_{EUR}$ were significantly associated with 17 of the 19 investigated phenotypes, excluding ever being offered tobacco and the BIS reward responsiveness score. The coefficients for $PGI_{META}$ and $PGI_{EUR}$ associations were very similar (correlation of coefficients: $r = 1$), with consistent patterns of significance. We observed relatively small differences between $PGI_{META}$ and $PGI_{EUR}$ ($\Delta R^2$ ranged from −0.1% to 0.1%; median change in $\Delta R^2$ = 0.02%). As with the other sections, we focus our reporting on the $PGI_{META}$ results. All results are provided in **Supplementary Table 31.**

The largest effects for $PGI_{META}$ were observed for rule-breaking behavior ($\Delta R^2$ = 2.5% for both parent and youth report), parent-reported externalizing behavior ($\Delta R^2$ = 2.1%), conduct disorder ($\Delta R^2$ = 2.1%), ADHD symptoms ($\Delta R^2$ = 1.7%), aggressive behavior ($\Delta R^2$ = 1.7%), and attentional problems ($\Delta R^2$ = 1.6%). Additional significant youth-reported associations included positive urgency ($\Delta R^2$ = 1.4%), social problems ($\Delta R^2$ = 1.2%), and impulsivity ($\Delta R^2$ = 1.1%). We investigated whether these associations were robust to genetic confounding by testing whether significant population-level associations were still significant after controlling for within-family effects. Only a subset of these associations remained significant in the sibling sample, and none were still significant after adjusting for family fixed effects (**Supplementary Table 31**).

***AFR-like ancestry***. Among participants with AFR-like ancestry, $PGI_{META}$ was significantly associated with 9 of the 19 assessed phenotypes, and all these phenotypes (except for youth-reported engagement in behaviors that violate rules or norms, such as lying, skipping school, or vandalism ) were also significantly associated with $PGI_{AFR}$. Associations were very similar between $PGI_{META}$ and $PGI_{AFR}$ (correlation of coefficients: $r = 0.931$, s.e. = 0.089). Differences in the variance explained by the $PGI_{META}$ and $PGI_{AFR}$ were also minimal ($\Delta R^2$ differences ranged from −0.1% to 0.5%; median $\Delta R^2$ change = 0.2%). All results are provided in **Supplementary Table 31.**

The largest effects were observed between PGIs and parent-reported CBCL externalizing score ($\Delta R^2_{META}$ = 1.3%, $\Delta R^2_{AFR}$= 0.08%), aggressive behavior ($\Delta R^2_{META}$ = 1.2%, $\Delta R^2_{AFR}$= 0.7%), conduct disorder symptoms ($\Delta R^2_{META}$ = 1.2%, $\Delta R^2_{AFR}$= 0.8%), rule-breaking behavior ($\Delta R^2_{META}$ = 1.1%, $\Delta R^2_{AFR}$= 0.9%), and ADHD symptoms ($\Delta R^2_{META}$ = 1.0%, $\Delta R^2_{AFR}$= 0.8%). Additional associations were observed for attention problems ($\Delta R^2_{META}$ = 0.8%, $\Delta R^2_{AFR}$= 0.8%), social problems ($\Delta R^2_{META}$ = 0.6%, $\Delta R^2_{AFR}$= 0.6%), and impulsivity ($\Delta R^2_{META}$ = 0.5%, $\Delta R^2_{AFR}$= 0.5%).

Youth rule-breaking behavior was significant for $PGI_{META}$ ($\Delta R^2_{META}$= 0.7%) but not for the $PGI_{AFR}$. The association between $PGI_{META}$ and ADHD symptoms was the only association that remained significant in the AFR-like ancestry sibling sample, and it was not significant after controlling for family fixed effects.

#### 7.4.5. MCS

We tested associations between EXT PGIs and 11 phenotypes measured in individuals from EUR-like ancestries in MCS. $PGI_{META}$ and $PGI_{EUR}$ were significantly associated with 10 of the 11 phenotypes, but not with heavy drinking. The coefficients for the $PGI_{META}$ and $PGI_{EUR}$ associations were very similar (correlation of coefficients: $r$ = 1), with consistent patterns of significance. We saw relatively small differences between the $PGI_{META}$ and $PGI_{EUR}$ (differences in $\Delta R^2$ ranged from −0.1% to 0.1%, median change in $\Delta R^2$ = 0.01%). As with the other sections, we focus our reporting on the $PGI_{META}$ results. All results are provided in **Supplementary Table 32.**

In the population-level analyses, $PGI_{META}$ was significantly associated with clinical symptoms measured with the SDQ, including externalizing symptoms ($\Delta R^2$ = 5.2%), hyperactivity symptoms ($\Delta R^2$ = 4.1%), and conduct disorder symptoms ($\Delta R^2$ = 4%). $PGI_{META}$ was also associated with several health-risk behaviors, including smoking initiation ($\Delta R^2$ = 3.4%), cannabis initiation ($\Delta R^2$ = 1.2%), ever having sexual intercourse ($\Delta R^2$ = 1%), other substance initiation ($\Delta R^2$ = 0.8%), and alcohol initiation ($\Delta R^2$ = 0.2%), but not with heavy drinking. $PGI_{META}$ was also associated with ever being pregnant ($\Delta R^2$ = 0.7%) and ever being arrested ($\Delta R^2$ = 0.4%).

All associations, except for ever drinking alcohol, remained significant when estimated in a within-family model using a trio design. We evaluated the attenuation of PGI effects by comparing beta coefficients between population-level and within-family models (**Supplementary Table 32**). Briefly, we found that attenuation was minimal and not statistically significant, except for the association with ever being pregnant.

#### 7.4.6. UKB

We tested the association between EXT PGIs and 37 phenotypes in the UKB sibling hold-out sample of EUR-like ancestry individuals. $PGI_{META}$ and $PGI_{EUR}$ were significantly associated with 32 phenotypes, but not with cigarettes per day, happiness (i.e., subjective well-being), and three neuroticism items (suffering from nerves, being a worrier, and worrying for a long time after embarrassment). The coefficients of the associations for the $PGI_{META}$ and $PGI_{EUR}$ were very similar (correlation of coefficients: $r = 1$), with consistent patterns of significance. We observed relatively small differences between $PGI_{META}$ and $PGI_{EUR}$ ($\Delta R^2$ ranged from −0.1% to 0.1%; median change in $\Delta R^2 = 0.007\%$). We present the within-family $PGI_{META}$ results below. All results are provided in **Supplementary Table 33.**

In the UKB sibling hold-out sample, the incremental $R^2$ values for the $PGI_{META}$ associations ranged from very small (e.g., feelings easily hurt, $\Delta R^2 = 0.01\%$) to substantial (e.g., lifetime smoking initiation, $\Delta R^2 = 5.6\%$). The associations with a $\Delta R^2 \geq 2\%$ included lifetime smoking initiation ($\Delta R^2 = 5.6\%$), age at first sexual intercourse ($\Delta R^2 = 3.5\%$), age at first birth ($\Delta R^2 = 2.9\%$), teenage conception ($\Delta R^2 = 2.1\%$), and number of sexual partners ($\Delta R^2 = 2.1\%$).

When controlling for family fixed effects, 53% of these associations remained significant after Bonferroni correction (17/32). Some of these associations remained significant despite showing evidence of attenuation, including age at first birth (among females with children), age at first sexual intercourse, automobile speeding propensity, overall health, and renting from a local authority or a private landlord. Other associations remained significant while also showing no meaningful attenuation.

#### 7.4.7. BioVU

***EUR-like ancestry.*** Among EUR-like ancestry individuals in BioVU, we tested for associations between $PGI_{META}$ and 1,440 medical outcomes, finding that 223 were significantly associated at Bonferroni-corrected significance ($P \leq 3.47e\text{-}5$; **Supplementary Table 36**). As expected, many associations were identified in the mental disorder category ($k = 29$). Noteworthy associations in that group are tobacco use disorder ($N_{cases} = 9{,}027$, OR = 1.51, $P = 1.30e\text{-}248$), substance addiction and disorders ($N_{cases} = 2{,}915$, OR = 1.42, $P = 8.74e\text{-}72$), alcoholism ($N_{cases} = 1{,}330$, OR = 1.35, $P = 5.42e\text{-}26$), mood disorders ($N_{cases} = 12{,}040$, OR = 1.16, $P = 1.28e\text{-}46$) and suicidal ideation or attempt ($N_{cases} = 813$, OR = 1.35, $P = 1.21e\text{-}16$), as well as bipolar disorder ($N_{cases} = 2{,}046$, OR = 1.28, $P = 5.98e\text{-}27$) and major depressive disorder ($N_{cases} = 6{,}143$, OR = 1.13, $P = 9.83e\text{-}18$). $PGI_{META}$ was also significantly associated with ADHD ($N_{cases} = 1{,}396$, OR = 1.22, $P = 3.08e\text{-}12$) and conduct disorders ($N_{cases} = 536$, OR = 1.28, $P = 2.18e\text{-}8$).

$PGI_{META}$ was also associated with a range of different medical outcomes in various disease categories, including the circulatory system ($k$ = 29), such as ischemic heart disease ($N_{cases}$ = 12,223, OR = 1.13, $P$ = 1.20e-25); respiratory diseases ($k$ = 27), such as chronic airway obstruction ($N_{cases}$ = 5,633, OR = 1.31, $P$ = 1.33e-75); endocrine/metabolic conditions ($k$ = 20), such as type 2 diabetes ($N_{cases}$ = 10,383, OR = 1.07, $P$ = 1.91e-9); digestive diseases ($k$ = 17), including cirrhosis of liver (e.g., $N_{cases}$ = 2,178, OR = 1.21, $P$ = 4.40e-18); neurological diseases ($k$ = 6), such as chronic pain ($N_{cases}$ = 6,406, OR = 1.10, $P$ = 2.17e-12); neoplasms ($k$ = 13), including lung cancer ($N_{cases}$ = 2,569, OR = 1.23, $P$ = 8.79e-24); and other categories including sense organs ($k$ = 13), dermatologic ($k$ = 11), musculoskeletal ($k$ = 10), genitourinary ($k$ = 10), injuries and poisonings ($k$ = 9), hematopoietic ($k$ = 6), and symptoms ($k$ = 5).

$PGI_{META}$ was also associated with medical outcomes that are linked to risky health behaviors. For example, increased drinking and alcohol use disorder may explain the association with chronic liver disease and cirrhosis ($N_{cases}$ = 4,422, OR = 1.13, $P$ = 1.24e-14), injuries including fracture of ribs ($N_{cases}$ = 603, OR = 1.22, $P$ = 1.51e-6), and skull and face fracture and other intracranial injury ($N_{cases}$ = 916, OR = 1.17, $P$ = 4.64e-6). Lifetime smoking initiation likely explains the associations with respiratory diseases known to be caused by tobacco smoking, such as lung cancer ($N_{cases}$ = 2,569 OR = 1.23, $P$ = 8.79e-24), emphysema ($N_{cases}$ = 1,547, OR = 1.33, $P$ = 2.52e-26), and chronic airway obstruction ($N_{cases}$ = 5,633, OR = 1.33, $P$ = 1.33e-75). Notably, we identified associations with viral hepatitis C ($N_{cases}$ = 1,340, OR = 1.56, $P$ =8.78e-55) and HIV diagnosis ($N_{cases}$ = 701, OR = 1.46, $P$ = 9.81e-22), which could be due to riskier sexual behaviors or unsafe substance use practices, such as needle sharing.

As a sensitivity analysis, we also tested for associations between $PGI_{EUR}$ and medical outcomes, finding 218 Bonferroni-significant associations with virtually identical results in terms of magnitude and significance across all categories (**Supplementary Table 36**).

*Age-stratified analyses.* We found that many associations between $PGI_{META}$ and medical outcomes differed across developmental windows (**Supplementary Table 37**). In the 0–11 age group, we identified 4 associations that included ADHD ($N_{cases}$ = 374, OR = 1.32, $P$ = 2.32e-7), muscle weakness ($N_{cases}$ = 441, OR = 0.81, $P$ =1.75e-5), allergic reaction to food ($N_{cases}$ = 163, OR = 0.73, $P$ = 9.56e-5), and mood disorders ($N_{cases}$ = 201, OR = 1.31, $P$ = 1.67e-4). In the 12–18 age group, we identified 39 associations across 10 categories, of which 35.9% (14/39) were mental disorders, including posttraumatic stress disorder ($N_{cases}$ = 159, OR = 1.74, $P$ = 2.20e-11), substance addiction and disorders ($N_{cases}$ = 108, OR = 1.46, $P$ = 1.33e-4), conduct disorders ($N_{cases}$ = 155, OR = 1.45, $P$ = 7.98e-6), suicidal ideation or attempt ($N_{cases}$ = 257, OR = 1.39, $P$ = 6.31e-7), and attention deficit hyperactivity disorder ($N_{cases}$ = 342, OR = 1.38, $P$ = 3.18e-8). In the 19–25 age group, we identified 39 associations across 14 categories, the majority falling within the pregnancy complications category (23%) and the mental disorder category (21%), including substance addiction and disorders ($N_{cases}$ = 191, OR = 1.77, $P$ = 3.23e-14), tobacco use disorder ($N_{cases}$ = 213, OR = 1.76, $P$ = 2.11e-15) and pregnancy complications, such as hemorrhage in early pregnancy ($N_{cases}$ = 377, OR = 1.59, $P$ = 4.34e-17). In the 26–40 age group, we identified 128 associations across 16 categories, including mental disorders (e.g., tobacco use disorder: $N_{cases}$ = 861, OR = 1.66, $P$ = 4.44e-43; bipolar disorder: $N_{cases}$ = 437, *OR* = 1.44, $P$ = 3.06e-13), infectious diseases (*e.g.*, viral hepatitis: $N_{cases}$ = 188, OR = 1.76, $P$ = 1.07e-13), and neurological conditions (e.g., pain: $N_{cases}$ = 964, OR = 1.21, $P$ = 1.60e-8). The highest number of associations ($k$ = 223) was observed in the 41–60 age group, spanning 16 categories. Categories with the largest number of significant associations included circulatory ($k$ = 32), respiratory ($k$ = 26), mental ($k$ = 24), and neurological ($k$ = 6) disorders. In the 61–100 age group, we identified 170 associations across 15 categories, with the highest counts in the circulatory system ($k$ = 31) and respiratory ($k$ = 23) categories.

*Sensitivity analyses.* After including socioeconomic status and comorbidities as covariates, we found that $PGI_{META}$ was still significantly associated with many of the originally observed outcomes (**Supplementary Table 38**). Of the 223 associations identified in the primary PheWAS, 205 (92.93%) remained FDR-significant after adjusting for the individual's area deprivation index, 221 (99.10%) after adjusting for the comorbidity score, and 106 (48.18%) after adjusting for diagnoses of alcohol, tobacco, and other substance use disorders.

***AFR-like ancestry***. Among AFR-like ancestry individuals in BioVU, we tested for associations between $PGI_{AFR}$ and 614 medical outcomes, identifying only a significant relationship with tobacco use disorder ($N_{cases}$ = 1,051, OR = 1.21, $P$ = 8.37e-7). The association with tobacco use disorder was also significant for the $PGI_{META}$ ($N_{cases}$ = 1,051, OR = 1.38, $P$ = 4.89e-16), along with two additional associations: substance addiction and disorders ($N_{cases}$ = 488, OR = 1.42, $P$ = 2.83e-10) and mood disorders ($N_{cases}$ = 1,376, OR = 1.15, $P$ = 4.18e-5) (**Supplementary Table 36**).

### 8. Deviations from the preregistration

**Section authors:** Camille M. Williams & Travis T. Mallard

We deviated from our preregistered analysis plan (OSF: https://osf.io/7pfgj/) in order to incorporate newly available data and methods. The deviations below were undertaken to improve genomic discovery and increase the predictive accuracy of polygenic indices, while preserving the study's core aims.

1. **Replacing CANN with CUD.** We replaced CANN with CUD as a model indicator because CUD is more proximate to the externalizing spectrum (**Supplementary Section 3**).
2. **Handling of highly correlated indicators.** The preregistration noted that we would consider combining indicators with high genetic overlap ($r_g \geq 0.70$), but this was poorly articulated and did not specify that traits must also conceptually and phenotypically overlap (e.g., AUD case status, AUDIT-P scores). Although the EUR-like CUD and ALCP GWAS were correlated at $r_g = 0.728$, we retained them as distinct indicators given their theoretical separability.
3. **Factor scaling.** For identification, we scaled the latent externalizing factor by fixing the CUD loading to 1 (rather than NSEX, as in EXT1). In the present models, CUD exhibited the highest loading in both EUR- and AFR-like analyses, making it the more appropriate choice (**Supplementary Section 3**).
4. **Cross-ancestry discovery approach.** We did not implement LOG-TRAM as preregistered. Instead, we used downstream, ancestry-aware methods that take ancestry-specific GWAS and reference panels as inputs for genetic discovery (**Supplementary Sections 4–5**) and polygenic prediction (**Supplementary Section 7**). We conduct a fixed-effects cross-ancestry meta-analysis with METAL (**Figure 2**) for the fine-mapping analyses to identify independent regions across ancestries harboring significant associations, for data visualization, and for the binomial sign concordance test in the replication analyses (**Supplementary Section 5**).
5. **Replication GWAS target.** We did not use the preregistered antisocial behavior GWAS[92] due to the substantial sample overlap with our discovery datasets (e.g., iPSYCH, FinGen, ALSPAC, QIMR, PGC). Instead, we used an externalizing GWAS from the All of Us Research Program for the replication analyses (**Supplementary Section 5**). The antisocial

behavior summary statistics were instead used to evaluate the convergent validity of our latent factor via genetic correlation analyses (**Supplementary Section 3**).

6. **Bioannotation and bioinformatic pipeline.** Rather than relying on the web-based FUMA platform for biological contextualization (as originally preregistered), we applied a variety of newer, state-of-the-art methods for fine-mapping, gene prioritization, and functional enrichment at both genome-wide and prioritized-set levels of analysis (**Supplementary Section 6**).
